# Effectiveness and Safety of Type- and Energy-based Extracorporeal Shockwave Therapy in Clinical Practice: Umbrella Review and Evidence Mapping

**DOI:** 10.1101/2024.01.07.24300948

**Authors:** Qingcong Mo, Jialing Zheng, Fangfei Hu, Peng Gao, Tong Wu, Simeng Wei, Jiaqi Zhang, Chun-Zhi Tang, Jihua Zou, Xiaoyan Zheng

**Author notes:** Corresponding authors: Xiaoyan Zheng; School of Rehabilitation Sciences, Southern Medical University, 1023-1063 Shatai South Road, Guangzhou, China. E-mail address.; Jihua Zou; School of Rehabilitation Sciences, Department of Rehabilitation Medicine, Zhujiang Hospital, Southern Medical University, 1023-1063 Shatai South Road, Guangzhou, China; Faculty of Health and Social Sciences, The Hong Kong Polytechnic University, No.11 Yucai Road, Hong Kong, China. E-mail address.; Chun-Zhi Tang; Medical College of Acu-Moxi and Rehabilitation, Guangzhou University of Chinese Medicine, No.232 Waihuandong Road, Guangzhou, China. Qingcong Mo and Jialing Zheng contributed equally to this work.

## Abstract

**Background:** The role of distinct type- and energy-based extracorporeal shockwave therapy (ESWT) in clinical practice is unclear.

**Objectives:** To appraise meta-analytically determined effectiveness and safety of type- and energy-based ESWT for diseases or conditions, and visualize evidence maps of findings.

**Methods:** Nine online databases and reference lists were systematically searched for systematic reviews (SRs) of randomized controlled trials (RCTs) evaluating the effectiveness or safety of ESWT from inception to September 2023. SRs were then updated if up-to-date RCTs were eligible. Overall effects were re-estimated using random-effects model and reported as relative risk or standardized mean difference with 95% confidence intervals. Methodological quality, certainty of evidence, and safety were assessed with AMSTAR 2, GRADE tool, and MedDRA, respectively.

**Results:** Our research identified 210 relevant SRs encompassing 636 RCTs and 41649 participants across 7 therapeutic areas and 37 diseases and conditions. Methodological quality of most published SRs was low or critically low. Four treatment statuses of type- and energy-based ESWT were identified, including potential dominant efficacy (plantar fasciitis, erectile dysfunction, lateral epicondylitis, knee osteoarthritis, frozen shoulder, cerebral palsy spasticity, post-stroke lower limb spasticity; GRADE moderate), potential positive efficacy (chronic prostatitis/chronic pelvic pain syndrome, myofascial pain syndrome, patellar tendinopathy, achilles tendinopathy, stenosing tenosynovitis, frozen shoulder, rotator cuff tear, cerebral palsy spasticity, post-stroke upper limb spasticity, cervical spondylotic radiculopathy; GRADE low or very low), potential similar efficacy (osteonecrosis of the femoral head, plantar heel pain, patellar tendinopathy; GRADE low or very low), and potential adverse efficacy (patellar tendinopathy; GRADE very low). Along with courses of ESWT treatment, pain, flushing, and swelling were the most prevalent side effects and serious adverse reactions were limited.

**Conclusion:** Variable type- and energy-based ESWT is probably effective and safe in clinical practice. Due to lack of available data and high certainty in current evidence, future research should prioritize large-scale and well-designed studies.

**Registration:** PROSPERO number CRD42023477234

## Introduction

Researchers and clinicians have made a wide investigation of extracorporeal shockwave therapy (ESWT) in more than one-third of the countries/regions (36%) worldwide, a form of noninvasive physiotherapy [1]. Since the initial management of ESWT in urological application as extracorporeal shock wave lithotripsy around the 1980s, its utilization subsequently expanded to a wide range of conditions [2]. Following the discovery of mechanisms, plentiful pre-clinical studies elucidated the physical characteristics, chemical reactions, and biological effects of ESWT [3–6]. Supported by these researches, physiatrists have utilized ESWT to treat multiple pathological conditions, involving musculoskeletal, neurological, genitourinary, dermatological, circulatory, and dental disorders [7–13]. As a result, numerous clinical evidence was available to evaluate its effectiveness and safety.

Based on the extensive application of ESWT, researchers have conducted considerable systematic reviews (SRs) and meta-analyses to synthesize the findings from primary studies over decades. Accordingly, indications and contraindications of ESWT were clarified for clinical practice by the International Society for Medical Shockwave Treatment (ISMST) [14] and the Shock Wave Medical Professional Committee of Chinese Research Hospital Association [15]. Analogously, interventional guidance of ESWT summarized its widespread application in diseases or conditions as well [16–22]. However, recommendations from these guidelines are insufficient to answer questions of optimal treatment parameters that are yet to be determined, as all of them provided a variable range of applications. The exact protocol of ESWT between physical parameters and clinical outcomes remains unclear [14,23]. Under the circumstances, the uncertainty of optimal determination implicates device setting of specific type and energy level, compromising the clinical value of the anticipated dose’s effect [14,23].

As the number of SRs increased rapidly up to now, physiatrists may spend vast amounts of valuable research resources identifying evidence on the optimal type- and energy-based ESWT for diseases or conditions. To overcome the gap of knowledge, umbrella review, a method of reviewing previous SRs, can be an appropriate and suitable option [24,25]. It is capable of comparing type- and energy-based ESWT to other interventions following a uniform approach and representing the highest level of comprehensive and systematic summary of evidence [26,27]. Meanwhile, evidence mapping is a useful tool to characterize and synthesize evidence through user-friendly visual graphics [28,29]. Integration of umbrella review and evidence mapping develops synergy effects in ensuring overall investigation [30,31].

To date, no umbrella review and evidence mapping reporting a comprehensive summary of the current evidence regarding type- and energy-based ESWT exists. High-quality evidence is urgently warranted for it to promote standard stewardship as well [32,33]. Therefore, we conducted this study to assess the effectiveness and safety of type- and energy-based ESWT, identify existing evidence gaps, promote knowledge dissemination, and guide future research.

## Methods

This umbrella review and evidence mapping was prospectively registered in the International Prospective Register of Systematic Reviews (PROSPERO-CRD42023477234). It was carried out in accordance with the Preferred Reporting Items for Systematic Reviews and Meta-Analysis (PRISMA) 2020 checklist (Supplementary Figure S1) [34] and the Cochrane Collaboration Handbook [35].

## Definition

### Extracorporeal shock wave therapy

Through a special device, ESWT is a procedure that presses against the skin by a probe and then propagates the energy until it meets the target affected areas [2]. Focused ESWT involves the transmission of shock waves in a narrow pattern that converges it into particular depth within the body [14]. In contrast, radial ESWT pressure waves are propagated at maximum pressure on the body surface in a non-focused or diffuse pattern [14]. We included 4 kinds of devices in conformity with focused or radial shockwaves categorized by working principles (electrohydraulic, piezoelectric, electromagnetic, and air pressure) [2,14]. Extracorporeal high-intensity focused ultrasound therapy and cardiac shockwave therapy are out of scope in this overview due to distinct devices.

### Disease or condition

Guided by the International Classification of Diseases 11th revision (ICD-11) [36], the term "disease and condition" refers to medically definite diseases or symptoms encountered by people and no standard conditions were established.

### Outcome

The core outcome set (https://comet-initiative.org/) provides a standard process for the selection, collection, and reporting of outcomes to determine the certain core outcome for clinical practice [37]. We selected it as primary outcome given the ability to reduce the risk of heterogeneity, inconsistency, and outcome-reporting bias between trials. Additionally, the patient-reported outcome measures [38,39] and patient-important outcomes [40,41] are capable of capturing patient perspectives surrounding their symptoms, functional status, and quality of life. They are used if interventions for diseases or conditions are not assessed by the core outcome set. The overall efficacy rate was excluded due to its instability, which has different assessment standards across SRs.

### Search strategy

Our search was conducted in the following electronic databases (Epistemonikos database, Web of Science (WOS), Scopus database, Chinese National Knowledge Infrastructure (CNKI), WANFANG Database, Chinese Scientific Journal Database (VIP), Chinese Biomedical Literature Database (CBM)) from inception to September 2023 for SRs. We also searched PROSPERO to identify any registered or yet unpublished SRs, as well as searching the grey literature through OpenGrey. Further, we hand-searched the reference lists of included studies for any eligible cited SRs in Database of International Science Citation, Science Citation Index, and Google Scholar. Medline, Embase, Cochrane Central Register of Controlled Trials, WOS, CNKI, CBM, VIP, WANFANG databases were searched during the retrieval period for newly published randomized controlled trials (RCTs) that were eligible for updating SRs. The search strategy was devised consisting of an integration of MeSH terms, keywords and free text terms related to ESWT following the guidance from ISMST: (extracorporeal shockwave therapy OR shock wave OR pressure wave OR ESWT) AND (systematic review OR meta-analysis) [42]. There were no language restrictions.

Two blinded independent literature reviewers (MQC and ZJL) screened titles and abstracts of SRs and up-to-date RCTs output through the EndNote V.21 software to identify eligible articles. The promising studies were downloaded and evaluated against the specified inclusion criteria by full-text reading.

Using Gwet’s AC1 statistics and associated 95% confidence interval (CI) [43], one reviewer (HFF) subsequently examined the agreement on study selection between two authors (MQC and ZJL). By applying two-tailed analysis, Fisher’s exact test was performed to determine its statistical significance (P value). R language (version 4.3.2) was used for its evaluation.

### Criteria for including

Standard-compliant SRs for ESWT which conducted meta-analyses of RCTs were eligible for inclusion. The criteria were as follows: (1) Full text available in at least one electronic database or citation, (2) Published in the formal journal, dissertation and conference, (3) Fulfill at least one outcome criterion, (4) Details accessible for primary RCTs in SRs. Overviews and protocols of SRs, narrative and scoping reviews, network meta-analyses, or studies published after September 2023 were excluded.

Eligible SRs and up-to-date RCTs were included if they conducted any intervention containing single or adjunctive ESWT as core treatment group. No strict limitations on the type of control group, yet we excluded ESWT as an intervention in this group. Compared to the treatment group, control group may include at least one of the following therapies: (1) no treatment, (2) standard care or usual care, (3) sham ESWT or placebo, (4) medication therapy or drug injection, (5) traditional Chinese medicine external therapy (e.g. acupuncture) or internal therapy (e.g. decoction), (6) other intervention (e.g. rehabilitation, surgical treatment). Up-to-date RCTs that did not offer energy usage (energy flux density or pressure field) or overlapped with the same eligible RCTs included in SRs were excluded.

Our team made no restrictions on human participants, ensuring that individuals of all ages, genders, regions and any conditions were included. And we forbade animals from participating.

### Data extraction

Independent two groups of reviewers (Group 1, ZJL, GP and ZJQ; Group 2, HFF, WTY and WSM) investigated SRs and extracted data in standardized tables based on predefined criteria. Data concerning the first author’s name, publication year, country of the first author’s affiliation, disease or condition, number of included primary RCTs and participants, comparison, outcome measurement, concrete intervention, method of implementation, study conclusion, evidence assessment tool and adverse reactions (ADRs) were extracted. Furthermore, we extracted the primary study specific data from SRs corresponding to concrete comparisons with outcomes. Discrepancies considered potentially pertinent by at least one reviewer were discussed and adjudicated for a shared consensus, if needed, the reviewer (ZXY) arbitrated.

### Methods and evidence assessment

A Measurement Tool to Assess Systematic Reviews (AMSTAR 2) [44] and the Grading of Recommendations Assessment, Development and Evaluation (GRADE) [45] contribute to measuring the methodological quality and certainty of evidence, respectively. We selected originally recommended items as critical items (2, 4, 7, 9, 11, 13 and 15) in AMSTAR 2. Methodological quality of SRs was judged and categorized by two groups of reviewers (Group 1, ZJL, GP and ZJQ; Group 2, HFF, WTY and WSM) as ‘high,’ ‘moderate,’ ‘low,’ or ‘critically low.’ Meanwhile, if SRs failed to assess the certainty of each outcome that was estimated, two groups of same reviewers re-evaluated and graded it by GRADE as ‘high,’ ‘moderate,’ ‘low,’ or ‘very low.’ For up-to-date RCTs, the Cochrane Risk of Bias assessment tool 2.0 (ROB 2.0) was used to evaluate its quality [46].

### Subgroup classification and re-estimation

#### Subgroup classification

To be applicable in real-world clinical practice, the following prespecified subgroup analyses were conducted based on impulse transmitted from ESWT devices: (1) single focused ESWT, (2) single radial ESWT, (3) adjunctive focused ESWT, (4) adjunctive radial ESWT, (5) single or adjunctive unknown ESWT. In addition, in each predetermined subgroup, we defined three energy levels (high, moderate, and low energy) in accordance with energy flux density or pressure field for in-depth classification (Supplementary Methods S1). Given that there is no global consensus on the energy parameters, we followed the categorization provided by Rompe et al. [47] and Avendaño-Coy et al [48]. It was given precedence over our definition if researchers predetermined their intensity usage. If RCTs offered an energy flux density or pressure field range, we selected its average value for analysis.

#### Subgroup data re-estimation

Based on the subgroup classification of comparisons, the primary study specific data for outcomes were re-estimated following strategy: (a) If outcomes were dichotomous or continuous, we utilized Review Manager V.5.4 software to re-estimated data using random-effects model in the form of relative risk (RR) or standardized mean difference (SMD) along with 95% CI, respectively, (b) If the latest SRs did not include the same comparisons as previous SRs, or if it can be updated by supplementing with up-to-date RCTs, we integrated the estimated summary effect of these SRs and RCTs, (c) If the comparisons in SRs contained different follow-up periods, we combined it for overall effects. We graded the re-estimated magnitude of summary effect on RR or SMD as ‘large,’ ‘moderate,’ ‘small,’ or ‘very small’ [49,50] (Supplementary Methods S1). The heterogeneity (Cochran’s Q test P value and I-square statistic) of outcomes were documented and outcomes without statistical significance (P>0.05) were excluded. Publication bias was measured using Egger’s regression test.

We linked re-estimated overall effect with evidentiary certainty in each outcome, complemented following control group interventions. Further, we classified treatment statuses of type- and energy-based ESWT into four categories following the criteria: diseases or conditions with potential dominant efficacy, potential positive efficacy, potential similar efficacy, and potential adverse efficacy (Table 1).

**Table 1.** Classification criteria of treatment statuses. GRADE, Grading of Recommendations Assessment, Development and Evaluation; RR, relative risk; SMD, standardized mean difference.

| Treatment status | Description |
| --- | --- |
| Potential dominant efficacy | Large or moderate re-estimated summary effect size ($RR \geq 2.0$ or $RR \leq 0.5$; $SMD \geq 0.5$ or $SMD \leq -0.5$), total participants $\geq 300$, positive effect (compared to control group), high or moderate certainty by GRADE |
| Potential positive efficacy | Large or moderate re-estimated summary effect size ($RR \geq 2.0$ or $RR \leq 0.5$; $SMD \geq 0.5$ or $SMD \leq -0.5$), total participants $\geq 200$, positive effect (compared to control group), low or very low certainty by GRADE |
| Potential similar efficacy | Small or very small re-estimated summary effect size ($0.5 < RR < 2.0$; $-0.5 < SMD < 0.5$), total participants $\geq 150$, positive or negative effect (compared to control group) |
| Potential adverse efficacy | Large or moderate re-estimated summary effect size ($RR \geq 2.0$ or $RR \leq 0.5$; $SMD \geq 0.5$ or $SMD \leq -0.5$), total participants $\geq 100$, negative effect (compared to control group) |

### Safety analysis

We reported generalized ADRs corresponding to single or adjunctive type- and energy-based ESWT treatments through providing quantitative synthesis of number of RCTs that occurred side effects. If the SRs did not report ADRs, we searched for their included primary RCTs to record it. Then, to provide standard terminology for safety reports, the Medical Dictionary for Regulatory Activities (MedDRA-version 26.1) was utilized to describe and classify included ADRs by preferred terms and system organ classes [51,52].

## Results

### Study selection and consistency

Figure 1 shows the flowchart of the literature search and study selection. Of 3639 records identified from the electronic databases, 126 from registrations, and 21 from citations, 3576 articles were excluded against our inclusion criteria. Application of searching for up-to-date RCTs yielded a total of 217. The remaining 210 SRs met the eligibility criteria, including 636 RCTs and 41649 individuals. Agreement between the two reviewers (MQC and ZJL) for study selection exhibited high consistency without significant difference (Gwet’s AC1= 0.912, 95% CI 0.869 to 0.955; P=1). A list of included 210 SRs is provided in Supplementary Figure S2 and the reasons for excluded SRs are shown in Supplementary Figure S3.

### Characteristics of studies

A total of 25 countries investigated ESWT in the form of SRs and the majority of the first author’s affiliations come from China (n=136, 65%) (Supplementary Figure S4). The included studies involved 7 therapeutic areas (diseases of the musculoskeletal system or connective tissue, diseases of the nervous system, diseases of the circulatory system, diseases of the genitourinary system, diseases of the skin, symptoms, signs or clinical findings, not elsewhere classified and injury, poisoning or certain other consequences of external causes) with 37 diseases or conditions (Figure 2). Most researchers drew positive or potentially positive conclusions on ESWT towards common clinical outcomes (Supplementary Figure S5). The Cochrane risk of bias tool (n=125, 60%) was the most commonly applied risk of bias assessment tool in SRs, followed by the Physiotherapy evidence database scale (n=32, 15%). Outcome evaluations through GRADE were missed in the majority of SRs (n=183, 87%).

**Figure 1.**
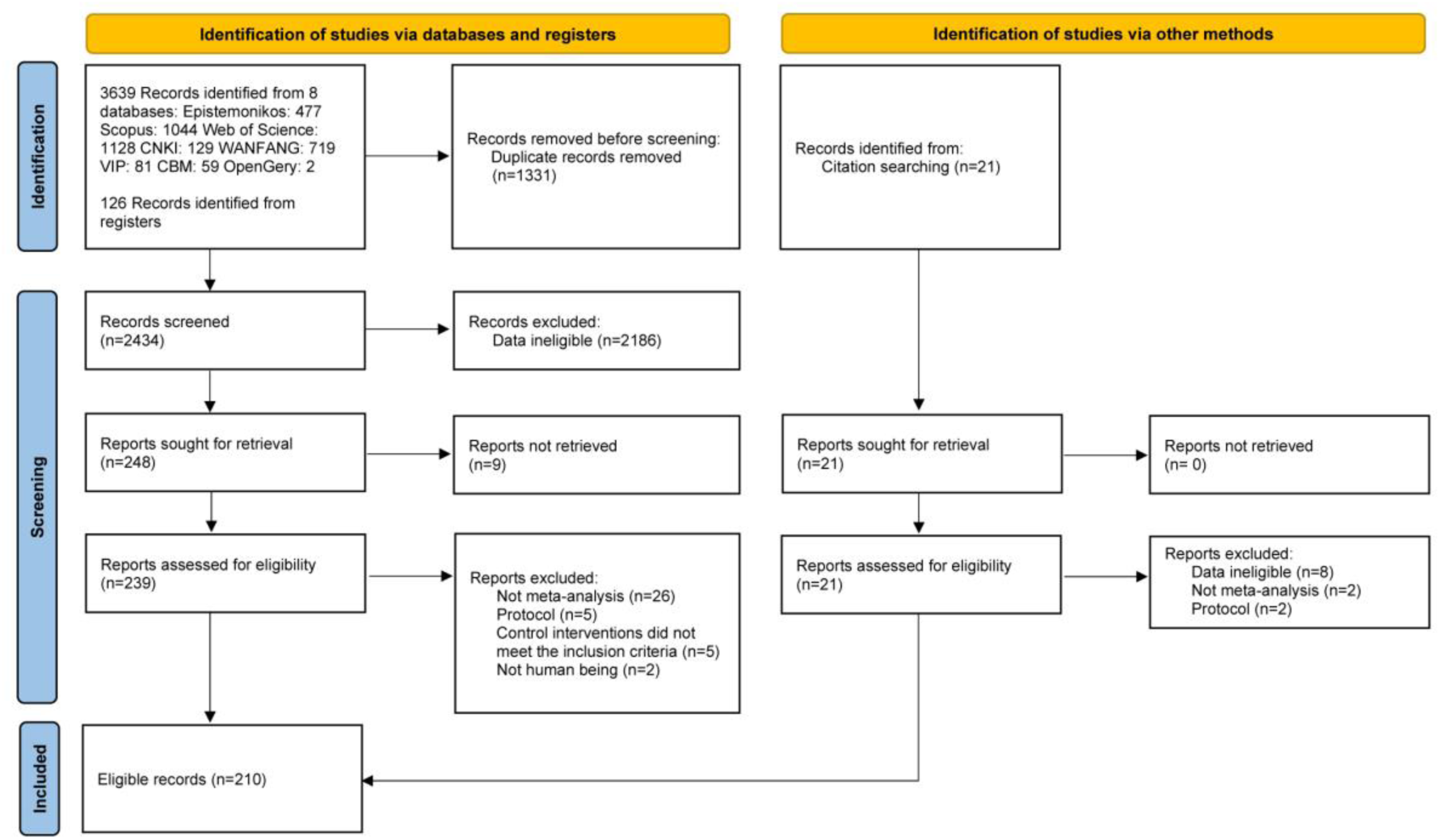
Flowchart of the literature search and study selection. CBM, Chinese Biomedical Literature Database; CNKI, Chinese National Knowledge Infrastructure; VIP, Chinese Scientific Journal Database.

**Figure 2.**
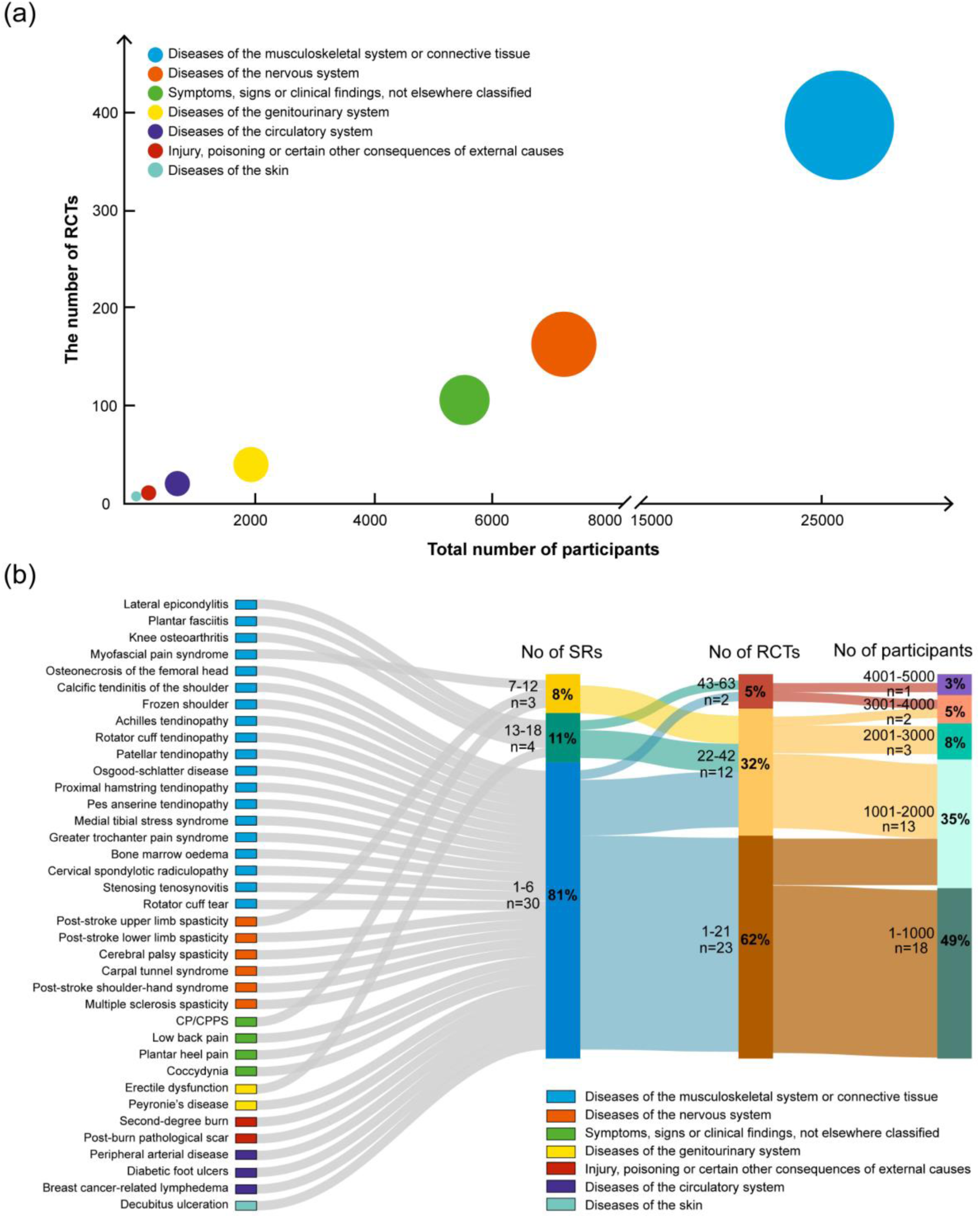
(a) Bubble plot of therapeutic areas corresponding to the number of systematic reviews, randomised controlled trials and participants. The size of pie represents the number of systematic reviews (larger=more studies). RCTs, randomised controlled trials. (b) Sankey diagram of diseases or conditions for the number range of systematic reviews, randomised controlled trials and participants. RCTs, randomised controlled trials; SRs, systematic reviews.

### Bubble plot and Sankey diagram

The bubble plot (Figure 2a) and sankey diagram (Figure 2b) were constructed based on published SRs. The former shows the distribution of evidence among therapeutic areas, along with the number of SRs, RCTs and participants. The most frequently investigated topic is diseases of the musculoskeletal system or connective tissue, followed by diseases of the nervous system. Only two therapeutic areas (diseases of the musculoskeletal system or connective tissue and diseases of the nervous system) include over 100 RCTs with more than 6000 participants. The latter visualizes the associations across diseases or conditions, the number range of SRs, RCTs and participants. Most diseases or conditions are reviewed by 1 to 6 SRs and the majority of SRs incorporate 1 to 21 RCTs with 1 to 1000 participants. Of the 37 diseases or conditions, 7 diseases or conditions (lateral epicondylitis, plantar fasciitis, knee osteoarthritis, myofascial pain syndrome, post-stroke upper limb spasticity, chronic prostatitis/chronic pelvic pain syndrome, erectile dysfunction) are evaluated in more than 6 SRs, with over 22 RCTs involving more than 1000 participants.

### Methodological quality and risk of bias assessment

According to the AMSTAR 2 assessments of the overall inspection for published SRs, their methodological qualities were rated as ‘moderate,’ (n=1, 1%) ‘low,’ (n=18, 12%), ‘critically low’ (n=129, 87%). Most SRs are appraised as low or critically low due to failure to provide a list of excluded studies, register before commencing the review, and assess publication bias. Risk of bias assessment of the majority of up-to-date RCTs is at some concern. Detailed information on items or domains of SRs and up-to-date RCTs is shown in Supplementary Figure S6, 7.

### Diseases or conditions with potential dominant efficacy

#### High and medium energy ESWT

All evidence was graded moderate certainty. In comparison, high energy and medium energy focused ESWT directed at the relief of pain intensity of patients who had plantar fasciitis demonstrated large or moderate effects. Medium energy radial ESWT also represented a moderate effect in decreasing pain intensity for plantar fasciitis versus rehabilitation. Compared to sham ESWT, medium energy radial ESWT had large positive impacts on reducing pain intensity, stiffness, discomfort and improving physical function, endurance of ambulation and activities of daily living in the treatment of knee osteoarthritis. In addition, medium energy radial ESWT had greater effects in reduction in pain intensity, stiffness, discomfort and enhancement in physical function than medication therapy focused on knee osteoarthritis. As an adjuvant therapy, medium energy focused ESWT combined with rehabilitation showed a moderate effect in reduction in pain intensity when treated with frozen shoulder. Details of comparisons and estimated summary effects are shown in Figure 3.

**Figure 3.**
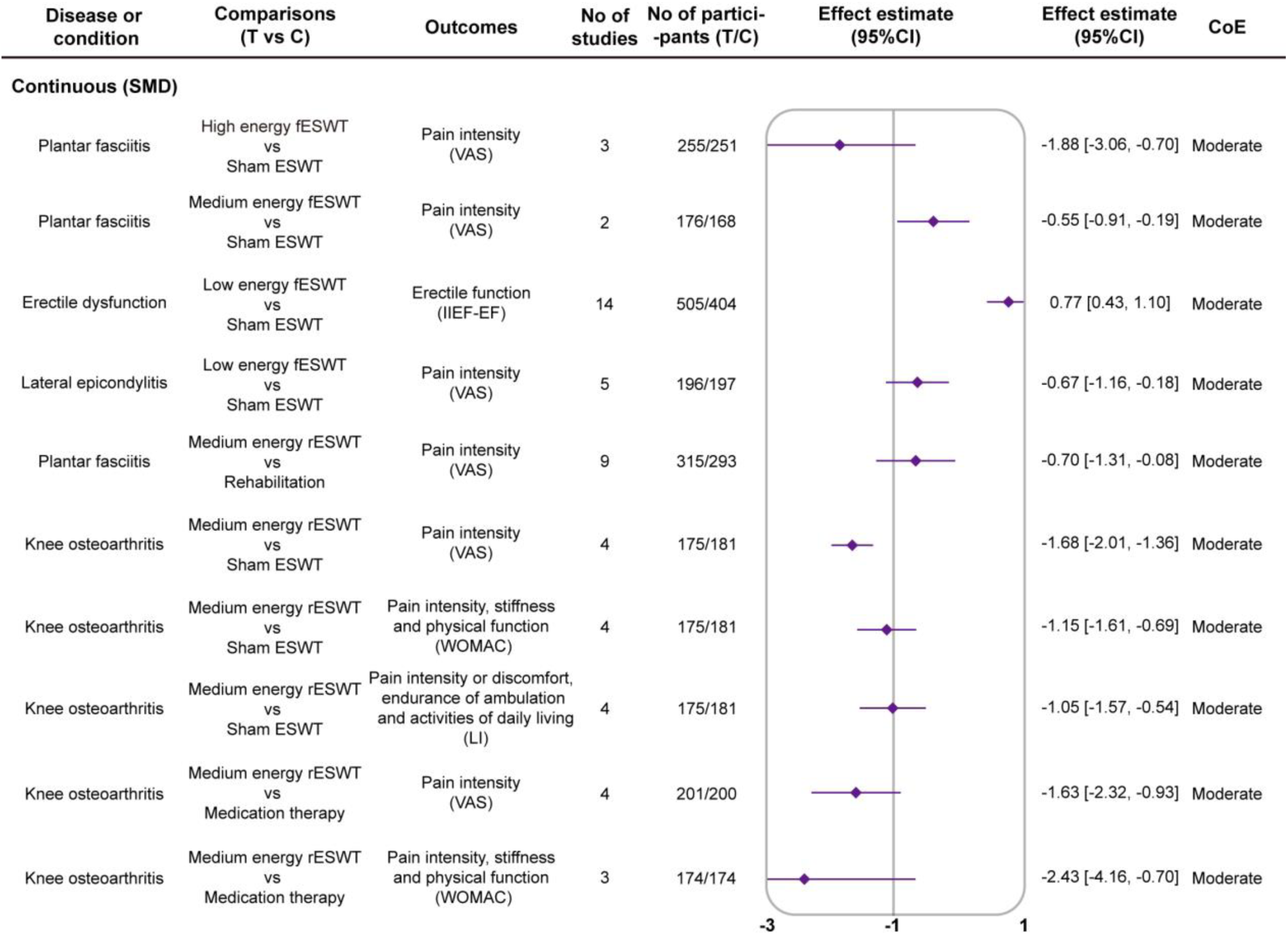
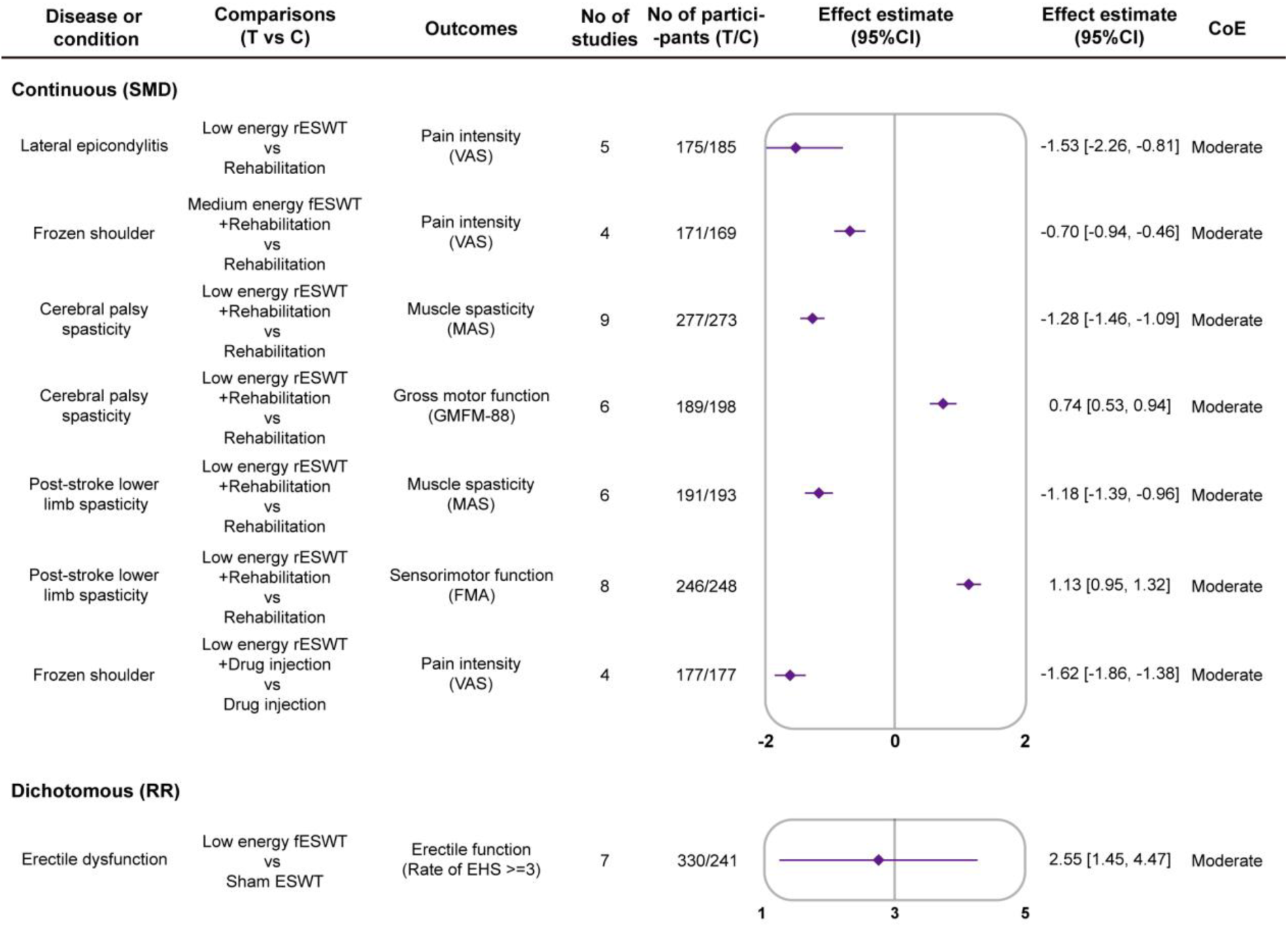
Diseases or conditions with potential dominant efficacy. C, control group; CI, confidence interval; CoE, certainty of evidence assessed using the Grading of Recommendations Assessment, Development and Evaluation approach; EHS, Erection Hardness Score; ESWT, extracorporeal shockwave therapy; fESWT, focused extracorporeal shockwave therapy; FMA, Fugl-Meyer Assessment; GMFM-88, Gross Motor Function Measure-88; IIEF-EF, International Index of Erectile Function-Erectile Function; LI, Lequesne Index; MAS, Modified Ashworth Scale; rESWT, radial extracorporeal shockwave therapy; RR, relative risk; SMD, standardized mean difference; T, treatment group; VAS, Visual Analogue Scale; WOMAC, Western Ontario and McMaster Universities Arthritis Index.

#### Low energy ESWT

All evidence was graded moderate certainty. Low energy focused ESWT had large or moderate positive influences on enhancing erectile function in the management of erectile dysfunction and decreasing pain intensity in patients with lateral epicondylitis. Low energy radial ESWT also represented a large effect in decreasing pain intensity for lateral epicondylitis versus rehabilitation. Integrated with rehabilitation, low energy radial ESWT exhibited large or moderate effects in reducing muscle spasticity and promoting gross motor function for cerebral palsy spasticity patients. Low energy radial ESWT combined with rehabilitation showed large effects in relief of muscle spasticity and improvement in sensorimotor function for post-stroke lower limb spasticity patients. Moreover, low energy radial ESWT with drug injection demonstrated a large effect for treating frozen shoulder in a reduction in pain intensity. Details of comparisons and estimated summary effects are shown in Figure 3.

### Diseases or conditions with potential positive efficacy

#### Medium energy ESWT

Compared to rehabilitation, medium energy focused ESWT directed at relief of global symptoms, pain intensity and increase in quality of life of patients who had chronic prostatitis/chronic pelvic pain syndrome demonstrated large effects (GRADE all very low). Medium energy radial ESWT directed at relief of pain intensity of patients who had myofascial pain syndrome or knee osteoarthritis or patellar tendinopathy demonstrated large or moderate effects in comparison (GRADE all low). Medium energy focused ESWT combined with other interventions showed large effects in relief of pain intensity and improvement in activity of daily living, range of motion, and strength for frozen shoulder patients in comparison (GRADE all low). Medium energy radial ESWT combined with rehabilitation and traditional Chinese medicine external therapy showed large effects in reduction in pain intensity when treated with rotator cuff tear (GRADE very low). Details of comparisons and estimated summary effects are shown in Figure 4.

**Figure 4.**
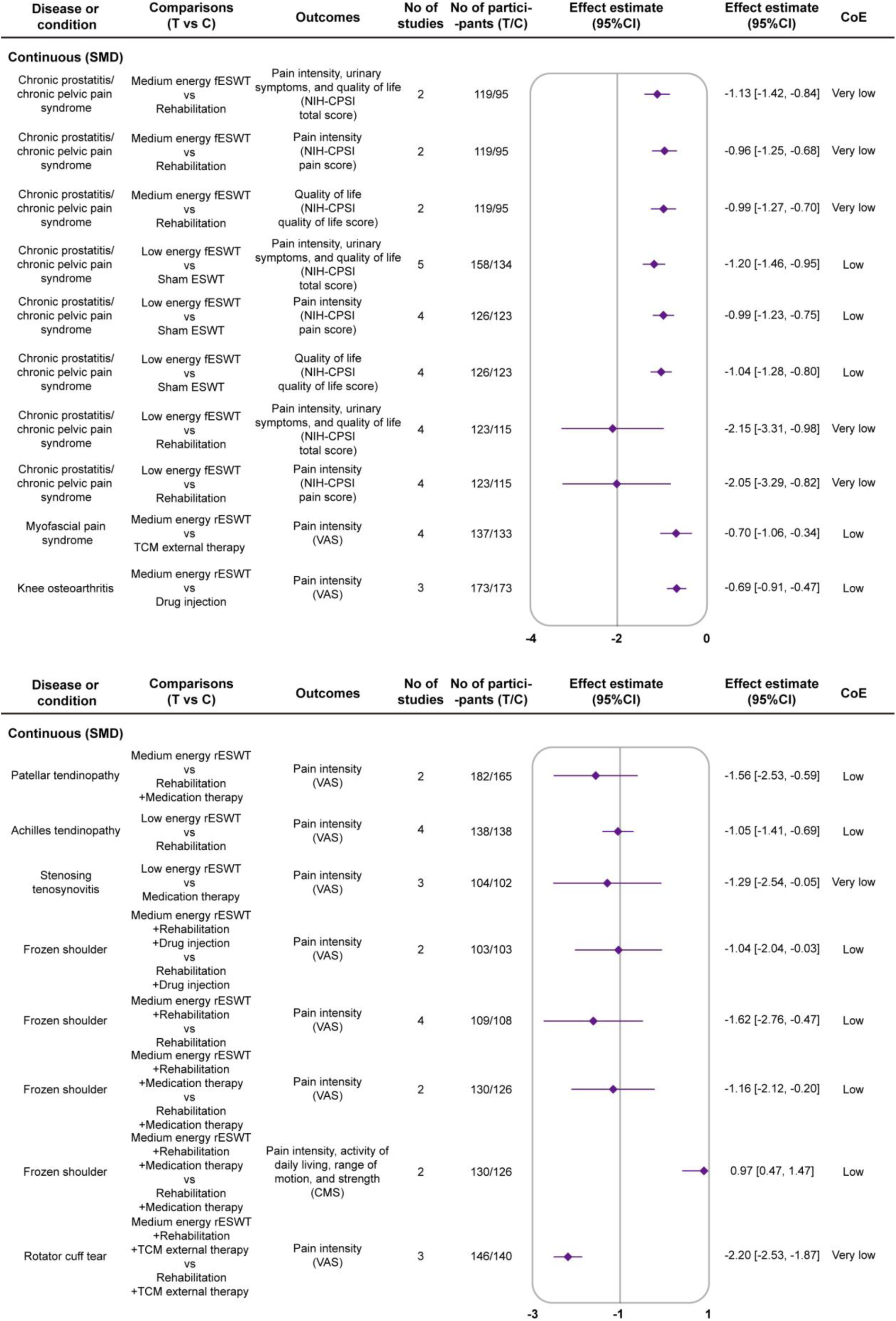
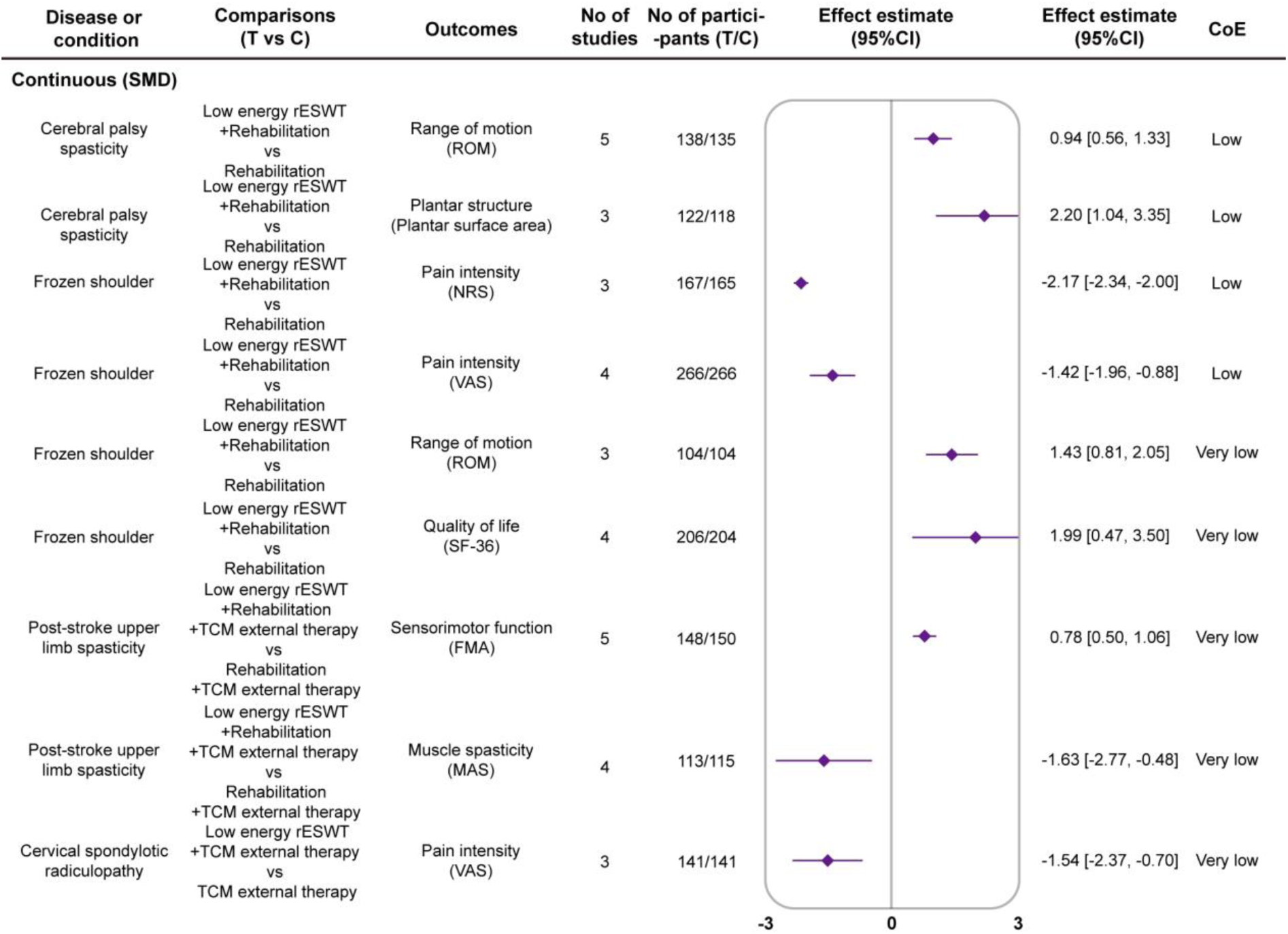
Diseases or conditions with potential positive efficacy. C, control group; CI, confidence interval; CMS, Constant-Murley Score; CoE, certainty of evidence assessed using the Grading of Recommendations Assessment, Development and Evaluation approach; ESWT, extracorporeal shockwave therapy; fESWT, focused extracorporeal shockwave therapy; FMA, Fugl-Meyer Assessment; MAS, Modified Ashworth Scale; NIH-CPSI, National Institutes of Health-Chronic Prostatitis Symptom Index; NRS, Numerical Rating Scale; rESWT, radial extracorporeal shockwave therapy; ROM, Range of Motion; SF-36, 36-item Short-Form; SMD, standardized mean difference; T, treatment group; TCM, traditional Chinese medicine; VAS, Visual Analogue Scale.

#### Low energy ESWT

Compared to sham ESWT, low energy focused ESWT directed at relief of global symptoms, pain intensity and increase in quality of life of patients who had chronic prostatitis/chronic pelvic pain syndrome demonstrated large effects (GRADE all low). Compared to rehabilitation, low energy focused ESWT directed at the relief of global symptoms, and pain intensity of patients who had chronic prostatitis/chronic pelvic pain syndrome also demonstrated large effects (GRADE both very low). Low energy radial ESWT had large positive influences on decreasing pain intensity versus rehabilitation or medication therapy when treated with achilles tendinopathy (GRADE low) or stenosing tenosynovitis (GRADE very low). Integrated with rehabilitation, low energy radial ESWT directed at the increase in range of motion and plantar surface area for cerebral palsy spasticity patients (GRADE both low), reduction in pain intensity (GRADE both low) and improvement in range of motion and quality of life (GRADE both very low) for frozen shoulder patients showed large effects. Low energy radial ESWT combined with rehabilitation and traditional Chinese medicine external therapy showed large or moderate effects in increasing sensorimotor function and decreasing muscle spasticity when treated with post-stroke upper limb spasticity (GRADE both very low). Low energy radial ESWT combined with traditional Chinese medicine external therapy showed a large effect in reduction in pain intensity when treated with cervical spondylotic radiculopathy (GRADE very low). Details of comparisons and estimated summary effects are shown in Figure 4.

### Diseases or conditions with potential similar and potential adverse efficacy

Compared to traditional Chinese medicine internal therapy and medication therapy, high energy focused ESWT directed at decreasing pain intensity, deformity and increasing function, range of motion of patients who had osteonecrosis of the femoral head demonstrated small or very small similar effects (GRADE both very low). Medium energy radial ESWT represented a small similar effect in decreasing pain intensity for plantar heel pain versus sham ESWT (GRADE low). Medium energy radial ESWT combined with rehabilitation showed a small similar effect in decreasing symptom severity for patellar tendinopathy (GRADE low). Details of comparisons and estimated summary effects are shown in Figure 5a.

**Figure 5.**
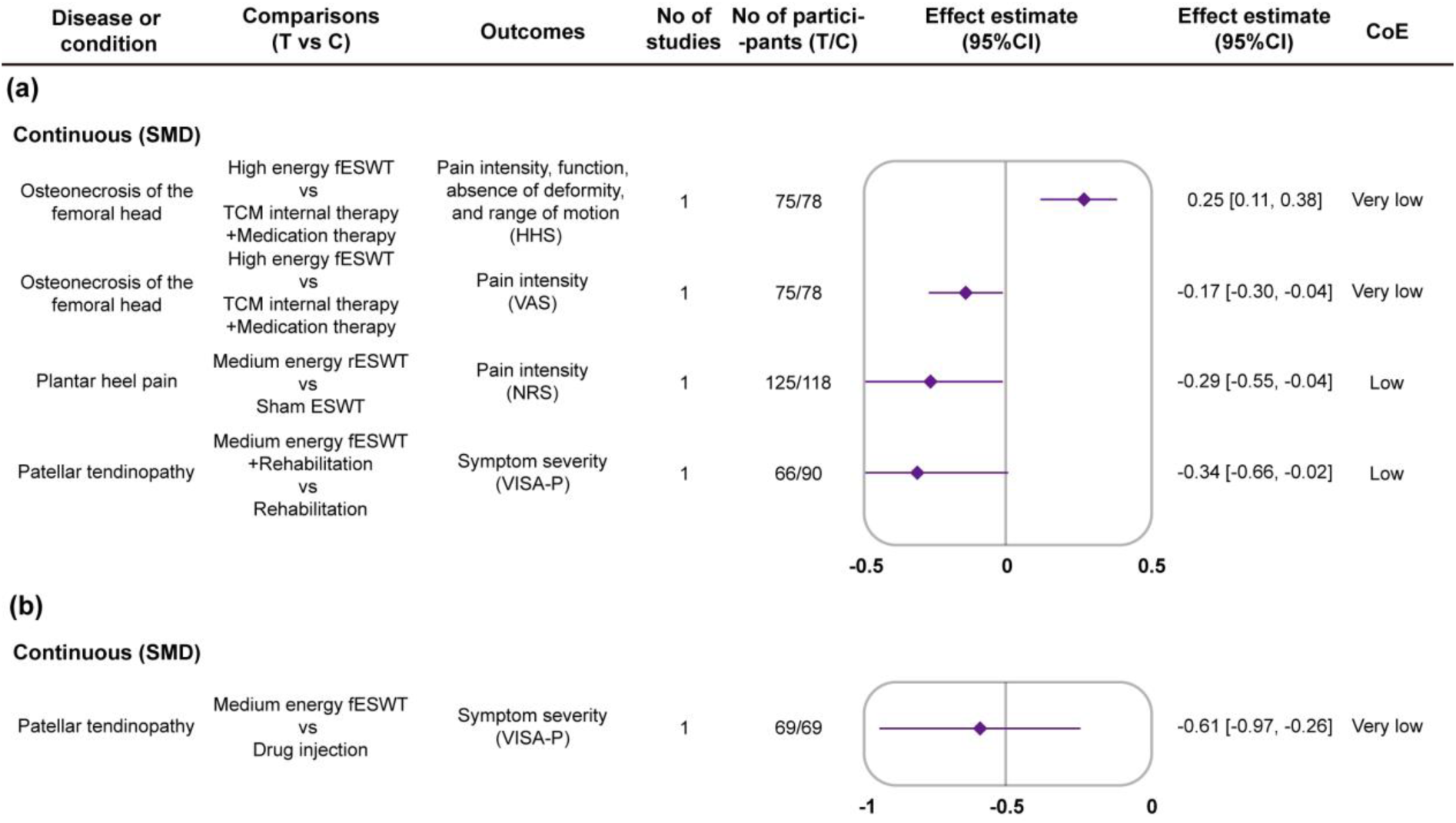
(a) Diseases or conditions with potential similar efficacy. C, control group; CI, confidence interval; CoE, certainty of evidence assessed using the Grading of Recommendations Assessment, Development and Evaluation approach; ESWT, extracorporeal shockwave therapy; fESWT, focused extracorporeal shockwave therapy; HHS, Harris Hip Scale; NRS, Numerical Rating Scale; rESWT, radial extracorporeal shockwave therapy; SMD, standardized mean difference; T, treatment group; TCM, traditional Chinese medicine; VAS, Visual Analogue Scale; VISA-P, Victorian Institute of Sport Assessment-Patella. (b) Diseases or conditions with potential adverse efficacy. C, control group; CI, confidence interval; CoE, certainty of evidence assessed using the Grading of Recommendations Assessment, Development and Evaluation approach; fESWT, focused extracorporeal shockwave therapy; SMD, standardized mean difference; T, treatment group; VISA-P, Victorian Institute of Sport Assessment-Patella.

Medium energy radial ESWT presented a moderate adverse effect in increasing symptom severity for patellar tendinopathy versus drug injection (GRADE very low). Details of comparisons and estimated summary effects are shown in Figure 5b.

### Safety summary

A total of 212 RCTs (33%) reported ADRs related to ESWT (Supplementary Table S2). General disorders and administration site conditions was the system organ classes most frequently appeared (n=89, 42%). The most common side effects during ESWT treatments were pain (n=35, 17%), followed by flushing (n=26, 12%), and swelling (n=18, 9%). Severe ADRs such as haematoma (n=9, 4%), infection (n=2, 1%), and joint dislocation (n=1, 1%) were also observed. More than 20 RCTs reported ADRs in each of the following settings of ESWT: single high and low energy focused ESWT, single medium and low energy radial ESWT, adjunctive low energy radial ESWT, and single or adjunctive high and low energy unknown ESWT. Details of the number of RCTs reporting specific ADRs are in Figure 6.

**Figure 6.**
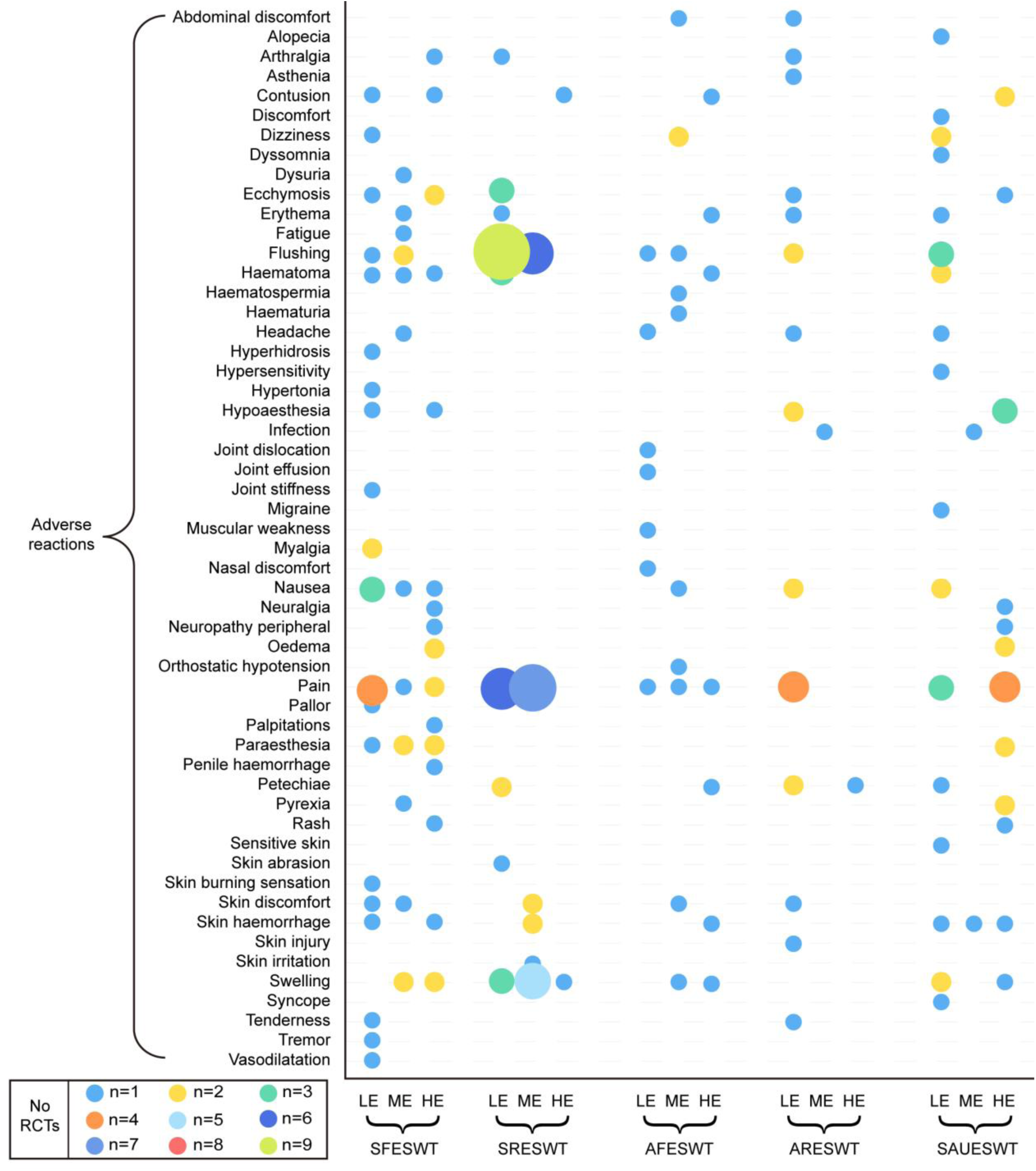
Multiaxial bubble chart of the number of randomised controlled trials reporting specific adverse reactions related to single or adjunctive extracorporeal shockwave therapy with different energy levels. AFESWT, adjunctive focused extracorporeal shockwave therapy; ARESWT, adjunctive radial extracorporeal shockwave therapy; ESWT, extracorporeal shockwave therapy; HE, high energy; LE, low energy; ME, medium energy; RCTs, randomised controlled trials; SAUESWT, single or adjunctive unknown extracorporeal shockwave therapy; SFESWT, single focused extracorporeal shockwave therapy; SRESWT, single radial extracorporeal shockwave therapy.

## Discussion

### Principal findings

We identified 210 SRs across 7 therapeutic areas and 37 diseases or conditions with high agreement from 3639 articles, 126 registrations and 21 citations, including 41649 participants from 636 RCTs. Diseases of the musculoskeletal system or connective tissue were the most frequently investigated topic and the majority of diseases or conditions were reviewed by 1 to 6 SRs, which incorporated 1 to 21 RCTs with 1 to 1000 participants (Figure 2). Most researchers assessed common clinical outcomes for diseases or conditions as positive or potentially positive that probably supported single or adjunctive ESWT as an effective therapy (Supplementary Figure S5). Type- and energy-based ESWT for diseases or conditions with potential dominant efficacy include the following: pain intensity in plantar fasciitis, erectile function in erectile dysfunction, pain intensity in lateral epicondylitis and so on (Figure 3). Type- and energy-based ESWT for diseases or conditions with potential positive efficacy include the following: global symptoms, pain intensity and quality of life in chronic prostatitis/chronic pelvic pain syndrome, pain intensity in myofascial pain syndrome, pain intensity in achilles tendinopathy and so on (Figure 4). Type- and energy-based ESWT for diseases or conditions with potential similar and adverse efficacy include the following: pain intensity in osteonecrosis of the femoral head (potential similar efficacy), pain intensity in plantar heel pain (potential similar efficacy), symptom severity in patellar tendinopathy (potential adverse efficacy) and so on (Figure 5). Following treatments of ESWT, pain, flushing, and swelling (general disorders and administration site conditions) were the most prevalent side effects and serious adverse reactions were limited (Figure 6).

### Challenges of ESWT from SRs to clinical practice

#### Lack of evidence for device settings

Large amount of the included published meta-analyses merged diverse application parameters of ESWT to assess outcomes without subgroup analysis (n=47, 32%) or conducted subgroup analyses unrelated to device settings (n=57, 39%) (Figure 7). In addition, review questions or subgroup analyses concerning types or energy levels were undertaken in small portions of SRs (n=40, 27%), and the majority of them only investigated one side (n=33, 83%) or two sides without integration (n=4, 10%) (Figure 7). In the remaining 3 SRs, knee tendinopathies and other soft tissue disorders, lower extremity tendinopathy, and calcifying tendinitis of the shoulder intervened by type- and energy-based ESWT were measured [53–55]. However, they did not reach consistent conclusions to ensure effectiveness of ESWT in these settings in comparison to other interventions. Furthermore, the measurement of impulse frequency and number of shocks involved limited diseases or conditions (Figure 7), where merely 3 SRs illustrated constructive results for clinical practice [56–58]. Evidence above indicated that even though common indications were evaluated by massive meta-analyses (Figure 2), evidence of physical treatment parameters is limited. Hence, physiatrists probably receive restricted guidance from SRs to determine primary device settings of ESWT in clinical practice.

**Figure 7.**
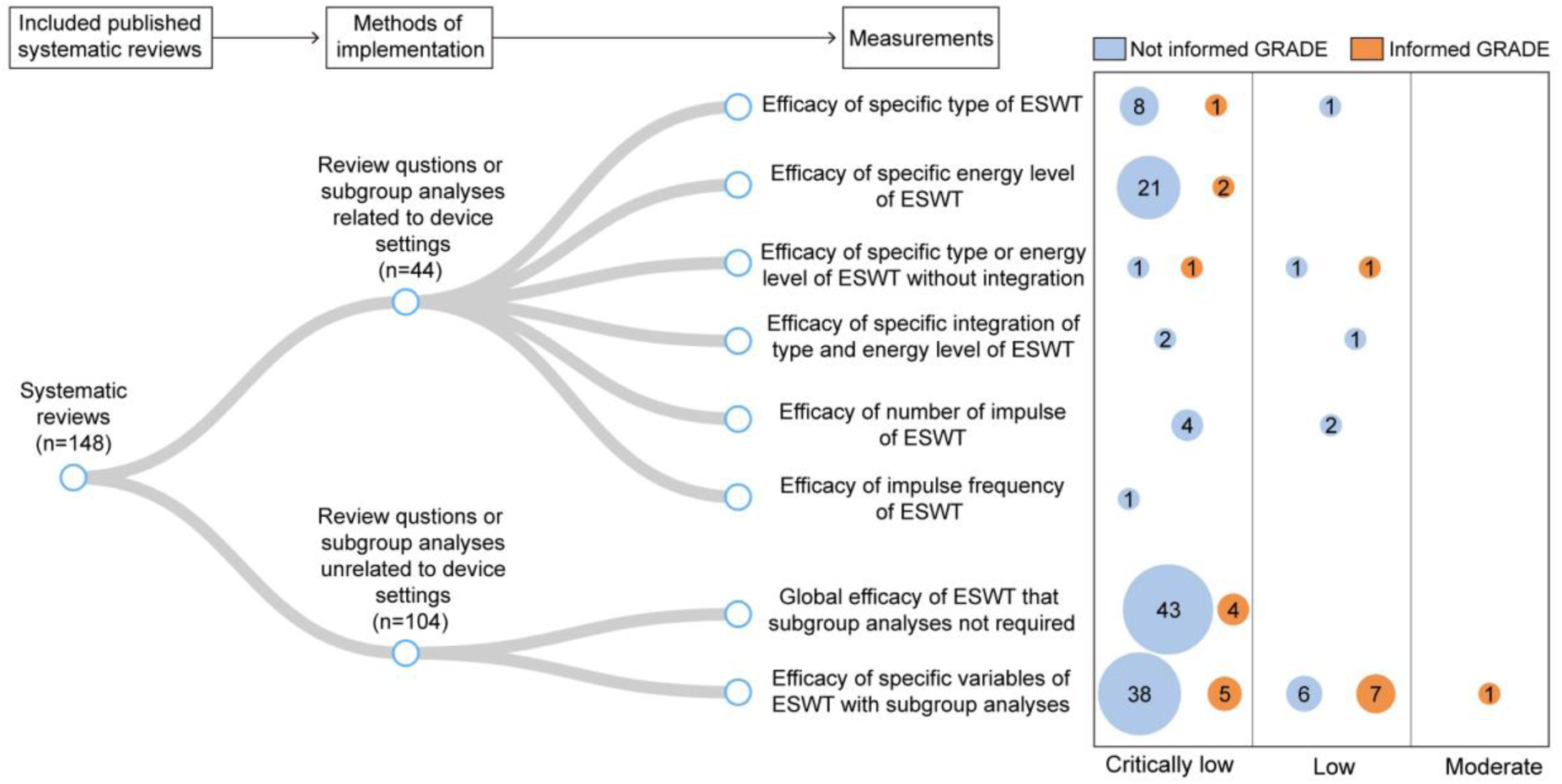
Distribution of measurements from methods of implementation in published systematic reviews corresponding to methodological quality and GRADE usage. ESWT, extracorporeal shockwave therapy; GRADE, Grading of Recommendations Assessment, Development and Evaluation.

#### Lack of credibility for decision-making

An extremely large amount of published SRs were classified as low and critically low in methodological quality (n=147, 99%) (Figure 7) that they may be insufficient to reflect the current standards for evidence synthesis and to offer accurate evidence about the literature [44]. Moreover, many published SRs did not assess the quality of outcomes by GRADE (n=126, 85%) (Figure 7). The deficiency of certainty in evidence may make recommendations weak although physiatrists realized the efficacy of ESWT on specific outcomes for diseases or conditions from SRs [59]. Influenced by these SRs with critical methodological flaws and evidence of inconclusive qualities, physiatrists might reach incorrect conclusions on the application of ESWT, resulting in negative impacts on decision-making reliability [60,61].

### Strengths and limitations of study

According to our knowledge, this overview is the first study synthesizing and visualizing a comprehensive summary of the highest level evidence on ESWT for diseases or conditions. Given the growing demand for rehabilitation worldwide, especially for musculoskeletal disorders, ESWT, as a cost- and time-efficient physiotherapy, is available to assertively benefit populations globally [62–64]. Hence, identifying the association among effectiveness, safety, and type- and energy-based ESWT has crucial clinical significance for developing application strategies against various disorders according to worldwide needs. Concentrating on RCTs, our study was conducted following systematic procedures with efforts to minimize bias. Two independent authors comprehensively searched SRs and related up-to-date RCTs with an assessment of study selection consistency to ensure our study’s reliability and timeliness. Data extraction and evaluation of quality (methods and evidence) were performed in independent and duplicate process by two groups of reviewers. After cross-checking the results, we re-estimated the magnitude of summary effect on RR or SMD with 95% CI against type- and energy-based ESWT application in subgroup analyses. We supplemented concrete interventions of control group for each outcome related to clinical practice. Generalized ADRs linked to single or adjunctive type- and energy-based ESWT treatments were reported. Appraisal of methodological quality (AMSTAR 2), certainty (GRADE), and treatment status (classification criteria) were applied to determine our confidence in the results. Furthermore, through a combination of umbrella review and evidence mapping approach, our study offers readers a broad perspective of the existing evidence landscape. The visual maps also assist readers in rapidly recognizing the clinical application gap of ESWT as well as future research needs in a user-friendly way.

Our study also has several limitations. First, we only included diseases or conditions that were investigated or remain under investigation by SRs and up-to-date RCTs. This drawback affected the comprehensiveness of relevant indications in our analysis, leading to distinction from consensus results [14,15], especially for bone pathologies. Excluding observational studies might also limit our study’s findings. Moreover, we integrated different variables (e.g. follow-up periods, number of impulses, and local anesthesia usage) to re-estimate the efficacy and safety of ESWT, resulting in substantial heterogeneity. Researchers may adjust device settings or movements over time in RCTs, yet our study simply carried out qualitative analysis. We could not demonstrate the severity of reported ADRs due to unrecorded number of participants. Another issue is that our research is saturated with small-study effects. Even though we included 636 RCTs, the number of studies included in the outcomes that corresponded to comparisons was relatively limited owing to the classification of subgroup analysis. This problem rendered our study incapable of detecting precise heterogeneity since both Cochran’s Q test and I-square statistic were biased [65,66]. As a result, due to exacerbated biases, we could not investigate heterogeneity and publication bias (at least 10 RCTs). A large number of SRs with low or critically low methodological quality aggravated our study’s limitations at the same time. Finally, we concluded that current evidence is not robust enough to draw a firm conclusion regarding the optimal type- and energy-based ESWT for specific diseases or conditions in light of aforementioned limitations. Thus, we provided type- and energy-based ESWT candidates against evidence classification criteria that require caution in interpreting our research.

### Comparison with other study

Currently, there was only one overview of SRs on ESWT [67]. Yuan et al. used the AMSTAR 2, the Risk of Bias in Systematic Reviews (ROBIS), PRISMA, and GRADE to review 8 SRs on low-intensity ESWT for erectile dysfunction. Their objective was to summarize the current clinical effectiveness evidence rather than presenting a comprehensive and systematic landscape of ESWT for diseases or conditions.

### Implications for clinical practice

Shared decision making is a collaborative approach between clinicians and patients that relies on evidence-based information [68]. In the clinical model, pertinent options from physiatrists are priorly provided to patients for determining a course of action according to their preferences [69]. Nevertheless, physiatrists and patients may confront problems in obtaining timely and comprehensive access and interpretation of the effectiveness of specific comparisons for diseases or conditions. This gap is what our findings can fill with abundant available evidence for ESWT. First, our subgroup analyses provided large-scale outcomes from comparisons between type- and energy-based ESWT and other interventions (Supplementary Figure S8, 9, 10 and 11). And we also supplemented concrete interventions of control group and visualized the evidence in a user-friendly way. These actions can directly assist physiatrists and patients in ascertaining the potential effectiveness of transparent candidates, increasing confidence in making choices, and facilitating problem-solving in communications, if shared decision making is needed. Safety of ESWT contributes to this process as well (Figure 6), since we reported generalized ADRs corresponding to single or adjunctive type- and energy-based ESWT treatments. This summary can address the needs of physiatrists and patients with respect to rapid and standard evidence identification [70–72].

### Implications for research and support

Four treatment statuses regarding effectiveness of type- and energy-based ESWT were identified (Figure 3, 4 and 5). Currently, these areas are not firmly determined and may reveal promising targets for future clinical trials. Trialists can review our findings before conducting new research to present high-level evidence with well-designed methods. Meanwhile, the knowledge gaps (evidence deficiency for device settings and credibility deficiency for decision-making) between SRs and real-world practice necessitate special attention. Our study plainly compared the external effectiveness of ESWT, yet its relevant internal comparisons for optimal type and energy involve limited diseases or conditions [73–79]. Beyond bone conditions, the dose-response effects of ESWT for various domains remain unclear [23]. Thus, more penetrating and conclusive head-to-head research is preferentially required for treatment parameters of ESWT. Additionally, our study provided the re-estimated summary effect of only energy-based ESWT (Supplementary Figure S12). These fragmentary results were derived from primary RCTs with the absence of detailed written protocols or documentation of their equipment usage. Future studies should offer multidimensional and precise information on shockwave parameters and protocols.

Policy-makers and funding agencies can utilize our evidence summaries to identify the priorities of need and relevance for ESWT research opportunities, ensuring that valuable resources are not wasted.

## Conclusion

In conclusion, this umbrella review and evidence mapping support the potential positive effectiveness and safety of varied type- and energy-based ESWT for most diseases or conditions, whether as monotherapy or combination therapy. It is noteworthy that the majority of existing SRs of ESWT probably neglect high-quality clinical guidance for device settings and decision-making (evidence deficiency and credibility deficiency). Evidence for the optimal treatment parameters on ESWT for specific diseases or conditions remains uncertain. Consequently, with the assistance of implications we provided, future research is urgently warranted to conduct more large-scale and well-designed studies in real-world practice for obtaining conclusive evidence as well as filling the knowledge gap.

## Declarations

### Ethics Approval and Consent to Participate

Not applicable.

### Availability of Data and Materials

All data in this study are collected from published articles; Materials are available in an online repository (Mendeley Data, v1; 2024. https://doi.org/10.17632/8jcbx3hzrv.1); Detailed datasets will be available upon reasonable request by emailing X.Y.Zheng.

## Acknowledgements

We gratefully acknowledge the rehabilitation therapists from Nanfang Hospital and Zhujiang Hospital, both affiliated to Southern Medical University, for putting forward their insights for extracorporeal shockwave therapy. We thank Liming Lu from Guangzhou University of Chinese Medicine for his assistance in preparing this manuscript.

## Author contributions

MQC and ZJL are joint first authors and contributed equally to this work. TCZ, ZJH and ZXY are joint corresponding authors and contributed equally to this work. MQC, ZJL, TCZ, ZJH and ZXY conceptualized the study. ZJL, HFF, GP, WT, WSM and ZJQ contributed to the method design and data extraction. MQC, ZJL, HFF and ZXY analyzed the results, created visual graphics, and drafted the manuscript. All the authors have read, revised, and approved the final manuscript.

