## Supplementary Figure S2 for "Effectiveness and Safety of Type- and Energy-based Extracorporeal Shockwave Therapy in Clinical Practice: Umbrella Review and Evidence Mapping"

### Supplementary Figure S2 A list of included 210 systematic reviews

**Systematic reviews from databases,**

**Diseases of the circulatory system:**

[1] Tsai YL, I TJ, Chuang YC, Cheng YY, Lee YC. Extracorporeal Shock Wave Therapy Combined with Complex Decongestive Therapy in Patients with Breast Cancer-Related Lymphedema: A Systemic Review and Meta-Analysis. J Clin Med 2021;10:5970. https://doi.org/10.3390/jcm10245970.

[2] Zhang L, Weng C, Zhao Z, Fu X. Extracorporeal shock wave therapy for chronic wounds: A systematic review and meta-analysis of randomized controlled trials. Wound Repair Regen 2017;25:697-706. https://doi.org/10.1111/wrr.12566.

[3] Huang Q, Yan P, Xiong H, Shuai T, Liu J, Zhu L, et al. Extracorporeal Shock Wave Therapy for Treating Foot Ulcers in Adults With Type 1 and Type 2 Diabetes: A Systematic Review and Meta-Analysis of Randomized Controlled Trials. Can J Diabetes 2020;44:196-204.e3. https://doi.org/10.1016/j.jcjd.2019.05.006.

[4] Munir Z, Akash M, Jaiprada F, Abu Tarboush B, Ijaz O, Bseiso A, et al. Evaluation of the Effects of Extracorporeal Shockwave Therapy in Patients With Peripheral Arterial Disease: A Meta-Analysis of Randomized Control Trials. Cureus 2023;15:e34729. https://doi.org/10.7759/cureus.34729.

**Diseases of the genitourinary system:**

gy.2017.09.011.

**Diseases of the musculoskeletal system or connective tissue:**

[21] Wang YC, Huang HT, Huang PJ, Liu ZM, Shih CL. Efficacy and Safety of Extracorporeal Shockwave Therapy for Treatment of Knee Osteoarthritis: A Systematic Review and Meta-analysis. Pain Med

2020;21:822-35. https://doi.org/10.1093/pm/pnz262.

[50] Liao CD, Tsauo JY, Chen HC, Liou TH. Efficacy of Extracorporeal Shock Wave Therapy for Lower-

Limb Tendinopathy: A Meta-analysis of Randomized Controlled Trials. Am J Phys Med Rehabil 2018;97:605-19. https://doi.org/10.1097/PHM.0000000000000925.
[51] Wu T, Li S, Ren J, Wang D, Ai Y. Efficacy of extracorporeal shock waves in the treatment of myofascial pain syndrome: a systematic review and meta-analysis of controlled clinical studies. Ann Transl Med 2022;10:165. https://doi.org/10.21037/atm-22-295.
[52] Avendaño-López C, Megía-García Á, Beltran-Alacreu H, Serrano-Muñoz D, Arroyo-Fernández R, Comino-Suárez N, et al. Efficacy of Extracorporeal Shockwave therapy on pain and function in Myofascial Pain Syndrome: A systematic review and meta-analysis of randomized clinical trials. Am J Phys Med Rehabil. https://doi.org/10.1097/PHM.0000000000002286.
[53] Zhang Q, Fu C, Huang L, Xiong F, Peng L, Liang Z, et al. Efficacy of Extracorporeal Shockwave Therapy on Pain and Function in Myofascial Pain Syndrome of the Trapezius: A Systematic Review and Meta-Analysis. Arch Phys Med Rehabil 2020;101:1437-46. https://doi.org/10.1016/j.apmr.2020.02.013.
[54] Jin R, Liu X, Fang C, Yue S. Extracorporeal Shock Wave Therapy for the Treatment of myofascial pain syndrome: a meta-analysis. Chinese J. Rehabilitation Med 2017;32:1167-71. https://doi.org/10.3969/j.issn.1001-1242.2017.10.016.
[55] Su M, Zhang Q, Zhang J. The clinical effect of extracorporeal shock wave therapy on myofascial pain syndrome: a meta-analysis. Health Prote and Prom 2022;22:837-40. https://doi.org/10.3969/j.issn.1671-0223(s).2022.06.001.
[56] Jun JH, Park GY, Chae CS, Suh DC. The Effect of Extracorporeal Shock Wave Therapy on Pain Intensity and Neck Disability for Patients With Myofascial Pain Syndrome in the Neck and Shoulder: A Meta-Analysis of Randomized Controlled Trials. Am J Phys Med Rehabil 2021;100:120-9. https://doi.org/10.1097/PHM.0000000000001493.
[57] Yoo JI, Oh MK, Chun SW, Lee SU, Lee CH. The effect of focused extracorporeal shock wave therapy on myofascial pain syndrome of trapezius: A systematic review and meta-analysis. Medicine 2020;99:e19085. https://doi.org/10.1097/MD.0000000000019085.
[58] Liao CD, Tsauo JY, Chen HC, Liou TH. Efficacy of Extracorporeal Shock Wave Therapy for Lower-Limb Tendinopathy: A Meta-analysis of Randomized Controlled Trials. Am J Phys Med Rehabil 2018;97:605-19. https://doi.org/10.1097/PHM.0000000000000925.
[59] Li J, Li J, Jin X, Liu S, Zhang L. Efficacy of Extracorporeal Shockwave therapy in Osteonecrosis of the Femoral Head: A meta-analysis. Med Health 2023;1:110-5.
[60] Li J, Zhang B, Kuang G, Lu M. Meta Analysis of Clinical Effect of Shockwave Therapy for Osteonecrosis of the Femoral Head. Med Innova Chin 2023;19:159-64. https://doi.org/10.3969/j.issn.1674-4985.2022.34.038.
[61] He X, Wang W, Li W, Zhang H. Meta-analysis of Chinese Medicine Combined with Shock Wave Therapy for Early Femoral Head Necrosis. J Yunnan Univers of Chin Med 2021;44:44-51. https://doi.org/10.19288/j.cnki.issn.1000-2723.2021.03.009.
[62] Hao Y, Guo H, Xu Z, Qi H, Wang Y, Lu C, et al. Meta-analysis of the potential role of extracorporeal shockwave therapy in osteonecrosis of the femoral head. J Orthop Surg Res 2018;13:166. https://doi.org/10.1186/s13018-018-0861-7.
[63] Mei J, Pang L, Jiang Z. The effect of extracorporeal shock wave on osteonecrosis of femoral head: a systematic review and meta-analysis. Phys Sportsmed 2022;50:280-8. https://doi.org/10.1080/00913847.2021.1936685.
[64] Liao CD, Tsauo JY, Chen HC, Liou TH. Efficacy of Extracorporeal Shock Wave Therapy for Lower-

Limb Tendinopathy: A Meta-analysis of Randomized Controlled Trials. Am J Phys Med Rehabil 2018;97:605-19. https://doi.org/10.1097/PHM.0000000000000925.

[77] Lou J, Wang S, Liu S, Xing G. Effectiveness of Extracorporeal Shock Wave Therapy Witho-ut Local Anesthesia in Patients With Recalcitrant Plantar Fasciitis: A Meta-Analysis of Rando

-mized Controlled Trials. Am J Phys Med Rehabil 2017;96:529-34. https://doi.org/10.1097/PHM.0000000000000666.

[86] Chen M. Extracorporeal Shock Wave Therapy for Treatment of Rotator Cuff Tear A Meta-analysis. Zhejiang Chinese Medical University 2022.

[87] Han X, Yuan X. Extracorporeal shock wave therapy for rotator cuff tendinopathy:a meta-analysis. Chin J Evi-Bas Med 2021;21:1126-32. https://doi.org/10.7507/1672-2531.202104149.

[88] Hou C, Zhu X, Wu L, Qing W, Dong Y, Zhao Y. Effectiveness of shockwave therapy for stenosing tenosynovitis: A meta-analysis. J Hainan Med Univers 2019;25:1670-5. https://doi.org/10.13210/j.cnki.jhmu.20191009.002.

#### Diseases of the nervous system:

**Diseases of the skin:**

[1] Zhang L, Fu XB, Chen S, Zhao ZB, Schmitz C, Weng CS. Efficacy and safety of extracorporeal shock wave therapy for acute and chronic soft tissue wounds: A systematic review and meta-analysis. Int Wound J 2018;15:590-9. https://doi.org/10.1111/iwj.12902.

**Injury, poisoning or certain other consequences of external causes:**

[1] Yang Y, Kang J, Jiang T, Schmitz C, Weng C, Zhang L. Safety and efficacy of treating post-burn pathological scars with extracorporeal shock wave therapy: A meta-analysis of randomised controlled trials. Wound Repair Regen 2022;30:595-607. https://doi.org/10.1111/wrr.13037.

[2] Liao CD, Tsauo JY, Chen HC, Liou TH. Efficacy of Extracorporeal Shock Wave Therapy for Lower-Limb Tendinopathy: A Meta-analysis of Randomized Controlled Trials. Am J Phys Med Rehabil 2018;97:605-19. https://doi.org/10.1097/PHM.0000000000000925.

**Symptoms, signs or clinical findings, not elsewhere classified:**

[1] Birowo P, Rangganata E, Rasyid N, Atmoko W. Efficacy and safety of extracorporeal shockwave therapy for the treatment of chronic non-bacterial prostatitis: A systematic review and meta-analysis. PLoS One 2020;15:e0244295. https://doi.org/10.1371/journal.pone.0244295.

[2] Ge J, Zhu J. Efficacy of extracorporeal shock wave therapy on chronic prostatitis B:Meta-analysis. J Clin Pathol Res 2015;9:1662-7. https://doi.org/10.3978/j.issn.2095-6959.2015.09.017.

[3] Deng G, Wu D. Efficacy of extracorporeal shockwave therapy for chronic non-bacterial prostatitis: A

meta-analysis. Health Frie 2020;24:73.

**Systematic reviews from PROSPERO,**

[1] Yang Han, Chang Qianzhen, Zhang Hong. A comparative study of the efficacy of extracorporeal shock wave and other musculoskeletal therapies in the treatment of lumbar disc herniation: a meta-analysis of randomized controlled trials. PROSPERO 2022 CRD42022222231 Available from: https://www.crd.york.ac.uk/prospero/display_record.php?ID=CRD42022222231.

[2] Yan Chenchen, Xiong Yuan, Chen Lang, Hu Liangcong, Yori Endo, Liu Guohui, Mi Bobin. A comparative study of the efficacy of ultrasonics and extracorporeal shock wave in the treatment of tennis elbow: a meta-analysis of randomized controlled trials. PROSPERO 2019 CRD42019134467 Available from: https://www.crd.york.ac.uk/prospero/display_record.php?ID=CRD42019134467.

[3] LiTing Wang, Gwo-Chi Hu. A Comparative Study on the Therapeutic Effects of Extracorporeal Shock Wave Therapy, Therapeutic Ultrasound, Laser therapy, Short Wave Diathermy in Combination with Exercise for Knee Osteoarthritis. PROSPERO 2023 CRD42023411073 Available from: https://www.crd.york.ac.uk/prospero/display_record.php?ID=CRD42023411073.

[4] Lezheng Wang, Jian Yang. A meta-analysis of medium- and long-term efficacy of extracorpo-real shock wave therapy in the treatment of external humeral epicondylitis. PROSPERO 20-19 CRD42019138667 Available from: https://www.crd.york.ac.uk/prospero/display_record.php?ID=CRD42019138667.

[14] Lun-Xue QING, Jin YANG, Duo-Duo Li, Bin WANG, Yuan LEI, Si-Na Li, Yan-Yan Sun, Chang-Xin LIU, Xi-You WANG, Zhi-Weng WENG. Effect of the extracorporeal shock wave thera-py for knee osteoarthritis: a systematic review and meta analysis of RCTs. PROSPERO 201

8 CRD42018084749 Available from: https://www.crd.york.ac.uk/prospero/display_record.php?ID=CRD42018084749.

[26] Huan Liu, fo yang, XiaoMin Liu, Li Jiang. Efficacy and safety of extracorporeal shock wave

therapy in the treatment of radial styloid stenosing tenosynovitis: systematic review and meta analysis. PROSPERO 2020 CRD42020205267 Available from: https://www.crd.york.ac.uk/prospero/display_record.php?ID=CRD42020205267.

[49] Chaoqun Feng, Junjie Yao, Yizhou Xie, Min Zhao, Youpeng Hu, Ziang Hu, Ruoyan Li, Hao

yang Wu, Yuanxin Ge, Fei Yang, Xiaohong Fan. Small needle-knife versus extracorporeal s-hock wave for the treatment of plantar fasciitis: a systematic review with meta-analysis. PROSPERO 2023 CRD42023448813 Available from: https://www.crd.york.ac.uk/prospero/display_record.php?ID=CRD42023448813.
