## Supplementary Figure S3 for "Effectiveness and Safety of Type- and Energy-based Extracorporeal Shockwave Therapy in Clinical Practice: Umbrella Review and Evidence Mapping"

### Supplementary Figure S3 A list of excluded systematic reviews and reasons for exclusion

#### Control interventions did not meet the inclusion criteria:

#### Full text unavailable:

[8] Kong X, Hu W, Dong Z, Tian J, Wang Y, Jin C, et al. The efficacy and safety of low-intensity extracorporeal shock wave treatment combined with or without medications in Chronic prostatitis/chronic pelvic pain syndrome: a systematic review and meta-analysis. Prostate Cancer

Prostatic Dis 2023;26:483-94. https://doi.org/10.1038/s41391-022-00571-0.
[9] Luo P, Hao Y, Xu K, Lu C, Xu P. Clinical analysis of focused shock wave in the treatment of osteonecrosis of the femoral head: Meta analysis. U.S. Chin Int J Traum 2021;20:1-4.

**Not human being:**

[1] Alavi SNR, Neishaboori AM, Yousefifard M. Extracorporeal shockwave therapy in spinal cord injury, early to advance to clinical trials? A systematic review and meta-analysis on animal studies. Neuroradiol J 2021;34:552-61. https://doi.org/10.1177/19714009211026899.
[2] Daeschler SC, Harhaus L, Schoenle P, Boecker A, Kneser U, Bergmeister KD. Ultrasound and shock-wave stimulation to promote axonal regeneration following nerve surgery: a systematic review and meta-analysis of preclinical studies. Sci Rep 2018;8:3168. https://doi.org/10.1038/s41598-018-21540-5.

**Not meta-analysis:**

[1] Wang J, Wang J, Zhang K, Wang Y, Bao X. Bayesian Network Meta-Analysis of the Effectiveness of Various Interventions for Nontraumatic Osteonecrosis of the Femoral Head. Biomed Res Int 2018;2018:2790163. https://doi.org/10.1155/2018/2790163.
[2] Hsiao MY, Hung CY, Chang KV, Chien KL, Tu YK, Wang TG. Comparative effectiveness of autologous blood-derived products, shock-wave therapy and corticosteroids for treatment of plantar fasciitis: a network meta-analysis. Rheumatology (Oxford) 2015;54:1735-43. https://doi.org/10.1093/rheumatology/kev010.
[3] Hsu PC, Chang KV, Chiu YH, Wu WT, Özçakar L. Comparative Effectiveness of Botulinum Toxin Injections and Extracorporeal Shockwave Therapy for Post-Stroke Spasticity: A System-atic Review and Network Meta-Analysis. EclinicalMedicine 2021;43:101222. https://doi.org/10.1016/j.eclinm.2021.101222.
[4] Chen PC, Wu KT, Chou WY, Huang YC, Wang LY, Yang TH, et al. Comparative Effectiveness of Different Nonsurgical Treatments for Patellar Tendinopathy: A Systematic Review and Network Meta-analysis. Arthroscopy 2019;35:3117-31.e2. https://doi.org/10.1016/j.arthro.2019.06.017.
[5] Li X, Zhang L, Gu S, Sun J, Qin Z, Yue J, et al. Comparative effectiveness of extracorporeal shock wave, ultrasound, low-level laser therapy, noninvasive interactive neurostimulation, and pulsed radiofrequency treatment for treating plantar fasciitis: A systematic review and network meta-analysis. Medicine 2018;97:e12819. https://doi.org/10.1097/MD.0000000000012819.
[6] Chang KV, Chen SY, Chen WS, Tu YK, Chien KL. Comparative effectiveness of focused shock wave therapy of different intensity levels and radial shock wave therapy for treating plantar fasciitis: a systematic review and network meta-analysis. Arch Phys Med Rehabil 2012;93:1259-68. https://doi.org/10.1016/j.apmr.2012.02.023.
[7] Wu YC, Tsai WC, Tu YK, Yu TY. Comparative Effectiveness of Nonoperative Treatments for Chronic Calcific Tendinitis of the Shoulder: A Systematic Review and Network Meta-Analysi-s of Randomized Controlled Trials. Arch Phys Med Rehabil 2017;98:1678-92.e6. https://doi.org/10.1016/j.apmr.2017.02.030.
[8] Babatunde OO, Legha A, Littlewood C, et al. Comparative effectiveness of treatment options for plantar heel pain: a systematic review with network meta-analysis. Br J Sports Med 2019;53:182-94. https://doi.org/10.1136/bjsports-2017-098998.
[9] Zhang J, Zhong S, Tan T, Li J, Liu S, Cheng R, et al. Comparative Efficacy and Patient-Specific

Moderating Factors of Nonsurgical Treatment Strategies for Frozen Shoulder: An Updated Systematic Review and Network Meta-analysis. Am J Sports Med 2021;49:1669-79. https://doi.org/10.1177/0363546520956293.

[21] Liao CD, Huang YY, Chen HC, Liou TH, Lin CL, Huang SW. Relative Effect of Extracorporeal Shockwave Therapy Alone or in Combination with Noninjective Treatments on Pain and Physical Function in Knee Osteoarthritis: A Network Meta-Analysis of Randomized Controlled

Trials. Biomedicines 2022;10:306. https://doi.org/10.3390/biomedicines10020306.

[22] Kang Y, Song P, Cao D, Di X, Lu Y, Liu P, et al. The Efficacy and Safety of Extracorporeal Shockwave Therapy versus Acupuncture in the Management of Chronic Prostatitis/Chronic Pelvic Pain Syndrome: Evidence Based on a Network Meta-analysis. Am J Mens Health 2021;15:15579883211057998. https://doi.org/10.1177/15579883211057998.

[23] van der Vlist AC, Winters M, Weir A, Ardern CL, Welton NJ, Caldwell DM, et al. Which treatment is most effective for patients with Achilles tendinopathy? A living systematic review with network meta-analysis of 29 randomised controlled trials. Br J Sports Med 2021;55:249-56. https://doi.org/10.1136/bjsports-2019-101872.

[24] Xie C. A Bayesian Network Meta-Analysis Of Multiple Conservative Hip Measures For Non-Traumatic Femoral Head Necrosis. Medical College of Nanchang University 2019.

[25] Shan G, Zhao J, Zhang H, Tian Y, An L, Luo G. Network Meta-analysis comparing the efficacy of different analgesics in extracorporeal shock wave lithotripsy. Chin J Urology 2020;41:936-41. https://doi.org/10.3760/cma.j.cn112330-20190821-00375.

[26] Liu C. External treatment of traditional Chinese and western medicine in the treatment of stenosing tenosynovitis of the radial styloid: network meta-analysis. Liaoning University of Traditional Chinese Medicine 2021.

[27] Lee HY, Pyun JH, Shim SR, Kim JH. Medical Treatment for Peyronie's Disease: Systematic Review and Network Bayesian Meta-Analysis. World J Mens Health 2023. https://doi.org/10.5534/wjmh.230016.

[28] He Y, Lin Y, He X, Li C, Lu Q, He J. The conservative management for improving Visual Analog Scale (VAS) pain scoring in greater trochanteric pain syndrome: a Bayesian analysis. BMC Musculoskelet Disord 2023;24:423. https://doi.org/10.1186/s12891-023-06443-5.

**Protocol:**
