## Supplementary figures and images for "Effectiveness and Safety of Type- and Energy-based Extracorporeal Shockwave Therapy in Clinical Practice: Umbrella Review and Evidence Mapping"

### Supplementary Figure S4

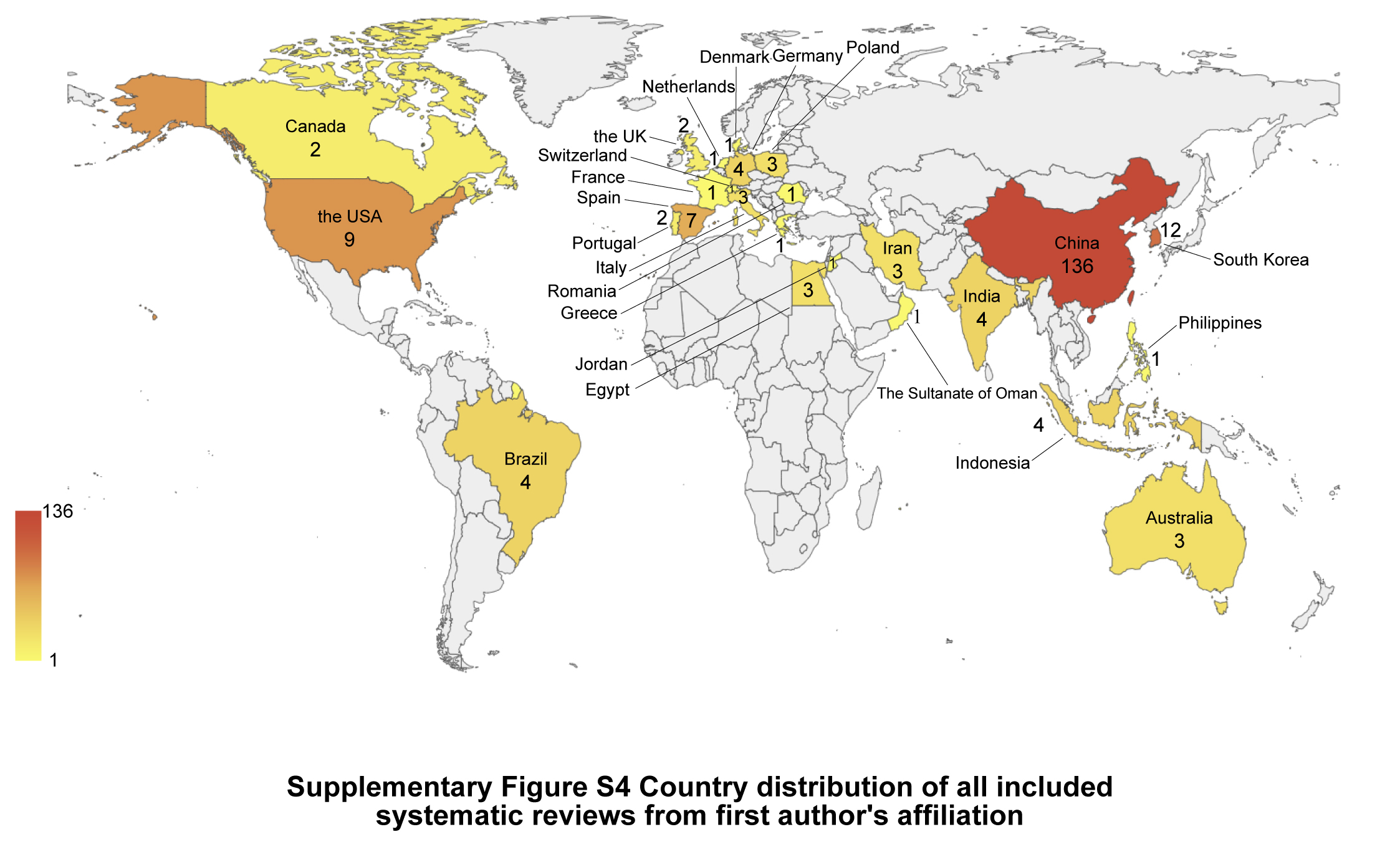

### Supplementary Figure S5

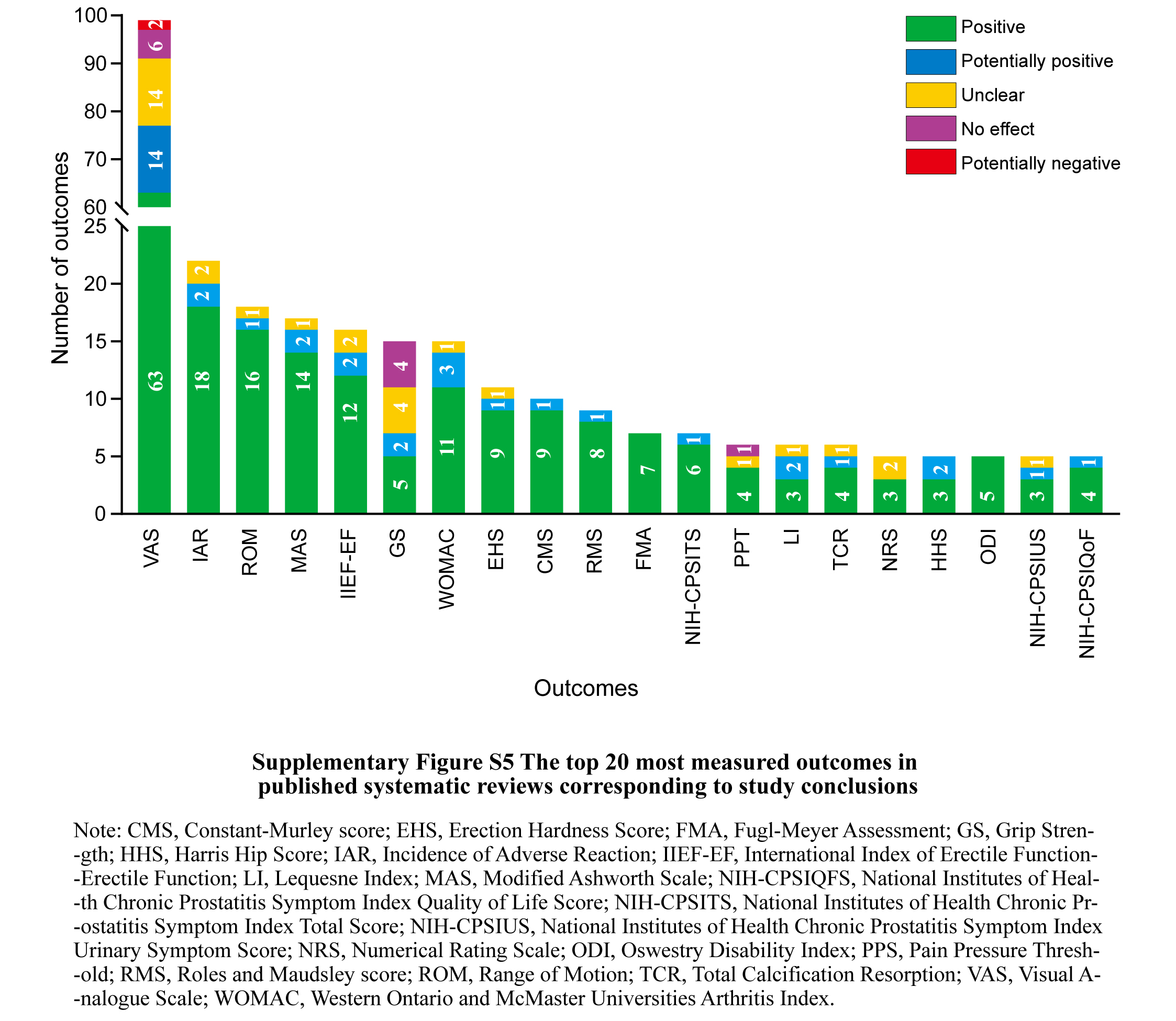

### Supplementary Figure S7

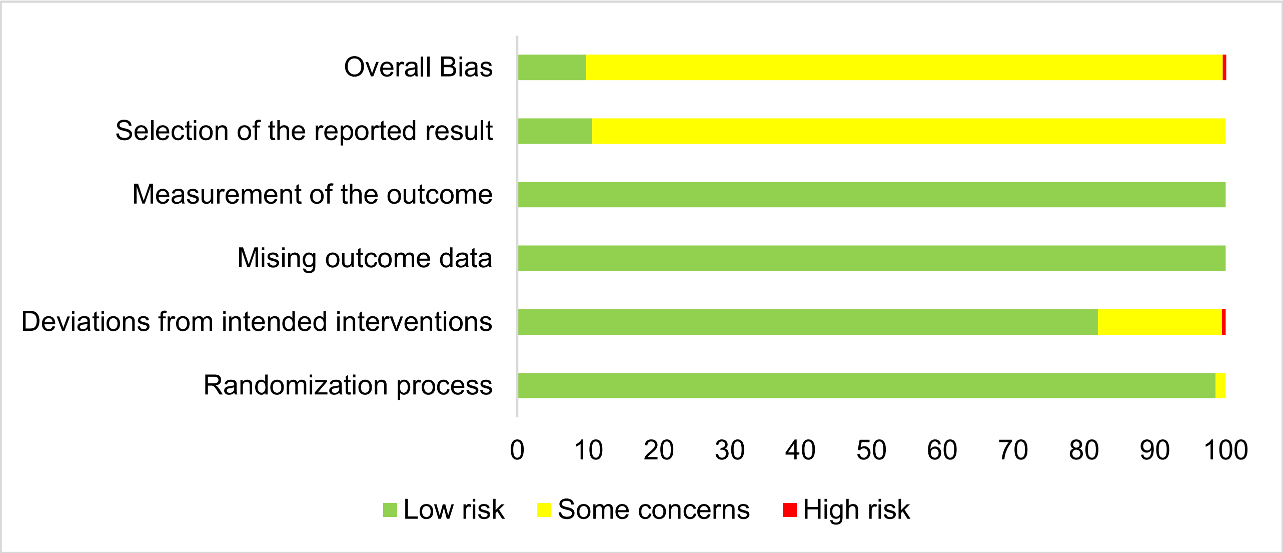


**Supplementary Figure S7 Risk of bias of up-to-date randomised controlled trials**
