## Supplementary Figure S6 for "Effectiveness and Safety of Type- and Energy-based Extracorporeal Shockwave Therapy in Clinical Practice: Umbrella Review and Evidence Mapping"

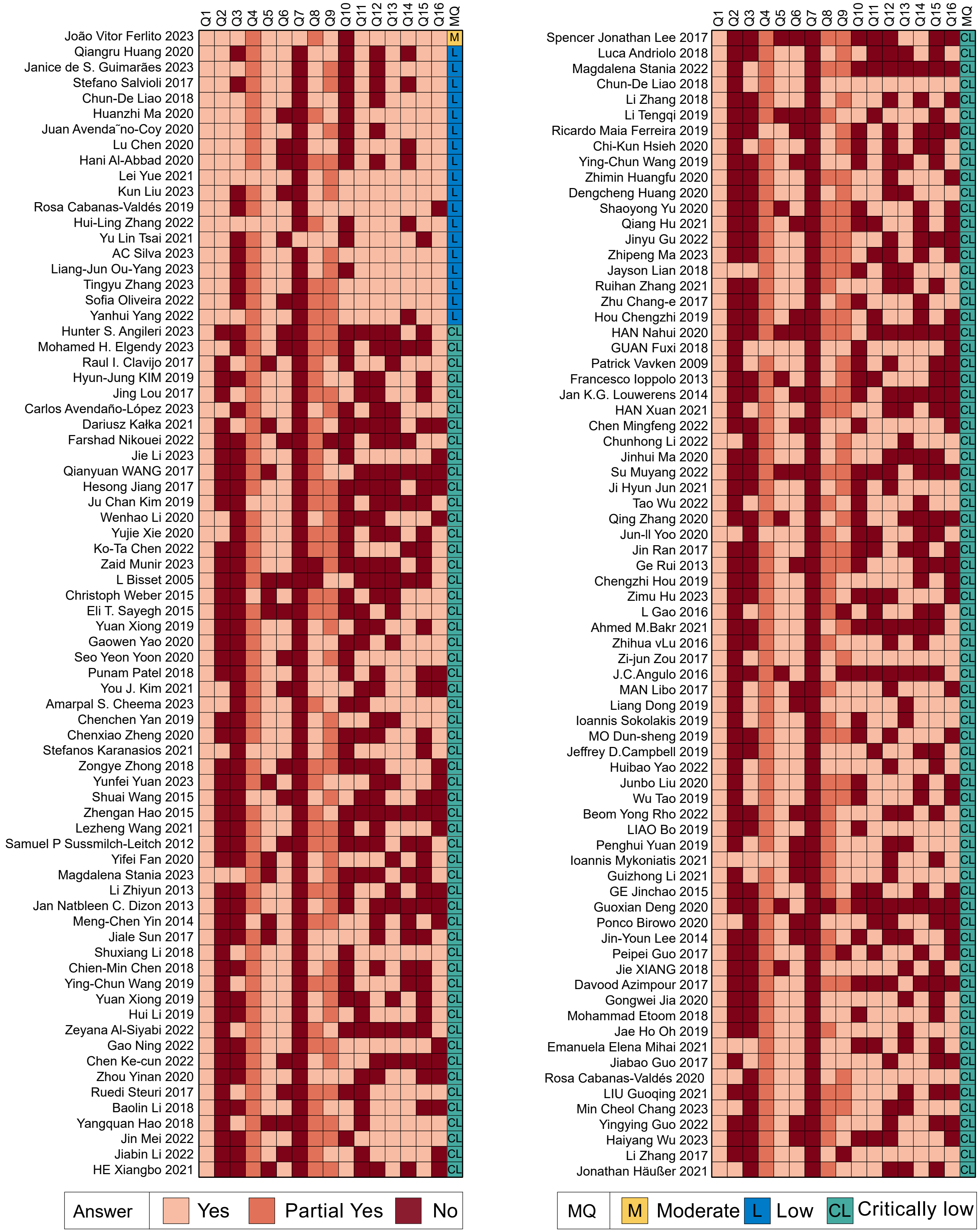


**Supplementary Figure S6 Heat map of methodological quality of the included published systematic reviews using AMSTAR 2. Questions (Q1, 2, 3, 4, 5, 6, 7, 8, 9, 10, 11, 12, 13, 14, 15, and 16) are items corresponding to original domains in AMSTAR 2.**

Note: AMSTAR 2, assessing the methodological quality of systematic review; MQ, methodological quality.
