## Supplementary Figure S8 for "Effectiveness and Safety of Type- and Energy-based Extracorporeal Shockwave Therapy in Clinical Practice: Umbrella Review and Evidence Mapping"

### Supplementary Figure S8 Distribution of single focused extracorporeal shockwave therapy evidence

| Energy level | Disease or condition | Outcome | No of RCTs | T sample | C sample | Effect estimate (95%CI) | CoE | Control group intervention |
|---|---|---|---|---|---|---|---|---|
| High energy | **Large Positive Effect with Moderate Certainty** | | | | | | | |
| | **Plantar fasciitis** | | | | | | | |
| | fESWT vs SESWT | Pain intensity (Visual Analogue Scale) | 3 | 255 | 251 | SMD=-1.88 [-3.06, -0.70] | moderate | SESWT: shockwave separated by thin foam cushion or air-chambered polyethylene foil |
| | **Large or Moderate Positive Effect with Very low Certainty** | | | | | | | |
| | **Myofascial pain syndrome** | | | | | | | |
| | fESWT vs RH | Pain intensity (Visual Analogue Scale) | 1 | 49 | 45 | SMD=-1.87 [-2.36, -1.39] | very low | RH: ultrasound therapy and transcutaneous electrical nerve stimulation |
| | fESWT vs RH | Sleep quality and quantity (Pittsburgh Sleep Quality Index) | 1 | 49 | 45 | SMD=-1.61 [-2.08, -1.14] | very low | |
| | fESWT vs RH | Depression symptom (Beck Depression Inventory) | 1 | 49 | 45 | SMD=-1.12 [-1.55, -0.68] | very low | |
| | fESWT vs RH | Function (Neck Disability Index) | 1 | 49 | 45 | SMD=-1.55 [-2.01, -1.08] | very low | |
| | **Osteonecrosis of the femoral head** | | | | | | | |
| | fESWT vs ST | Function and symptom (Harris Hip Score) | 1 | 23 | 25 | SMD=4.51 [3.41, 5.61] | very low | ST: core decompression and bone grafting |
| | fESWT vs ST | Pain intensity (Visual Analogue Scale) | 1 | 23 | 25 | SMD=-3.44 [-4.36, -2.53] | very low | |
| | **Calcific tendinitis of the shoulder** | | | | | | | |
| | fESWT vs RH | Function (Constant-Murley Score) | 1 | 96 | 84 | SMD=1.17 [0.85, 1.49] | very low | RH: transcutaneous electrical nerve stimulation |
| | fESWT vs RH | Pain intensity (Visual Analogue Scale) | 1 | 96 | 84 | SMD=-0.68 [-1.11, -0.25] | very low | |
| | **Achilles tendinopathy** | | | | | | | |
| | fESWT vs TCMET | Pain intensity (Visual Analogue Scale) | 1 | 60 | 60 | SMD=-0.83 [-1.59, -0.06] | very low | TCMET: electro-acupuncture |
| | fESWT vs SESWT | Pain intensity, function and alignment (American Orthopedic Foot and Ankle Society Score) | 1 | 44 | 42 | SMD=0.58 [0.14, 1.01] | very low | SESWT: shockwave without energy stimulation |
| | **Knee osteoarthritis** | | | | | | | |
| | fESWT vs DI | Pain intensity (Visual Analogue Scale) | 1 | 82 | 44 | SMD=-1.47 [-1.78, -1.16] | very low | DI: alprostadil |
| | fESWT vs DI | Pain intensity, stiffness and physical function (Western Ontario and McMaster Universities Arthritis Index) | 1 | 82 | 44 | SMD=-1.77 [-2.04, -1.50] | very low | |
| | **Plantar heel pain** | | | | | | | |
| | fESWT vs NT | Pain intensity (Visual Analogue Scale) | 1 | 12 | 10 | SMD=-0.52 [-0.95, -0.09] | very low | |
| | fESWT vs DI | Pain intensity (Visual Analogue Scale) | 1 | 33 | 34 | SMD=-2.12 [-2.49, -1.74] | very low | DI: triamcinolone acetonide and lidocaine |
| | fESWT vs DI | Function (Maryland Foot Score) | 1 | 33 | 34 | SMD=0.64 [0.36, 0.92] | very low | |
| | **Small or Very small Effect with Very low Certainty** | | | | | | | |
| | **Achilles tendinopathy** | | | | | | | |
| | fESWT vs SESWT | Pain intensity (Visual Analogue Scale) | 1 | 44 | 42 | SMD=-0.47 [-0.90, -0.04] | very low | SESWT: shockwave without energy stimulation |
| | **Osteonecrosis of the femoral head** | | | | | | | |
| | fESWT vs TCMIT + MT | Function and symptom (Harris Hip Score) | 1 | 75 | 78 | SMD=0.25 [0.11, 0.38] | very low | MT: alendronate; TCMIT: xianling gubao capsule |
| | fESWT vs TCMIT + MT | Pain intensity (Visual Analogue Scale) | 1 | 75 | 78 | SMD=-0.17 [-0.30, -0.04] | very low | |
| Medium energy | **Large Positive Effect with Moderate Certainty** | | | | | | | |
| | **Plantar fasciitis** | | | | | | | |
| | fESWT vs SESWT | Pain intensity (Visual Analogue Scale) | 2 | 176 | 168 | SMD=-0.55 [-0.91, -0.19] | moderate | SESWT: shockwave with sound-reflecting pad or minimal energy stimulation |

| | | | | | | | | |
|---|---|---|---|---|---|---|---|---|
| **Medium energy** | **Large or Moderate Positive Effect with Low or Very low Certainty** | | | | | | | |
| | **Rotator cuff tendinopathy** | | | | | | | |
| | fESWT vs TCMET | Pain intensity (Visual Analogue Scale) | 2 | 40 | 30 | SMD=-5.20 [-7.54, -2.87] | low | TCMET: electro-acupuncture |
| | **Erectile dysfunction** | | | | | | | |
| | fESWT vs SESWT | Function (Sexual Encounter Profile Question 3) | 1 | 30 | 30 | RR=5.50 [2.15, 14.04] | low | SESWT: shockwave without energy stimulation |
| | **Stenosing tenosynovitis** | | | | | | | |
| | fESWT vs DI | Function (Cooney Score) | 1 | 29 | 28 | SMD=0.63 [0.10, 1.16] | low | DI: triamcinolone acetonide and lidocaine |
| | **Low back pain** | | | | | | | |
| | fESWT vs SESWT | Pain intensity (Visual Analogue Scale) | 2 | 34 | 31 | SMD=-1.19 [-1.72, -0.66] | low | SESWT: shockwave separated by special polyethylene cap or of the same sound with minimal energy |
| | **Pes anserine tendinopathy** | | | | | | | |
| | fESWT + RH + MT vs SESWT + RH + MT | Pain intensity (McGill Pain Questionnaire) | 1 | 20 | 20 | SMD=-0.79 [-1.24, -0.33] | low | SESWT: shockwave of the same sound without energy stimulation; RH: stretching exercises; MT: gelofen |
| | **Chronic prostatitis/ chronic pelvic pain syndrome** | | | | | | | |
| | fESWT vs SESWT | Pain intensity, urinary symptoms, and quality of life (National Institutes of Health-Chronic Prostatitis Symptom Index Total Score) | 3 | 69 | 68 | SMD=-4.88 [-7.77, -1.98] | low | SESWT: shockwave separated by absorbing materials or without energy stimulation |
| | fESWT vs SESWT | Pain intensity (National Institutes of Health-Chronic Prostatitis Symptom Index Pain Score) | 3 | 69 | 68 | SMD=-5.84 [-9.36, -2.32] | low | |
| | fESWT vs SESWT | Symptom (National Institutes of Health-Chronic Prostatitis Symptom Index Urinary Score) | 2 | 39 | 38 | SMD=-1.32 [-1.82, -0.82] | very low | |
| | fESWT vs SESWT | Quality of life (National Institutes of Health-Chronic Prostatitis Symptom Index Quality of Life Score) | 2 | 39 | 38 | SMD=-1.87 [-2.41, -1.32] | very low | |
| | fESWT vs RH | Pain intensity, urinary symptoms, and quality of life (National Institutes of Health-Chronic Prostatitis Symptom Index Total Score) | 2 | 119 | 95 | SMD=-1.13 [-1.42, -0.84] | very low | RH: high-frequency hyperthermia |
| | fESWT vs RH | Pain intensity (National Institutes of Health-Chronic Prostatitis Symptom Index Pain Score) | 2 | 119 | 95 | SMD=-0.96 [-1.25, -0.68] | very low | |
| | fESWT vs RH | Quality of life (National Institutes of Health-Chronic Prostatitis Symptom Index Quality of Life Score) | 2 | 119 | 95 | SMD=-0.99 [-1.27, -0.70] | very low | |
| | **Lateral epicondylitis** | | | | | | | |
| | fESWT vs RH | Pain intensity (Visual Analogue Scale) | 1 | 40 | 40 | SMD=-0.93 [-1.39, -0.47] | low | RH: ultrasound therapy and cryotherapy |
| | fESWT vs TCMET | Pain intensity (Visual Analogue Scale) | 1 | 20 | 20 | SMD=-2.95 [-3.87, -2.03] | very low | TCMET: electro-acupuncture |
| | **Osteonecrosis of the femoral head** | | | | | | | |
| | fESWT + TCMIT vs TCMIT | Function and symptom (Harris Hip Score) | 1 | 32 | 25 | SMD=2.90 [2.14, 3.66] | very low | TCMIT: taohongsiwu decoction |
| | **Achilles tendinopathy** | | | | | | | |
| | fESWT vs SESWT | Function (Lower Limb Functional Index) | 1 | 22 | 27 | SMD=1.05 [0.45, 1.65] | very low | SESWT: shockwave separated by opaque cloth |
| | **Patellar tendinopathy** | | | | | | | |
| | fESWT vs RH + MT | Pain intensity (Visual Analogue Scale) | 1 | 30 | 24 | SMD=-3.47 [-4.34, -2.60] | very low | MT: nonsteroidal anti-inflammatory drugs; RH: physiotherapy, exercise program, knee strap and modification of activity levels |
| | fESWT vs RH + MT | Symptom (Victorian Institute of Sport Assessment-Patella Questionnaire) | 1 | 30 | 24 | SMD=4.77 [3.69, 5.85] | very low | |
| | fESWT vs DI | Pain intensity (Visual Analogue Scale) | 1 | 69 | 69 | SMD=-0.62 [-0.96, -0.28] | very low | DI: platelet-rich plasma |
| | **Frozen shoulder** | | | | | | | |
| | fESWT vs TCMET | Function (Constant-Murley Score) | 1 | 39 | 39 | SMD=2.00 [1.45, 2.55] | very low | TCMET: tuina |
| | fESWT vs RH | Pain intensity (Visual Analogue Scale) | 1 | 47 | 47 | SMD=-1.24 [-1.68, -0.80] | very low | RH: medium-frequency electrotherapy and ultrasound therapy |
| | **Small Effect with Low Certainty** | | | | | | | |
| | **Low back pain** | | | | | | | |
| | fESWT vs SESWT | Pain intensity (Laitinen Pain Scale) | 1 | 20 | 20 | SMD=-0.49 [-0.85, -0.13] | low | SESWT: shockwave separated by special polyethylene cap |

| | | | | | | | | |
|---|---|---|---|---|---|---|---|---|
| **Medium energy** | **Large or Moderate Negative Effect with Low or Very low Certainty** | | | | | | | |
| | **Plantar heel pain** | | | | | | | |
| | fESWT vs ST | Pain intensity (Visual Analogue Scale) | 1 | 11 | 14 | SMD=1.41 [0.51, 2.31] | low | ST: endoscopic plantar fasciotomy |
| | fESWT vs ST | Pain intensity and function (Roles and maudsley score) | 1 | 11 | 14 | SMD=1.53 [0.61, 2.44] | low | |
| | **Patellar tendinopathy** | | | | | | | |
| | fESWT vs DI | Symptom (Victorian Institute of Sport Assessment-Patella Questionnaire) | 1 | 69 | 69 | SMD=-0.61 [-0.97, -0.26] | very low | DI: platelet-rich plasma |
| | **Carpal tunnel syndrome** | | | | | | | |
| | fESWT vs DI | Pain intensity (Visual Analogue Scale) | 2 | 35 | 36 | SMD=0.52 [0.04, 0.99] | very low | DI: triamcinolone acetonide or mecobalamin |
| **Low energy** | **Large or Moderate Positive Effect with Moderate Certainty** | | | | | | | |
| | **Erectile dysfunction** | | | | | | | |
| | fESWT vs SESWT | Function(International Index of Erectile Function-Erectile Function Domain Score) | 14 | 505 | 404 | SMD=0.77 [0.43, 1.10] | moderate | SESWT: shockwave separated by absorbing materials (gel pad, cap, mental plate) or without energy stimulation |
| | fESWT vs SESWT | Funtion(Erection Hardness Score >= 3 rate) | 7 | 330 | 241 | RR=2.55 [1.45, 4.47] | moderate | |
| | **Lateral epicondylitis** | | | | | | | |
| | fESWT vs SESWT | Pain intensity (Visual Analogue Scale) | 5 | 196 | 197 | SMD=-0.67 [-1.16, -0.18] | moderate | SESWT: shockwave without energy stimulation |
| | **Large or Moderate Positive Effect with Low or Very low Certainty** | | | | | | | |
| | **Carpal tunnel syndrome** | | | | | | | |
| | fESWT vs RH | Funtion and symptom (Boston Carpal Tunnel Syndrome Questionnaire) | 1 | 16 | 9 | SMD=-0.72 [-1.21, -0.24] | low | RH: ultrasound therapy |
| | **Rotator cuff tendinopathy** | | | | | | | |
| | fESWT vs SESWT | Function (Constant-Murley Score) | 1 | 22 | 18 | SMD=1.15 [0.46, 1.83] | low | SESWT: shockwave of the same sound without energy stimulation |
| | **Chronic prostatitis/ chronic pelvic pain syndrome** | | | | | | | |
| | fESWT vs SESWT | Pain intensity, urinary symptoms, and quality of life (National Institutes of Health-Chronic Prostatitis Symptom Index Total Score) | 5 | 158 | 134 | SMD=-1.20 [-1.61, -0.79] | low | SESWT: shockwave of the same sound without energy stimulation |
| | fESWT vs SESWT | Pain intensity (National Institutes of Health-Chronic Prostatitis Symptom Index Pain Score) | 4 | 126 | 123 | SMD=-0.99 [-1.23, -0.75] | low | |
| | fESWT vs SESWT | Quality of life (National Institutes of Health-Chronic Prostatitis Symptom Index Quality of Life Score) | 4 | 126 | 123 | SMD=-1.04 [-1.28, -0.80] | low | |
| | fESWT vs SESWT | Pain intensity (Visual Analogue Scale) | 2 | 28 | 26 | SMD=-1.32 [-1.74, -0.90] | low | |
| | fESWT vs RH | Pain intensity, urinary symptoms, and quality of life (National Institutes of Health-Chronic Prostatitis Symptom Index Total Score) | 4 | 123 | 115 | SMD=-2.15 [-3.31, -0.98] | very low | RH: high-frequency hyperthermia |
| | fESWT vs RH | Pain intensity (National Institutes of Health-Chronic Prostatitis Symptom Index Pain Score) | 4 | 123 | 115 | SMD=-2.05 [-3.29, -0.82] | very low | |
| | **Lateral epicondylitis** | | | | | | | |
| | fESWT vs SESWT | Funtion and symptom (Patient-Rated Tennis Elbow Evaluation Questionnaire) | 1 | 30 | 28 | SMD=1.64 [1.22, 2.07] | low | SESWT: shockwave with minimal energy stimulation |
| | fESWT vs DI | Pain intensity (Visual Analogue Scale) | 1 | 24 | 24 | SMD=-1.28 [-1.90, -0.65] | very low | DI: triamcinolone acetonide and lidocaine |
| | **Calcific tendinitis of the shoulder** | | | | | | | |
| | fESWT vs SESWT | Pain intensity (Visual Analogue Scale) | 1 | 20 | 18 | SMD=-3.00 [-3.43, -2.57] | low | SESWT: shockwave without energy stimulation |
| | fESWT vs SESWT | Function (Constant-Murley Score) | 1 | 40 | 40 | SMD=1.40 [0.91, 1.90] | very low | |
| | **Cerebral palsy spasticity** | | | | | | | |
| | fESWT vs SESWT | Symptom (Modified Ashworth Scale) | 1 | 12 | 12 | SMD=-2.56 [-6.39, -1.44] | very low | SESWT: shockwave of the same sound without energy stimulation |
| | fESWT vs SESWT | Funtion (Range of Motion) | 1 | 12 | 12 | SMD=4.19 [2.67, 5.71] | very low | |
| | fESWT vs SESWT | Structure (Plantar Surface Area) | 1 | 12 | 12 | SMD=6.43 [4.28, 8.57] | very low | |

| | | | | | | | | |
|---|---|---|---|---|---|---|---|---|
| Low energy | **Post-stroke upper limb spasticity** | | | | | | | |
| | fESWT vs SESWT | Funtion (Range of Motion) | 1 | 20 | 20 | SMD=1.35 [0.36, 2.35] | very low | SESWT: shockwave of the same sound without energy stimulation |
| | **Myofascial pain syndrome** | | | | | | | |
| | fESWT vs SESWT | Pain intensity (Visual Analogue Scale) | 2 | 18 | 22 | SMD=-0.99 [-1.66, -0.32] | very low | SESWT: shockwave of the same sound without energy stimulation |
| | fESWT vs SESWT | Pain intensity (Pain Pressure Threshold) | 2 | 18 | 22 | SMD=1.43 [0.71, 2.14] | very low | |
| | fESWT vs RH + DI | Funtion (Range of Motion) | 1 | 30 | 30 | SMD=1.05 [0.38, 1.72] | very low | RH: transcutaneous electrical nerve stimulation; DI: not inform |
| | fESWT vs DI | Pain intensity (Visual Analogue Scale) | 1 | 30 | 30 | SMD=-1.51 [-2.09, -0.92] | very low | |
| | fESWT vs DI | Pain intensity (Pain Pressure Threshold) | 1 | 30 | 30 | SMD=1.17 [0.62, 1.73] | very low | DI: trigger point injections |
| | fESWT vs DI | Funtion (Quebec Back Pain Disability Scale) | 1 | 30 | 30 | SMD=-0.66 [-1.18, -0.14] | very low | |
| | **Plantar fasciitis** | | | | | | | |
| | fESWT vs DI | Pain intensity (Visual Analogue Scale) | 1 | 16 | 16 | SMD=-0.78 [-1.50, -0.06] | very low | DI: methylprednisolone and mepivacaine |
| | **Stenosing tenosynovitis** | | | | | | | |
| | fESWT vs DI | Function (Cooney Score) | 1 | 30 | 28 | SMD=1.21 [0.65, 1.78] | very low | DI: triamcinolone acetonide and lidocaine |
| | fESWT vs SESWT | Pain intensity (Visual Analogue Scale) | 1 | 20 | 20 | SMD=-0.76 [-1.13, -0.38] | very low | SESWT: shockwave separated by a specific applicator |
| | fESWT vs SESWT | Funtion and symptom (Disabilities of the Arm, Shoulder, and Hand Questionnatire) | 1 | 20 | 20 | SMD=-0.52 [-0.99, -0.04] | very low | |
| | **Small Effect with Low or Very low Certainty** | | | | | | | |
| | **Rotator cuff tendinopathy** | | | | | | | |
| | fESWT vs SESWT | Funtion and pain intensity (Shoulder Pain and Disability Index) | 1 | 34 | 40 | SMD=-0.29 [-0.52, -0.06] | low | SESWT: shockwave of the same sound without energy stimulation |
| | **Achilles tendinopathy** | | | | | | | |
| | fESWT vs RH | Pain intensity, function and alignment (American Orthopedic Foot and Ankle Society Score) | 1 | 30 | 30 | SMD=-0.42 [-0.78, -0.06] | very low | RH: cold air therapy and high power laser therapy |
| | **Large Negative Effect with Very low Certainty** | | | | | | | |
| | **Achilles tendinopathy** | | | | | | | |
| | fESWT vs RH | Pain intensity (Visual Analogue Scale) | 1 | 30 | 30 | SMD=1.56 [0.63, 2.48] | very low | RH: cold air therapy and high power laser therapy |

Note: C, control group; CI, confidence interval; CoE, certainty of evidence assessed using the Grading of Recommendations Assessment, Development and Evaluation approach; DI, drug injection; fESWT, focused extracorporeal shockwave therapy; MT, medication therapy; NT, no treatment; RCTs, randomised controlled trials; RH, rehabilitation; RR, relative risk; SESWT, sham extracorporeal shockwave therapy; SMD, standardized mean difference; ST, surgical treatment; T, treatment group; TCMET, traditional Chinese medicine external therapy; TCMIT, traditional Chinese medicine internal therapy.
