## Supplementary Figure S9 for "Effectiveness and Safety of Type- and Energy-based Extracorporeal Shockwave Therapy in Clinical Practice: Umbrella Review and Evidence Mapping"

### Supplementary Figure S9 Distribution of single radial extracorporeal shockwave therapy evidence

| Energy level | Disease or condition | Outcome | No of RCTs | T sample | C sample | Effect estimate (95%CI) | CoE | Control group intervention |
|---|---|---|---|---|---|---|---|---|
| High energy | **Large or Moderate Positive Effect with Low Certainty** | | | | | | | |
| | **Post-stroke upper limb spasticity** | | | | | | | |
| | rESWT vs SESWT | Pain intensity (Visual Analogue Scale) | 1 | 17 | 17 | SMD=-0.91 [-1.63, -0.20] | low | SESWT: shockwave of the same sound without energy stimulation |
| | **Frozen shoulder** | | | | | | | |
| | rESWT vs RH | Pain intensity (Visual Analogue Scale) | 1 | 34 | 34 | SMD=-1.65 [-2.20, -1.09] | low | RH: medium-frequency electrotherapy |
| | **Myofascial pain syndrome** | | | | | | | |
| | rESWT vs RH | Pain intensity (Visual Analogue Scale) | 1 | 22 | 23 | SMD=-0.73 [-1.34, -0.13] | low | RH: ultrasound therapy and hot pack therapy |
| | rESWT vs RH | Pain intensity (Pain Pressure Threshold) | 1 | 22 | 23 | SMD=0.79 [0.18, 1.40] | low | |
| | **Knee osteoarthritis** | | | | | | | |
| | rESWT vs SESWT | Pain intensity (Visual Analogue Scale) | 1 | 20 | 40 | SMD=-2.25 [-2.93, -1.57] | low | SESWT: shockwave of the same sound without energy stimulation |
| | rESWT vs DI | Pain intensity (Visual Analogue Scale) | 1 | 20 | 40 | SMD=-1.72 [-2.35, -1.10] | low | DI: corticosteroids |
| | **Lateral epicondylitis** | | | | | | | |
| | rESWT vs RH | Pain intensity (Visual Analogue Scale) | 3 | 75 | 78 | SMD=-2.42 [-4.47, -0.37] | low | RH: ultrasound therapy or kinesiology taping |
| | **Plantar fasciitis** | | | | | | | |
| | rESWT vs RH + MT | Pain intensity (Visual Analogue Scale) | 1 | 30 | 30 | SMD=-1.54 [-2.12, -0.95] | low | RH: stretching exercises; MT: diclofenac |
| Medium energy | **Large or Moderate Positive Effect with Moderate Certainty** | | | | | | | |
| | **Plantar fasciitis** | | | | | | | |
| | rESWT vs RH | Pain intensity (Visual Analogue Scale) | 9 | 315 | 293 | SMD=-0.70 [-1.31, -0.08] | moderate | RH: ultrasound therapy or kinesiology taping or foot orthotics or laser therapy |
| | **Knee osteoarthritis** | | | | | | | |
| | rESWT vs SESWT | Pain intensity (Visual Analogue Scale) | 4 | 175 | 181 | SMD=-1.68 [-2.01, -1.36] | moderate | SESWT: shockwave of the same sound without energy stimulation |
| | rESWT vs SESWT | Pain intensity, stiffness and physical function (Western Ontario and McMaster Universities Arthritis Index) | 4 | 175 | 181 | SMD=-1.15 [-1.61, -0.69] | moderate | |
| | rESWT vs SESWT | Symptom (Lequesne Index) | 4 | 175 | 181 | SMD=-1.05 [-1.57, -0.54] | moderate | |
| | rESWT vs MT | Pain intensity (Visual Analogue Scale) | 4 | 201 | 200 | SMD=-1.63 [-2.32, -0.93] | moderate | MT: etoricoxib or celecoxib |
| | rESWT vs MT | Pain intensity, stiffness and physical function (Western Ontario and McMaster Universities Arthritis Index) | 3 | 174 | 174 | SMD=-2.43 [-4.16, -0.70] | moderate | |
| | **Large or Moderate Positive Effect with Low or Very low Certainty** | | | | | | | |
| | **Proximal hamstring tendinopathy** | | | | | | | |
| | rESWT vs RH + MT | Pain intensity (Visual Analogue Scale) | 1 | 20 | 20 | SMD=-2.65 [-3.54, -1.75] | low | RH: physiotherapy and a exercise program; MT: nonsteroidal anti-inflammatory drugs |
| | rESWT vs RH + MT | Pain intensity (Nirschl Phase Rating Scale) | 1 | 20 | 20 | SMD=-2.70 [-3.21, -2.19] | low | |
| | **Myofascial pain syndrome** | | | | | | | |
| | rESWT vs RH | Pain intensity (Visual Analogue Scale) | 1 | 22 | 23 | SMD=-1.71 [-2.41, -1.02] | low | RH: ultrasound therapy |
| | rESWT vs TCMET | Pain intensity (Visual Analogue Scale) | 4 | 137 | 133 | SMD=-0.70 [-1.06, -0.34] | low | TCMET: dry needling or tuina or electro-acupuncture |
| | **Low back pain** | | | | | | | |
| | rESWT vs SESWT | Pain intensity (Visual Analogue Scale) | 2 | 78 | 66 | SMD=-1.85 [-2.82, -0.87] | low | SESWT: shockwave of the same sound without energy stimulation |
| | rESWT vs SESWT | Function (Oswestry Disability Index) | 2 | 78 | 66 | SMD=-2.67 [-4.97, -0.37] | low | |

| | | | | | | | | |
|---|---|---|---|---|---|---|---|---|
| Medium energy | **Plantar fasciitis** | | | | | | | |
| | rESWT vs SESWT | Pain intensity (Visual Analogue Scale) | 2 | 94 | 97 | SMD=-13.84 [-22.56, -5.11] | low | SESWT: shockwave separated by a clasp or without energy stimulation |
| | rESWT vs RH | Function (Patient Specific Functional Scale) | 1 | 15 | 15 | SMD=-2.87 [-3.93, -1.81] | low | RH: ultrasound therapy or kinesiology taping or foot orthotics or laser therapy |
| | **Frozen shoulder** | | | | | | | |
| | rESWT vs SESWT | Pain intensity (Visual Analogue Scale) | 1 | 53 | 53 | SMD=-3.18 [-3.76, -2.60] | low | SESWT: shockwave separated by a clasp |
| | rESWT vs SESWT | Function (Range of Motion) | 1 | 53 | 53 | SMD=3.17 [2.59, 3.75] | low | |
| | rESWT vs DI | Pain intensity (Visual Analogue Scale) | 2 | 70 | 70 | SMD=-0.86 [-1.51, -0.21] | very low | DI: triamcinolone acetonide or vitamin B12 or mecobalamin or diprospan or lidocaine |
| | **Patellar tendinopathy** | | | | | | | |
| | rESWT vs RH | Pain intensity (Visual Analogue Scale) | 2 | 96 | 96 | SMD=-3.93 [-5.89, -1.97] | low | RH: manual therapy |
| | rESWT vs RH + MT | Pain intensity (Visual Analogue Scale) | 2 | 182 | 165 | SMD=-1.56 [-2.53, -0.59] | low | MT: glucosamine sulfate or celebrex; RH: manual therapy or ultrasound therapy |
| | rESWT vs RH + MT | Symptom (Victorian Institute of Sport Assessment-Patella Questionnaire) | 1 | 102 | 93 | SMD=5.25 [3.48, 7.01] | very low | |
| | **Knee osteoarthritis** | | | | | | | |
| | rESWT vs DI | Pain intensity (Visual Analogue Scale) | 3 | 173 | 173 | SMD=-0.69 [-0.91, -0.47] | low | DI: hyaluronic acid or lidocaine or betamethasone |
| | rESWT vs DI | Pain intensity, stiffness and physical function (Western Ontario and McMaster Universities Arthritis Index) | 2 | 78 | 77 | SMD=-0.79 [-1.38, -0.21] | low | |
| | rESWT vs DI | Function (Lysholm Score) | 1 | 37 | 37 | SMD=0.61 [0.14, 1.07] | very low | |
| | rESWT vs MT | Symptom (Lequesne Index) | 1 | 50 | 50 | SMD=-0.61 [-1.01, -0.21] | very low | MT: etoricoxib or celecoxib |
| | **Chronic prostatitis/ chronic pelvic pain syndrome** | | | | | | | |
| | rESWT vs SESWT | Pain intensity, urinary symptoms, and quality of life (National Institutes of Health-Chronic Prostatitis Symptom Index Total Score) | 1 | 20 | 20 | SMD=-2.89 [-4.25, -1.53] | low | SESWT: shockwave without energy stimulation |
| | rESWT vs SESWT | Pain intensity (National Institutes of Health-Chronic Prostatitis Symptom Index Pain Score) | 1 | 20 | 20 | SMD=-2.79 [-4.23, -1.34] | low | |
| | rESWT vs SESWT | Symptom (National Institutes of Health-Chronic Prostatitis Symptom Index Urinary Score) | 1 | 20 | 20 | SMD=-1.29 [-2.29, -0.30] | low | |
| | rESWT vs SESWT | Quality of life (National Institutes of Health-Chronic Prostatitis Symptom Index Quality of Life Score) | 1 | 20 | 20 | SMD=-2.08 [-2.74, -1.41] | low | |
| | rESWT vs MT | Pain intensity, urinary symptoms, and quality of life (National Institutes of Health-Chronic Prostatitis Symptom Index Total Score) | 1 | 30 | 15 | SMD=-1.28 [-1.96, -0.60] | very low | MT: tamsulosin |
| | rESWT vs MT | Pain intensity (National Institutes of Health-Chronic Prostatitis Symptom Index Pain Score) | 1 | 30 | 15 | SMD=-1.26 [-1.94, -0.59] | very low | |
| | rESWT vs MT | Symptom (National Institutes of Health-Chronic Prostatitis Symptom Index Urinary Score) | 1 | 30 | 15 | SMD=-0.69 [-1.33, -0.05] | very low | |
| | rESWT vs MT | Quality of life (National Institutes of Health-Chronic Prostatitis Symptom Index Quality of Life Score) | 1 | 30 | 15 | SMD=-0.88 [-1.53, -0.23] | very low | |
| | rESWT vs MT | Pain intensity (Visual Analogue Scale) | 1 | 30 | 15 | SMD=-1.13 [-1.80, -0.47] | very low | |
| | rESWT vs RH | Pain intensity, urinary symptoms, and quality of life (National Institutes of Health-Chronic Prostatitis Symptom Index Total Score) | 1 | 49 | 49 | SMD=-1.49 [-1.94, -1.04] | very low | RH: thermal therapy |
| | rESWT vs RH | Pain intensity (Visual Analogue Scale) | 1 | 49 | 49 | SMD=-0.68 [-1.09, -0.28] | very low | |
| | **Coccydynia** | | | | | | | |
| | rESWT vs RH | Pain intensity (Visual Analogue Scale) | 1 | 20 | 21 | SMD=-0.74 [-1.19, -0.29] | very low | RH: interferential current therapy and short-wave diathermy |
| | **Stenosing tenosynovitis** | | | | | | | |
| | rESWT vs MT | Pain intensity (Visual Analogue Scale) | 2 | 70 | 70 | SMD=-1.52 [-2.27, -0.77] | very low | MT: indomethacin |
| | rESWT vs MT | Function (Cooney Score) | 2 | 70 | 70 | SMD=1.47 [1.09, 1.84] | very low | |
| | **Achilles tendinopathy** | | | | | | | |
| | rESWT vs RH | Pain intensity (Numerical Rating Scale) | 1 | 25 | 25 | SMD=-0.86 [-1.44, -0.27] | very low | RH: eccentric exercises |
| | rESWT vs RH | Symptom (Victorian Institute of Sport Assessment-Achilles Questionnaire) | 1 | 25 | 25 | SMD=1.40 [0.78, 2.03] | very low | |

| | | | | | | | | |
|---|---|---|---|---|---|---|---|---|
| Medium energy | rESWT vs RH | Pain intensity (Pain Pressure Threshold) | 1 | 22 | 23 | SMD=1.06 [0.47, 1.66] | very low | RH: eccentric exercises |
| | **Small Effect with Low or Very low Certainty** | | | | | | | |
| | **Plantar heel pain** | | | | | | | |
| | rESWT vs SESWT | Pain intensity (Numerical Rating Scale) | 1 | 125 | 118 | SMD=-0.29 [-0.55, -0.04] | low | SESWT: shockwave of the same sound without energy stimulation |
| | **Carpal tunnel syndrome** | | | | | | | |
| | rESWT vs UC | Pain intensity (Visual Analogue Scale) | 1 | 26 | 23 | SMD=0.35 [0.06, 0.63] | very low | UC: splint therapy |
| | **Plantar fasciitis** | | | | | | | |
| | rESWT vs RH | Function (Foot Function Index) | 1 | 40 | 40 | SMD=0.33 [0.05, 0.62] | very low | RH: ultrasound therapy or kinesiology taping or foot orthotics or laser therapy |
| | **Large or Moderate Negative Effect with Low or Very low Certainty** | | | | | | | |
| | **Achilles tendinopathy** | | | | | | | |
| | rESWT vs SESWT | Pain intensity, function and alignment (American Orthopedic Foot and Ankle Society Score) | 1 | 22 | 23 | SMD=-0.95 [-1.57, -0.33] | low | SESWT: shockwave without energy stimulation |
| | **Cervical spondylotic radiculopathy** | | | | | | | |
| | rESWT vs TCMET | Function (Neck Disability Index) | 1 | 29 | 31 | SMD=0.76 [0.23, 1.28] | very low | TCMET: tuina |
| | **Calcific tendinitis of the shoulder** | | | | | | | |
| | rESWT vs DI | Function (Constant-Murley Score) | 1 | 30 | 30 | SMD=-1.38 [-1.95, -0.81] | very low | DI: betamethasone or lidocaine or ropivacaine |
| | rESWT vs DI | Function and symptom (Oxford Shoulder Score) | 1 | 30 | 30 | SMD=3.59 [2.75, 4.42] | very low | |
| Low energy | **Large Positive Effect with Moderate Certainty** | | | | | | | |
| | **Lateral epicondylitis** | | | | | | | |
| | rESWT vs RH | Pain intensity (Visual Analogue Scale) | 5 | 177 | 185 | SMD=-1.53 [-2.26, -0.81] | moderate | RH: ultrasound therapy or wrist extension splint or exercises |
| | **Large or Moderate Positive Effect with Very low Certainty** | | | | | | | |
| | **Post-stroke upper limb spasticity** | | | | | | | |
| | rESWT vs RH | Symptom (Modified Ashworth Scale) | 1 | 53 | 53 | SMD=-0.76 [-1.03, -0.48] | low | RH: transcutaneous electrical nerve stimulation |
| | rESWT vs RH | Funtion (Fugl-Meyer Assessment Scale) | 1 | 53 | 53 | SMD=0.89 [0.52, 1.27] | low | |
| | **Post-stroke shoulder-hand syndrome** | | | | | | | |
| | rESWT vs SESWT | Pain intensity (Visual Analogue Scale) | 1 | 17 | 17 | SMD=-0.90 [-1.31, -0.49] | low | SESWT: shockwave without energy stimulation |
| | **Achilles tendinopathy** | | | | | | | |
| | rESWT vs RH | Pain intensity (Visual Analogue Scale) | 4 | 138 | 138 | SMD=-1.05 [-1.41, -0.69] | low | RH: eccentric exercises or ultrashort wave radiation therapy or stability training or stretching exercises or ultrasound therapy |
| | rESWT vs NT | Pain intensity (Numerical Rating Scale) | 1 | 25 | 25 | SMD=-0.80 [-1.17, -0.42] | low | |
| | rESWT vs NT | Symptom (Victorian Institute of Sport Assessment-Achilles Questionnaire) | 1 | 25 | 25 | SMD=0.87 [0.49, 1.25] | low | |
| | rESWT vs NT | Pain intensity (Pain Pressure Threshold) | 1 | 25 | 25 | SMD=0.72 [0.15, 1.30] | low | |
| | **Post-stroke lower limb spasticity** | | | | | | | |
| | rESWT vs NT | Funtion (Passive Range of Motion) | 2 | 31 | 31 | SMD=0.86 [0.34, 1.39] | low | |
| | **Plantar fasciitis** | | | | | | | |
| | rESWT vs SESWT | Pain intensity (Visual Analogue Scale) | 1 | 16 | 21 | SMD=-0.51 [-0.80, -0.21] | low | SESWT: shockwave without energy stimulation |
| | **Knee osteoarthritis** | | | | | | | |
| | rESWT vs SESWT | Pain intensity, stiffness and physical function (Western Ontario and McMaster Universities Arthritis Index) | 1 | 32 | 32 | SMD=-1.57 [-2.13, -1.01] | low | SESWT: shockwave without energy stimulation |
| | rESWT vs RH | Funtion (Range of Motion) | 1 | 20 | 20 | SMD=1.77 [1.03, 2.52] | low | RH: kinesiotherapy |

| | | | | | | | | |
|---|---|---|---|---|---|---|---|---|
| Low energy | rESWT vs DI | Pain intensity (Visual Analogue Scale) | 2 | 82 | 82 | SMD=-1.36 [-1.77, -0.95] | very low | DI: hyaluronic acid |
| | rESWT vs DI | Pain intensity, stiffness and physical function (Western Ontario and McMaster Universities Arthritis Index) | 2 | 82 | 82 | SMD=-1.16 [-1.49, -0.83] | very low | |
| | **Lateral epicondylitis** | | | | | | | |
| | rESWT vs SESWT | Funtion and symptom (Patient-Rated Tennis Elbow Evaluation Questionnaire) | 1 | 29 | 28 | SMD=1.29 [0.89, 1.69] | low | SESWT: shockwave with minimal energy stimulation |
| | rESWT vs DI | Function (Grip Strength) | 1 | 20 | 19 | SMD=2.18 [1.37, 2.99] | very low | DI: triamcinolone acetonide and lidocaine |
| | rESWT vs TCMET | Function (Grip Strength) | 1 | 23 | 23 | SMD=0.86 [0.25, 1.47] | very low | TCMET: manual acupuncture |
| | **Carpal tunnel syndrome** | | | | | | | |
| | rESWT vs RH | Funtion and symptom (Boston Carpal Tunnel Syndrome Questionnaire) | 1 | 36 | 36 | SMD=-1.22 [-2.34, -0.10] | very low | RH: ultrasound therapy |
| | **Myofascial pain syndrome** | | | | | | | |
| | rESWT vs DI | Funtion (Range of Motion) | 1 | 26 | 29 | SMD=2.06 [1.39, 2.72] | very low | DI: triamcinolone acetonide and lidocaine and mecobalamin |
| | rESWT vs DI | Function (Oswestry Disability Index) | 1 | 27 | 27 | SMD=-4.52 [-5.55, -3.48] | very low | |
| | **Cerebral palsy spasticity** | | | | | | | |
| | rESWT vs SESWT | Symptom (Modified Ashworth Scale) | 2 | 35 | 35 | SMD=-4.96 [-8.36, -1.56] | very low | SESWT: shockwave of the same sound without energy stimulation |
| | rESWT vs SESWT | Funtion (Range of Motion) | 2 | 35 | 35 | SMD=4.51 [1.31, 7.71] | very low | |
| | rESWT vs SESWT | Structure (Plantar Surface Area) | 1 | 10 | 10 | SMD=4.48 [3.64, 5.32] | very low | |
| | **Chronic prostatitis/ chronic pelvic pain syndrome** | | | | | | | |
| | rESWT vs MT | Pain intensity, urinary symptoms, and quality of life (National Institutes of Health-Chronic Prostatitis Symptom Index Total Score) | 3 | 83 | 70 | SMD=-1.74 [-2.78, -0.70] | very low | MT: α-blocker or nonsteroidal anti-inflammatory drugs or celecoxib or tamsulosin or antibiotics or muscle relaxants |
| | rESWT vs MT | Pain intensity (National Institutes of Health-Chronic Prostatitis Symptom Index Pain Score) | 1 | 33 | 30 | SMD=-3.52 [-4.56, -2.49] | very low | |
| | rESWT vs MT | Symptom (National Institutes of Health-Chronic Prostatitis Symptom Index Urinary Score) | 1 | 33 | 30 | SMD=-0.44 [-0.73, -0.15] | very low | |
| | rESWT vs MT | Quality of life (National Institutes of Health-Chronic Prostatitis Symptom Index Quality of Life Score) | 1 | 33 | 30 | SMD=-3.29 [-4.92, -1.67] | very low | |
| | **Stenosing tenosynovitis** | | | | | | | |
| | rESWT vs RH | Function (Cooney Score) | 1 | 36 | 36 | SMD=2.09 [1.51, 2.67] | very low | RH: ultrashort wave radiation therapy or light therapy |
| | rESWT vs RH | Funtion (Range of Motion) | 1 | 32 | 32 | SMD=2.09 [1.51, 2.67] | very low | |
| | rESWT vs MT | Pain intensity (Visual Analogue Scale) | 3 | 104 | 102 | SMD=-1.29 [-2.54, -0.05] | very low | MT: diclofenac or indometacin |
| | rESWT vs MT | Function (Cooney Score) | 2 | 74 | 72 | SMD=1.76 [0.15, 3.38] | very low | |
| | rESWT vs TCMET | Pain intensity (Visual Analogue Scale) | 1 | 30 | 30 | SMD=-1.18 [-1.73, -0.62] | very low | TCMET: tuina |
| | rESWT vs TCMET | Function (Cooney Score) | 1 | 30 | 30 | SMD=1.14 [0.60, 1.69] | very low | |
| | **Plantar heel pain** | | | | | | | |
| | rESWT + TCMET vs MT | Pain intensity (Visual Analogue Scale) | 1 | 30 | 30 | SMD=-0.59 [-1.11, -0.07] | very low | TCMET: tuina; MT: celecoxib |
| | rESWT vs DI | Pain intensity (Visual Analogue Scale) | 1 | 40 | 40 | SMD=-1.65 [-2.24, -1.05] | very low | DI: triamcinolone acetonide and mecobalamin and lidocaine |
| | rESWT vs TCMET | Function (Maryland Foot Score) | 1 | 39 | 36 | SMD=0.86 [0.17, 1.56] | very low | TCMET: small needle-knife therapy |
| | rESWT vs TCMET + DI | Pain intensity (Numerical Rating Scale) | 1 | 32 | 32 | SMD=1.86 [0.78, 2.95] | very low | TCMET: small needle-knife therapy; DI: triamcinolone acetonide and lidocaine |
| | rESWT vs TCMET + DI | Pain intensity (Visual Analogue Scale) | 1 | 38 | 38 | SMD=-2.09 [-3.47, -0.71] | very low | TCMET: small needle-knife therapy; DI: betamethasone and lidocaine |
| | rESWT vs TCMET + DI | Quality of life (36-item Short-Form) | 1 | 38 | 38 | SMD=1.69 [1.18, 2.19] | very low | |
| | **Osgood-schlatter disease** | | | | | | | |
| | rESWT vs RH | Pain intensity (Visual Analogue Scale) | 1 | 30 | 30 | SMD=-1.84 [-2.45, -1.23] | very low | RH: ultrasound diathermy |
| | **Calcific tendinitis of the shoulder** | | | | | | | |

| | | | | | | | | |
|---|---|---|---|---|---|---|---|---|
| Low energy | rESWT vs TCMET | Pain intensity (Visual Analogue Scale) | 1 | 60 | 60 | SMD=-1.09 [-1.46, -0.71] | very low | TCMET: electro-acupuncture |
| | rESWT vs TCMET | Function (Constant-Murley Score) | 1 | 60 | 60 | SMD=0.80 [0.31, 1.28] | very low | |
| | **Patellar tendinopathy** | | | | | | | |
| | rESWT vs TCMET + RH | Pain intensity (Visual Analogue Scale) | 1 | 63 | 63 | SMD=-5.54 [-7.22, -3.86] | very low | TCMET: acupuncture or tuina; RH: ultrasound therapy or microwave therapy or infrared therapy |
| | rESWT vs MT | Pain intensity (Visual Analogue Scale) | 1 | 48 | 48 | SMD=-7.25 [-8.37, -6.12] | very low | MT: diclofenac |
| | **Pes anserine tendinopathy** | | | | | | | |
| | rESWT vs DI | Pain intensity (Visual Analogue Scale) | 1 | 73 | 73 | SMD=-1.00 [-1.35, -0.66] | very low | DI: triamcinolone acetonide and lidocaine |
| | **Diabetic foot ulcers** | | | | | | | |
| | rESWT vs MT | Pain intensity (Visual Analogue Scale) | 1 | 33 | 33 | SMD=-4.56 [-5.50, -3.63] | very low | MT: routine medication |
| | **Bone marrow edema** | | | | | | | |
| | rESWT vs MT | Pain intensity (Visual Analogue Scale) | 1 | 20 | 20 | SMD=-1.91 [-2.67, -1.15] | very low | MT: diclofenac and alendronate |
| | rESWT vs MT | Pain intensity, function and alignment (American Orthopedic Foot and Ankle Society Score) | 1 | 20 | 20 | SMD=1.42 [0.72, 2.13] | very low | |
| | **Small Effect with Low Certainty** | | | | | | | |
| | **Lateral epicondylitis** | | | | | | | |
| | rESWT vs SESWT | Function (Grip Strength) | 2 | 54 | 53 | SMD=0.47 [0.09, 0.86] | low | SESWT: shockwave with minimal energy stimulation or without energy stimulation |
| | **Large or Moderate Negative Effect with Very low Certainty** | | | | | | | |
| | **Stenosing tenosynovitis** | | | | | | | |
| | rESWT vs DI | Pain intensity (Visual Analogue Scale) | 1 | 30 | 30 | SMD=1.69 [1.27, 2.12] | very low | DI: dexamethasone and lidocaine |
| | rESWT vs DI | Function (Cooney Score) | 1 | 30 | 30 | SMD=-1.09 [-1.47, -0.70] | very low | |
| | **Plantar fasciitis** | | | | | | | |
| | rESWT vs RH | Pain intensity, function and alignment (American Orthopedic Foot and Ankle Society Score) | 1 | 23 | 23 | SMD=-0.80 [-1.41, -0.20] | very low | RH: deep muscle stimulator |
| | rESWT vs RH | Pain intensity and function (Roles and maudsley score) | 1 | 23 | 23 | SMD=0.84 [0.24, 1.45] | very low | |
| | rESWT vs RH | Function (Foot Function Index) | 1 | 32 | 32 | SMD=0.66 [0.15, 1.16] | very low | RH: laser therapy |

Note: C, control group; CI, confidence interval; CoE, certainty of evidence assessed using the Grading of Recommendations Assessment, Development and Evaluation approach; DI, drug injection; MT, medication therapy; NT, no treatment; RCTs, randomised controlled trials; rESWT, radial extracorporeal shockwave therapy; RH, rehabilitation; SESWT, sham extracorporeal shockwave therapy; SMD, standardized mean difference; T, treatment group; TCMET, traditional Chinese medicine external therapy; UC, usual care.
