## Supplementary Figure S10 for "Effectiveness and Safety of Type- and Energy-based Extracorporeal Shockwave Therapy in Clinical Practice: Umbrella Review and Evidence Mapping"

### Supplementary Figure S10 Distribution of adjunctive focused extracorporeal shockwave therapy evidence

| Energy level | Disease or condition | Outcome | No of RCTs | T sample | C sample | Effect estimate (95%CI) | CoE | Control group intervention |
|---|---|---|---|---|---|---|---|---|
| High energy | **Large or Moderate Positive Effect with Very low Certainty** | | | | | | | |
| | **Bone marrow edema** | | | | | | | |
| | fESWT + RH vs RH + MT | Pain intensity (Visual Analogue Scale) | 1 | 40 | 40 | SMD=-1.51 [-2.02, -1.00] | very low | RH: partial weight-bearing gait and walking aids; MT: alendronate and alprostadil |
| | fESWT + RH vs RH + MT | Pain intensity, stiffness and physical function (Western Ontario and McMaster Universities Arthritis Index) | 1 | 40 | 40 | SMD=-1.04 [-1.38, -0.71] | very low | |
| | **Frozen shoulder** | | | | | | | |
| | fESWT + RH vs RH | Pain intensity (Visual Analogue Scale) | 1 | 42 | 42 | SMD=-1.39 [-1.87, -0.91] | very low | RH: manual therapy |
| | **Rotator cuff tear** | | | | | | | |
| | fESWT + RH + TCMET vs RH + TCMET | Pain intensity (Visual Analogue Scale) | 1 | 40 | 40 | SMD=-1.98 [-2.52, -1.43] | very low | RH: mobility exercises and transcutaneous electrical nerve stimulation; TCMET: herbal fumigation |
| | fESWT + RH + TCMET vs RH + TCMET | Function (University of California at Los Angeles Shoulder Score) | 1 | 40 | 40 | SMD=0.62 [0.35, 0.89] | very low | |
| | fESWT + RH + TCMET vs RH + TCMET | Function (Constant-Murley Score) | 1 | 40 | 40 | SMD=1.35 [0.78, 1.91] | very low | |
| | **Achilles tendinopathy** | | | | | | | |
| | fESWT + TCMET vs TCMET | Pain intensity (Visual Analogue Scale) | 1 | 60 | 60 | SMD=-1.61 [-2.18, -1.04] | very low | TCMET: electro-acupuncture |
| | **Plantar fasciitis** | | | | | | | |
| | fESWT + RH vs RH | Pain intensity (Visual Analogue Scale) | 1 | 30 | 30 | SMD=-0.74 [-1.27, -0.22] | very low | RH: stretching exercises and muscle energy techniques and kinesiology taping |
| | fESWT + RH vs RH | Pain intensity, function and alignment (American Orthopedic Foot and Ankle Society Score) | 1 | 30 | 30 | SMD=1.87 [1.26, 2.49] | very low | |
| | fESWT + RH vs RH | Function (Range of Motion) | 1 | 30 | 30 | SMD=1.11 [0.57, 1.66] | very low | |
| | **Osteonecrosis of the femoral head** | | | | | | | |
| | fESWT + TCMIT + TCMET vs TCMIT + TCMET + MT | Function and symptom (Harris Hip Score) | 1 | 30 | 30 | SMD=0.93 [0.40, 1.47] | very low | TCMIT: yishenqiangguwan; TCMET: warm acupuncture and tuina; MT: calcium carbonate |
| | fESWT + TCMIT + TCMET vs TCMIT + TCMET + MT | Pain intensity (Visual Analogue Scale) | 1 | 30 | 30 | SMD=-2.31 [-2.97, -1.64] | very low | |
| | fESWT + TCMIT vs TCMIT | Function and symptom (Harris Hip Score) | 1 | 35 | 35 | SMD=0.61 [0.33, 0.89] | very low | TCMIT: gugutouhuaisiyujiaonang |
| | fESWT + MT vs SESWT + MT | Function and symptom (Harris Hip Score) | 1 | 35 | 35 | SMD=0.52 [0.28, 0.77] | very low | SESWT: shockwave with minimal energy stimulation; MT: celecoxib and alendronate |
| | fESWT + MT vs SESWT + MT | Pain intensity (Visual Analogue Scale) | 1 | 35 | 35 | SMD=-0.77 [-1.46, -0.09] | very low | |
| Medium energy | **Moderate Positive Effect with Moderate Certainty** | | | | | | | |
| | **Frozen shoulder** | | | | | | | |
| | fESWT + RH vs RH | Pain intensity (Visual Analogue Scale) | 4 | 171 | 169 | SMD=-0.70 [-0.94, -0.46] | moderate | RH: short wave diathermy or manual therapy or massage or joint mobilization |
| | **Large or Moderate Positive Effect with Low or Very low Certainty** | | | | | | | |
| | **Diabetic foot ulcers** | | | | | | | |
| | fESWT + SC vs SC | Structure (Wound Surface Area) | 1 | 10 | 11 | SMD=-1.49 [-2.48, -0.50] | low | SC: reference to Danish national clinical guidelines |
| | **Rotator cuff tendinopathy** | | | | | | | |
| | fESWT + TCMET vs TCMET | Pain intensity (Visual Analogue Scale) | 1 | 40 | 40 | SMD=-5.94 [-8.73, -3.15] | low | TCMET: electro-acupuncture |
| | fESWT + TCMET vs TCMET | Function (Constant-Murley Score) | 1 | 40 | 40 | SMD=1.60 [0.14, 3.05] | low | |
| | **Rotator cuff tear** | | | | | | | |

| | | | | | | | | |
|---|---|---|---|---|---|---|---|---|
| Medium energy | fESWT + TCMIT +RH vs TCMIT +RH | Pain intensity (Visual Analogue Scale) | 1 | 60 | 60 | SMD=-2.20 [-3.40, -1.00] | low | TCMIT: xujinjiegu decoction; RH: manual therapy and massage |
| | fESWT + TCMIT +RH vs TCMIT +RH | Function (Constant-Murley Score) | 1 | 60 | 60 | SMD=1.22 [0.76, 1.67] | low | |
| | **Knee osteoarthritis** | | | | | | | |
| | fESWT + RH vs RH | Pain intensity (Visual Analogue Scale) | 1 | 19 | 28 | SMD=-1.34 [-1.92, -0.75] | low | RH: muscular strength exercises and ultrasound therapy |
| | fESWT + RH vs RH | Symptom (Lequesne Index) | 1 | 28 | 27 | SMD=-2.23 [-2.91, -1.54] | low | |
| | fESWT + RH vs RH | Function (Range of Motion) | 1 | 27 | 43 | SMD=0.99 [0.42, 1.55] | low | |
| | fESWT + DI vs DI | Pain intensity (Visual Analogue Scale) | 1 | 40 | 40 | SMD=-1.07 [-1.54, -0.60] | low | DI: hyaluronic acid |
| | fESWT + DI vs DI | Symptom (Lequesne Index) | 1 | 28 | 27 | SMD=-0.58 [-1.03, -0.13] | low | |
| | fESWT + MT vs MT | Pain intensity, stiffness and physical function (Western Ontario and McMaster Universities Arthritis Index) | 1 | 27 | 43 | SMD=-1.96 [-2.47, -1.45] | low | MT: glucosamine |
| | **Post-burn pathological scar** | | | | | | | |
| | fESWT + RH vs SESWT + RH | Pain intensity (Numerical Rating Scale) | 2 | 43 | 45 | SMD=-1.13 [-1.58, -0.68] | low | SESWT: shockwave of the same sound without energy stimulation; RH: comprehensive rehabilitation therapy |
| | fESWT + RH vs SESWT + RH | Structure (Scar Thickness) | 2 | 43 | 45 | SMD=-0.75 [-1.19, -0.32] | low | |
| | fESWT + RH vs SESWT + RH | Symptom (Transepidermal water loss) | 1 | 20 | 20 | SMD=-1.61 [-2.33, -0.88] | low | |
| | fESWT + RH vs RH | Pain intensity (Visual Analogue Scale) | 2 | 63 | 65 | SMD=-0.78 [-1.14, -0.42] | low | RH: comprehensive rehabilitation therapy |
| | fESWT + RH vs RH | Structure (Scar Thickness) | 1 | 25 | 23 | SMD=-0.94 [-1.54, -0.34] | low | |
| | **Chronic prostatitis/ chronic pelvic pain syndrome** | | | | | | | |
| | fESWT + MT vs MT | Pain intensity, urinary symptoms, and quality of life (National Institutes of Health-Chronic Prostatitis Symptom Index Total Score) | 1 | 90 | 90 | SMD=-1.07 [-1.57, -0.57] | low | MT: α-blocker or nonsteroidal anti-inflammatory drugs or muscle relaxants |
| | fESWT + MT vs MT | Pain intensity (National Institutes of Health-Chronic Prostatitis Symptom Index Pain Score) | 1 | 90 | 90 | SMD=-1.23 [-1.72, -0.74] | low | |
| | fESWT + MT vs MT | Quality of life (National Institutes of Health-Chronic Prostatitis Symptom Index Quality of Life Score) | 1 | 90 | 90 | SMD=-0.86 [-1.17, -0.55] | low | |
| | fESWT + MT vs SESWT + MT | Pain intensity, urinary symptoms, and quality of life (National Institutes of Health-Chronic Prostatitis Symptom Index Total Score) | 1 | 16 | 15 | SMD=-1.34 [-2.13, -0.55] | very low | SESWT: shockwave without energy stimulation; MT: α-blocker or nonsteroidal anti-inflammatory drugs or antibiotics or muscle relaxants |
| | fESWT + MT vs SESWT + MT | Pain intensity (National Institutes of Health-Chronic Prostatitis Symptom Index Pain Score) | 1 | 16 | 15 | SMD=-0.85 [-1.59, -0.11] | very low | |
| | fESWT + MT vs SESWT + MT | Symptom (National Institutes of Health-Chronic Prostatitis Symptom Index Urinary Score) | 1 | 16 | 15 | SMD=-0.95 [-1.70, -0.21] | very low | |
| | fESWT + MT vs SESWT + MT | Quality of life (National Institutes of Health-Chronic Prostatitis Symptom Index Quality of Life Score) | 1 | 16 | 15 | SMD=-1.31 [-2.09, -0.52] | very low | |
| | fESWT + MT vs SESWT + MT | Pain intensity (Visual Analogue Scale) | 1 | 16 | 15 | SMD=-1.35 [-2.14, -0.56] | very low | |
| | **Frozen shoulder** | | | | | | | |
| | fESWT + RH vs RH | Function (Constant-Murley Score) | 2 | 64 | 64 | SMD=1.53 [1.13, 1.93] | low | RH: short wave diathermy or manual therapy or massage or joint mobilization |
| | fESWT + RH vs RH | Funtion and pain intensity (Shoulder Pain and Disability Index) | 1 | 49 | 49 | SMD=-7.79 [-8.97, -6.61] | very low | |
| | fESWT + DI vs DI | Pain intensity (Visual Analogue Scale) | 1 | 60 | 60 | SMD=-1.73 [-2.15, -1.31] | very low | DI: prednisolone and lidocaine |
| | fESWT + DI vs DI | Function (Constant-Murley Score) | 1 | 60 | 60 | SMD=2.11 [1.66, 2.56] | very low | |
| | fESWT + DI vs DI | Quality of life (Activity of Daily Living Scale) | 1 | 60 | 60 | SMD=5.23 [4.47, 5.99] | very low | |
| | **Osteonecrosis of the femoral head** | | | | | | | |
| | fESWT + RH vs RH | Function and symptom (Harris Hip Score) | 2 | 61 | 61 | SMD=1.86 [0.97, 2.74] | very low | RH: hyperbaric oxygen therapy or ozone therapy |
| | fESWT + RH vs RH | Pain intensity (Visual Analogue Scale) | 1 | 40 | 40 | SMD=-1.22 [-1.70, -0.74] | very low | |
| | fESWT + TCMIT vs TCMIT | Function and symptom (Harris Hip Score) | 1 | 30 | 30 | SMD=4.66 [3.66, 5.66] | very low | TCMIT: xianlinggubaojiaonang |
| | fESWT + TCMIT vs TCMIT | Pain intensity (Visual Analogue Scale) | 1 | 30 | 30 | SMD=-2.15 [-2.80, -1.51] | very low | |
| | fESWT + TCMIT vs TCMIT | Quality of life (36-item Short-Form) | 1 | 30 | 30 | SMD=0.97 [0.78, 1.16] | very low | TCMIT: xianlinggubaojiaonang |

| | | | | | | | | |
|---|---|---|---|---|---|---|---|---|
| Medium energy | **Post-stroke shoulder-hand syndrome** | | | | | | | |
| | fESWT + RH vs RH | Pain intensity (Visual Analogue Scale) | 1 | 35 | 35 | SMD=-0.65 [-1.13, -0.16] | very low | RH: massage and exercises |
| | fESWT + RH vs RH | Funtion (Fugl-Meyer Assessment Scale) | 1 | 35 | 35 | SMD=1.52 [0.99, 2.06] | very low | |
| | fESWT + RH + TCMET vs RH + TCMET | Pain intensity (Visual Analogue Scale) | 2 | 78 | 76 | SMD=-1.06 [-1.40, -0.72] | very low | RH: medium-frequency electrotherapy or mobility exercises; TCMET: manual acupuncture |
| | fESWT + RH + TCMET vs RH + TCMET | Funtion (Fugl-Meyer Assessment Scale) | 2 | 78 | 76 | SMD=3.03 [2.56, 3.50] | very low | |
| | **Calcific tendinitis of the shoulder** | | | | | | | |
| | fESWT + RH + MT vs RH + MT | Pain intensity (Visual Analogue Scale) | 1 | 28 | 28 | SMD=-1.55 [-3.03, -0.06] | very low | DI: diclofenac; RH: microwave therapy |
| | **Plantar heel pain** | | | | | | | |
| | fESWT + TCMET vs TCMET | Function (Maryland Foot Score) | 1 | 30 | 30 | SMD=0.91 [0.60, 1.21] | very low | TCMET: laser needle-knife |
| | **Carpal tunnel syndrome** | | | | | | | |
| | fESWT + MT + DI vs DI + MT | Pain intensity (Visual Analogue Scale) | 1 | 20 | 20 | SMD=-0.85 [-1.50, -0.20] | very low | MT: mecobalamin; DI: betamethasone |
| | fESWT + MT + DI vs DI + MT | Funtion and symptom (Boston Carpal Tunnel Syndrome Questionnaire) | 1 | 20 | 20 | SMD=-0.55 [-1.00, -0.11] | very low | |
| | fESWT + MT + DI vs DI + MT | Quality of life (36-item Short-Form) | 1 | 20 | 20 | SMD=0.69 [0.05, 1.33] | very low | |
| | **Small Effect with Low Certainty** | | | | | | | |
| | **Patellar tendinopathy** | | | | | | | |
| | fESWT + RH vs RH | Symptom (Victorian Institute of Sport Assessment-Patella Questionnaire) | 1 | 66 | 90 | SMD=-0.34 [-0.66, -0.02] | low | RH: eccentric exercises |
| Low energy | **Large or Moderate Positive Effect with Low or Very low Certainty** | | | | | | | |
| | **Erectile dysfunction** | | | | | | | |
| | fESWT + MT vs MT | Function(International Index of Erectile Function-Erectile Function Domain Score) | 1 | 34 | 41 | SMD=0.75 [0.29, 1.21] | low | MT: tadalafil |
| | fESWT + RH vs SESWT + RH | Function(International Index of Erectile Function-Erectile Function Domain Score) | 1 | 21 | 21 | SMD=1.45 [0.76, 2.13] | low | RH: kegel exercises |
| | **Calcific tendinitis of the shoulder** | | | | | | | |
| | fESWT + RH vs SESWT + RH | Function (Constant-Murley Score) | 1 | 47 | 41 | SMD=1.54 [1.06, 2.01] | low | SESWT: shockwave separated by airchambered polyethylene foil; RH: joint mobilization and traction techniques and massage and manual therapy |
| | fESWT + RH vs SESWT + RH | Pain intensity (Visual Analogue Scale) | 1 | 47 | 41 | SMD=-1.93 [-2.44, -1.42] | low | |
| | **Post-stroke upper limb spasticity** | | | | | | | |
| | fESWT + DI vs SESWT + DI | Symptom (Modified Ashworth Scale) | 1 | 32 | 32 | SMD=-0.86 [-1.38, -0.35] | low | SESWT: shockwave of the same sound without energy stimulation; DI: botulinum toxin type A |
| | fESWT + RH + MT vs RH + MT | Symptom (Modified Ashworth Scale) | 2 | 41 | 39 | SMD=-0.86 [-1.32, -0.39] | very low | RH: range of motion exercises or stretching exercises or physical therapy; MT: antispastic medication |
| | fESWT + RH + MT vs RH + MT | Funtion (Passive Range of Motion) | 2 | 41 | 39 | SMD=0.60 [0.13, 1.06] | very low | |
| | **Low back pain** | | | | | | | |
| | fESWT + RH vs DI + RH | Pain intensity (Visual Analogue Scale) | 1 | 27 | 27 | SMD=-1.02 [-1.59, -0.45] | low | DI: triamcinolone and lidocaine; RH: stretching exercises |
| | fESWT + RH vs DI + RH | Function (Oswestry Disability Index) | 1 | 27 | 27 | SMD=-4.52 [-5.55, -3.48] | low | |
| | fESWT + RH vs RH | Pain intensity (Visual Analogue Scale) | 1 | 15 | 15 | SMD=-1.34 [-2.14, -0.54] | very low | RH: manual therapy and stretching exercises |
| | **Carpal tunnel syndrome** | | | | | | | |
| | fESWT + UC vs SESWT + UC | Pain intensity (Visual Analogue Scale) | 1 | 10 | 10 | SMD=-1.43 [-2.21, -0.66] | very low | SESWT: shockwave of the same sound without energy stimulation; UC: wrist splint |
| | **Diabetic foot ulcers** | | | | | | | |
| | fESWT + SC vs SC | Structure (Wound Surface Area) | 2 | 49 | 29 | SMD=-0.69 [-1.18, -0.19] | very low | SC: regular dressing or therapeutic footwear or debridement |
| | **Frozen shoulder** | | | | | | | |
| | fESWT + TCMET vs TCMET | Pain intensity (Visual Analogue Scale) | 1 | 65 | 64 | SMD=-0.68 [-1.03, -0.32] | very low | TCMET: tuina |

| | | | | | | | | |
|---|---|---|---|---|---|---|---|---|
| Low energy | **Rotator cuff tear** | | | | | | | |
| | fESWT + RH vs RH | Pain intensity (Visual Analogue Scale) | 1 | 34 | 34 | SMD=-0.53 [-0.81, -0.25] | very low | RH: comprehensive rehabilitation therapy |
| | **Knee osteoarthritis** | | | | | | | |
| | fESWT + RH vs RH | Pain intensity, stiffness and physical function (Western Ontario and McMaster Universities Arthritis Index) | 2 | 19 | 28 | SMD=-0.84 [-1.47, -0.22] | very low | RH: mobilization with movement or interferential current therapy or exercises |
| | fESWT + RH vs RH | Function (Range of Motion) | 2 | 27 | 43 | SMD=1.46 [0.68, 2.24] | very low | |
| | **Osteonecrosis of the femoral head** | | | | | | | |
| | fESWT + ST vs ST | Function and symptom (Harris Hip Score) | 2 | 73 | 79 | SMD=0.50 [0.26, 0.74] | very low | ST: advanced core decompression or synthetic bone substitutes |
| | fESWT + ST vs ST | Pain intensity (Visual Analogue Scale) | 2 | 73 | 79 | SMD=-0.59 [-0.93, -0.26] | very low | |
| | **Breast cancer-related lymphedema** | | | | | | | |
| | fESWT + RH vs RH | Function (Range of Motion) | 1 | 30 | 30 | SMD=1.08 [0.55, 1.61] | very low | RH: complex decongestive therapy |
| | **Bone marrow edema** | | | | | | | |
| | fESWT + TCMIT vs TCMIT | Function and symptom (Harris Hip Score) | 1 | 20 | 20 | SMD=0.98 [0.35, 1.60] | very low | TCMIT: yishenhuoxue decoction |
| | fESWT + TCMIT vs TCMIT | Quality of life (36-item Short-Form) | 1 | 20 | 20 | SMD=0.83 [0.60, 1.06] | very low | |
| | **Proximal hamstring tendinopathy** | | | | | | | |
| | fESWT + RH + TCMET vs RH + TCMET | Pain intensity (Visual Analogue Scale) | 1 | 50 | 50 | SMD=-7.77 [-11.34, -4.20] | very low | RH: medium-frequency electrotherapy; TCMET: electro-acupuncture |
| | **Small Effect with Very low Certainty** | | | | | | | |
| | **Cerebral palsy spasticity** | | | | | | | |
| | fESWT + RH + TCMET vs RH + TCMET | Funtion (Passive Range of Motion) | 1 | 28 | 32 | SMD=-0.31 [-0.56, -0.05] | very low | RH: exercises; TCMET: tuina |
| | **Rotator cuff tear** | | | | | | | |
| | fESWT + RH vs RH | Function (University of California at Los Angeles Shoulder Score) | 1 | 34 | 34 | SMD=0.44 [0.16, 0.72] | very low | RH: comprehensive rehabilitation therapy |
| | fESWT + RH vs RH | Function (Constant-Murley Score) | 1 | 34 | 34 | SMD=0.48 [0.20, 0.75] | very low | |
| | fESWT + RH vs RH | Function (Range of Motion) | 1 | 34 | 34 | SMD=0.48 [0.20, 0.76] | very low | |
| | **Bone marrow edema** | | | | | | | |
| | fESWT + TCMIT vs TCMIT | Pain intensity (Visual Analogue Scale) | 1 | 20 | 20 | SMD=-0.48 [-0.79, -0.16] | very low | TCMIT: yishenhuoxue decoction |

Note: C, control group; CI, confidence interval; CoE, certainty of evidence assessed using the Grading of Recommendations Assessment, Development and Evaluation approach; DI, drug injection; fESWT, focused extracorporeal shockwave therapy; MT, medication therapy; RCTs, randomised controlled trials; RH, rehabilitation; SC, standard care; SESWT, sham extracorporeal shockwave therapy; SMD, standardized mean difference; ST, surgical treatment; T, treatment group; TCMET, traditional Chinese medicine external therapy; TCMIT, traditional Chinese medicine internal therapy; UC, usual care.
