## Supplementary Figure S11 for "Effectiveness and Safety of Type- and Energy-based Extracorporeal Shockwave Therapy in Clinical Practice: Umbrella Review and Evidence Mapping"

### Supplementary Figure S11 Distribution of adjunctive radial extracorporeal shockwave therapy evidence

| Energy level | Disease or condition | Outcome | No of RCTs | T sample | C sample | Effect estimate (95%CI) | CoE | Control group intervention |
|---|---|---|---|---|---|---|---|---|
| High energy | **Large or Moderate Positive Effect with Low or Very low Certainty** | | | | | | | |
| | **Frozen shoulder** | | | | | | | |
| | rESWT + RH vs RH | Pain intensity (Visual Analogue Scale) | 1 | 26 | 26 | SMD=-0.67 [-1.23, -0.11] | low | RH: joint mobilization |
| | rESWT + RH vs DI + RH | Pain intensity (Visual Analogue Scale) | 1 | 52 | 51 | SMD=-0.84 [-1.08, -0.61] | low | RH: shoulder stretching exercises and pendulum exercises; DI: corticosteroids |
| | **Post-stroke upper limb spasticity** | | | | | | | |
| | rESWT + MT + RH vs SESWT + MT + RH | Symptom(Modified ashworth scale) | 1 | 80 | 80 | SMD=-4.38 [-6.84, -1.92] | low | SESWT: shockwave of the same sound without energy stimulation; RH: not inform; MT: not inform |
| | **Chronic prostatitis/ chronic pelvic pain syndrome** | | | | | | | |
| | rESWT + MT vs MT | Pain intensity, urinary symptoms, and quality of life (National Institutes of Health-Chronic Prostatitis Symptom Index Total Score) | 1 | 52 | 52 | SMD=-1.32 [-1.75, -0.90] | very low | MT: tamsulosin |
| | rESWT + MT vs MT | Pain intensity (National Institutes of Health-Chronic Prostatitis Symptom Index Pain Score) | 1 | 52 | 52 | SMD=-0.89 [-1.29, -0.48] | very low | |
| | rESWT + MT vs MT | Symptom (National Institutes of Health-Chronic Prostatitis Symptom Index Urinary Score) | 1 | 52 | 52 | SMD=-0.69 [-1.33, -0.05] | very low | |
| | rESWT + MT vs MT | Quality of life (National Institutes of Health-Chronic Prostatitis Symptom Index Quality of Life Score) | 1 | 52 | 52 | SMD=-0.80 [-1.20, -0.40] | very low | |
| | **Myofascial pain syndrome** | | | | | | | |
| | rESWT + RH vs RH | Pain intensity (Visual Analogue Scale) | 1 | 15 | 15 | SMD=3.97 [2.67, 5.26] | very low | RH: ultrasound therapy and ischemic compression |
| | rESWT + RH vs RH | Pain intensity (Pain Pressure Threshold) | 1 | 15 | 15 | SMD=3.41 [2.24, 4.57] | very low | |
| | **Patellar tendinopathy** | | | | | | | |
| | rESWT + TCMET vs TCMET | Pain intensity (Visual Analogue Scale) | 1 | 30 | 30 | SMD=-0.77 [-1.30, -0.25] | very low | TCMET: small needle-knife therapy |
| | **Calcific tendinitis of the shoulder** | | | | | | | |
| | rESWT + DI vs DI | Pain intensity (Visual Analogue Scale) | 1 | 20 | 20 | SMD=-1.03 [-1.73, -0.32] | very low | DI: triamcinolone acetonide and lidocaine |
| | rESWT + DI vs DI | Function (Constant-Murley Score) | 1 | 20 | 20 | SMD=0.87 [0.04, 1.70] | very low | |
| | rESWT + RH vs RH | Pain intensity (Numerical Rating Scale) | 1 | 20 | 20 | SMD=-0.70 [-1.15, -0.24] | very low | RH: pulsed shortwave diathermy and ultrasound therapy and transcutaneous electrical nerve stimulation |
| Medium energy | **Large or Moderate Positive Effect with Low or Very low Certainty** | | | | | | | |
| | **Osteonecrosis of the femoral head** | | | | | | | |
| | rESWT + TCMIT vs TCMIT | Function and symptom (Harris Hip Score) | 1 | 64 | 54 | SMD=0.90 [0.52, 1.28] | low | TCMIT: not inform |
| | rESWT + MT + DI + RH vs MT + DI + RH | Function and symptom (Harris Hip Score) | 1 | 37 | 35 | SMD=0.73 [0.25, 1.21] | very low | MT: calcium carbonate and vitamin D3; DI: technetium methylenediphosphonate; RH: ultrashort-wave therapy and exercises |
| | **Knee osteoarthritis** | | | | | | | |
| | rESWT + TCMIT vs TCMIT | Pain intensity (Visual Analogue Scale) | 1 | 46 | 52 | SMD=-0.99 [-1.41, -0.57] | low | TCMIT: duhuojisheng decoction |
| | rESWT + TCMIT vs TCMIT | Pain intensity, stiffness and physical function (Western Ontario and McMaster Universities Arthritis Index) | 1 | 46 | 52 | SMD=-1.60 [-2.06, -1.14] | low | |
| | rESWT + ST vs ST | Pain intensity (Visual Analogue Scale) | 1 | 68 | 73 | SMD=-0.76 [-1.10, -0.41] | low | ST: arthroscopic debridement |

| | | | | | | | | |
|---|---|---|---|---|---|---|---|---|
| Medium energy | rESWT + DI vs DI | Pain intensity, stiffness and physical function (Western Ontario and McMaster Universities Arthritis Index) | 1 | 40 | 38 | SMD=-0.92 [-1.39, -0.45] | low | |
| | rESWT + RH vs RH | Pain intensity (Visual Analogue Scale) | 1 | 30 | 30 | SMD=-5.02 [-6.08, -3.96] | very low | RH: zhanzhuang training |
| | rESWT + RH vs RH | Pain intensity, stiffness and physical function (Western Ontario and McMaster Universities Arthritis Index) | 1 | 30 | 30 | SMD=-1.43 [-2.00, -0.86] | very low | |
| | **Frozen shoulder** | | | | | | | |
| | rESWT + RH + DI vs RH + DI | Pain intensity (Visual Analogue Scale) | 2 | 103 | 103 | SMD=-1.04 [-2.04, -0.03] | low | RH: manual therapy; DI: hyaluronic acid or triamcinolone acetonide or mecobalamin or diprospan or lidocaine |
| | rESWT + RH vs RH | Pain intensity (Visual Analogue Scale) | 4 | 109 | 108 | SMD=-1.62 [-2.76, -0.47] | low | RH: joint mobilizaton |
| | rESWT + RH vs RH | Funtion (Range of Motion) | 3 | 86 | 85 | SMD=4.60 [2.40, 6.80] | low | |
| | rESWT + RH + MT vs RH + MT | Pain intensity (Visual Analogue Scale) | 2 | 130 | 126 | SMD=-1.16 [-2.12, -0.20] | low | MT: corticosteroids or nonsteroidal anti-inflammatory drugs; RH: hot pack therapy and transcutaneous electrical nerve stimulation |
| | rESWT + RH + MT vs RH + MT | Function (Constant-Murley Score) | 2 | 130 | 126 | SMD=0.97 [0.47, 1.47] | low | |
| | rESWT + DI vs DI | Pain intensity (Visual Analogue Scale) | 2 | 96 | 93 | SMD=-1.16 [-2.04, -0.28] | very low | DI: lidocaine or cobamamide or betamethasone or hyaluronic acid |
| | rESWT + TCMET vs TCMET | Pain intensity (Visual Analogue Scale) | 2 | 78 | 78 | SMD=-0.96 [-1.29, -0.63] | very low | TCMET: acupuncture or tuina |
| | rESWT + TCMET vs TCMET | Function (Constant-Murley Score) | 1 | 16 | 20 | SMD=1.74 [0.95, 2.52] | very low | |
| | rESWT + DI vs TCMET + DI | Pain intensity (Visual Analogue Scale) | 1 | 26 | 25 | SMD=-1.79 [-2.45, -1.13] | very low | |
| | rESWT + DI vs TCMET + DI | Quality of life (Activities of Daily Living Scale) | 1 | 26 | 25 | SMD=1.17 [0.57, 1.77] | very low | DI: ropivacaine and triamcinolone acetonide; TCMET: tuina |
| | rESWT + DI vs TCMET + DI | Funtion (Range of Motion) | 1 | 26 | 25 | SMD=4.14 [1.93, 6.34] | very low | |
| | **Post-stroke upper limb spasticity** | | | | | | | |
| | rESWT + RH vs RH | Symptom (Modified Ashworth Scale) | 2 | 45 | 45 | SMD=-1.66 [-2.15, -1.18] | very low | |
| | rESWT + RH vs RH | Pain intensity (Visual Analogue Scale) | 2 | 45 | 45 | SMD=-2.49 [-4.71, -0.27] | very low | RH: range of motion exercises or passive stretching exercises or manual therapy or transcutaneous electrical nerve stimulation or kinesiology tape |
| | rESWT + RH vs RH | Funtion (Range of Motion) | 1 | 15 | 15 | SMD=3.83 [2.57, 5.10] | very low | |
| | rESWT + RH vs RH | Funtion (Fugl-Meyer Assessment Scale) | 1 | 30 | 30 | SMD=5.85 [4.65, 7.04] | very low | |
| | **Post-stroke shoulder-hand syndrome** | | | | | | | |
| | rESWT + RH vs RH | Pain intensity (Visual Analogue Scale) | 1 | 25 | 25 | SMD=-0.83 [-1.41, -0.25] | very low | RH: exercises and transcutaneous electrical nerve stimulation |
| | rESWT + RH vs TCMET + RH | Pain intensity (Visual Analogue Scale) | 1 | 21 | 21 | SMD=-0.71 [-1.34, -0.09] | very low | RH: occupational therapy; TCMET: manual acupuncture |
| | rESWT + RH vs TCMET + RH | Funtion (Fugl-Meyer Assessment Scale) | 1 | 21 | 21 | SMD=1.23 [0.57, 1.90] | very low | |
| | **Rotator cuff tear** | | | | | | | |
| | rESWT + RH vs RH | Pain intensity (Visual Analogue Scale) | 1 | 29 | 28 | SMD=-2.24 [-2.91, -1.57] | very low | |
| | rESWT + RH vs RH | Function (University of California at Los Angeles Shoulder Score) | 2 | 83 | 82 | SMD=1.89 [0.78, 2.99] | very low | |
| | rESWT + RH vs RH | Funtion (Range of Motion) | 2 | 83 | 82 | SMD=1.62 [0.73, 2.52] | very low | RH: mobilization with movement or electromagnetic pulse therapy |
| | rESWT + RH vs RH | Pain intensity (Numerical Rating Scale) | 1 | 54 | 54 | SMD=-1.35 [-1.77, -0.93] | very low | |
| | rESWT + RH vs RH | Function (Constant-Murley Score) | 1 | 54 | 54 | SMD=1.28 [0.99, 1.58] | very low | |
| | rESWT + RH + TCMET vs RH +TCMET | Pain intensity (Visual Analogue Scale) | 3 | 146 | 140 | SMD=-2.20 [-2.53, -1.87] | very low | RH: medium-frequency electrotherapy or ultrashort |

| | | | | | | | | |
|---|---|---|---|---|---|---|---|---|
| Medium energy | rESWT + RH + TCMET vs RH +TCMET | Funtion (Range of Motion) | 1 | 80 | 80 | SMD=1.31 [0.96, 1.65] | very low | wave therapy or hyperthermia therapy or joint mobilization or manual therapy; TCMET: acupuncture or herbal fumigation or tuina |
| | rESWT + RH + TCMET vs RH +TCMET | Pain and Function (American Shoulder and Elbow Surgeons Score) | 1 | 80 | 80 | SMD=0.79 [0.12, 1.46] | very low | |
| | **Myofascial pain syndrome** | | | | | | | |
| | rESWT + TCMET vs TCMET | Pain intensity (Visual Analogue Scale) | 2 | 90 | 90 | SMD=-2.76 [-4.51, -1.01] | very low | TCMET: tuina or electro-acupuncture |
| | rESWT + TCMET vs TCMET | Function (Oswestry Disability Index) | 1 | 60 | 60 | SMD=-9.97 [-11.33, -8.62] | very low | |
| | rESWT + RH vs RH | Pain intensity (Pain Pressure Threshold) | 1 | 20 | 20 | SMD=1.86 [1.10, 2.61] | very low | RH: ultrasound therapy |
| | **Low back pain** | | | | | | | |
| | rESWT + RH vs RH | Pain intensity (Visual Analogue Scale) | 1 | 22 | 21 | SMD=-1.25 [-1.90, -0.59] | very low | RH: hyperthermia therapy and therapeutic exercise and transcutaneous electrical nerve stimulation |
| | rESWT + RH vs RH | Function (Oswestry Disability Index) | 1 | 17 | 15 | SMD=-1.34 [-2.11, -0.56] | very low | |
| | rESWT + RH vs SESWT + RH | Pain intensity (Visual Analogue Scale) | 1 | 40 | 34 | SMD=-0.77 [-1.24, -0.29] | very low | SESWT: shockwave separated by polyethylene cap; RH: stability training |
| | rESWT + TCMET + RH vs TCMET + RH | Funtion (Japanese Orthopaedic Association Score) | 1 | 15 | 15 | SMD=1.98 [1.09, 2.88] | very low | RH: medium-frequency electrotherapy; TCMET: cupping therapy and electro-acupuncture |
| | **Lateral epicondylitis** | | | | | | | |
| | rESWT + RH vs RH | Pain intensity (Visual Analogue Scale) | 1 | 45 | 45 | SMD=-0.52 [-0.94, -0.10] | very low | RH: mobilization with movement |
| | rESWT + RH vs RH | Quality of life (Activities of Daily Living Scale) | 1 | 45 | 45 | SMD=2.66 [2.09, 3.23] | very low | |
| | rESWT + RH vs RH | Function (Grip Strength) | 1 | 43 | 43 | SMD=1.07 [0.49, 1.66] | very low | RH: antagonistic exercises |
| | **Carpal tunnel syndrome** | | | | | | | |
| | rESWT + UC vs SESWT + UC | Pain intensity (Visual Analogue Scale) | 1 | 20 | 20 | SMD=-0.96 [-1.45, -0.47] | very low | SESWT: shockwave of the same sound without energy stimulation; UC: wrist splint |
| | rESWT + UC vs SESWT + UC | Funtion and symptom (Boston Carpal Tunnel Syndrome Questionnaire) | 2 | 49 | 50 | SMD=-1.19 [-1.55, -0.84] | very low | |
| | rESWT + RH vs RH | Pain intensity (Visual Analogue Scale) | 1 | 42 | 42 | SMD=-3.34 [-3.81, -2.86] | very low | UC: wrist splint and home exercises |
| | rESWT + RH vs RH | Funtion and symptom (Boston Carpal Tunnel Syndrome Questionnaire) | 1 | 42 | 42 | SMD=-1.61 [-1.85, -1.36] | very low | |
| | rESWT + DI vs SESWT + DI | Funtion and symptom (Boston Carpal Tunnel Syndrome Questionnaire) | 1 | 32 | 32 | SMD=-1.57 [-2.59, -0.55] | very low | SESWT: shockwave of the same sound without energy stimulation; DI: platelet-rich plasma |
| | rESWT + MT vs MT | Pain intensity (Visual Analogue Scale) | 1 | 35 | 35 | SMD=-5.23 [-6.24, -4.23] | very low | MT: mecobalamin |
| | rESWT + MT vs MT | Funtion and symptom (Boston Carpal Tunnel Syndrome Questionnaire) | 1 | 35 | 35 | SMD=-3.23 [-3.75, -2.72] | very low | |
| | rESWT + RH + MT vs RH + MT | Pain intensity (Visual Analogue Scale) | 1 | 28 | 28 | SMD=-0.95 [-1.34, -0.55] | very low | RH: neural mobilization; MT: mecobalamin |
| | rESWT + RH + MT vs RH + MT | Symptom (Global Symptom Score) | 1 | 28 | 28 | SMD=-0.83 [-1.21, -0.44] | very low | |
| | **Cerebral palsy spasticity** | | | | | | | |
| | rESWT + RH vs RH | Symptom (Modified Ashworth Scale) | 1 | 46 | 46 | SMD=-1.09 [-1.53, -0.65] | very low | RH: exercises or occupational therapy or transcutaneous electrical nerve stimulation or physiotherapy or balance training |
| | rESWT + RH vs RH | Funtion(Gross Motor Function Measure-88) | 2 | 64 | 64 | SMD=1.46 [1.15, 1.77] | very low | |
| | rESWT + RH + DI vs RH + DI | Symptom (Modified Ashworth Scale) | 1 | 29 | 29 | SMD=-0.88 [-1.42, -0.34] | very low | DI: botulinum toxin type A; RH: exercises and occupational therapy and wax therapy and transcutaneous electrical nerve stimulation |
| | rESWT + DI vs DI | Symptom (Modified Ashworth Scale) | 1 | 20 | 20 | SMD=-0.91 [-1.29, -0.53] | very low | DI: botulinum toxin type A |

| | | | | | | | | |
|---|---|---|---|---|---|---|---|---|
| Medium energy | **Calcific tendinitis of the shoulder** | | | | | | | |
| | rESWT + TCMET + RH vs TCMET + RH | Function (Constant-Murley Score) | 1 | 43 | 42 | SMD=1.03 [0.29, 1.78] | very low | TCMET: herbal fumigation; RH: massage |
| | rESWT + DI vs DI | Function (Constant-Murley Score) | 1 | 35 | 35 | SMD=0.69 [0.05, 1.33] | very low | DI: triamcinolone acetonide and lidocaine |
| | rESWT + DI vs DI | Function (University of California at Los Angeles Shoulder Score) | 1 | 35 | 35 | SMD=1.38 [0.18, 2.58] | very low | |
| | **Rotator cuff tendinopathy** | | | | | | | |
| | rESWT + RH vs RH | Function (Constant-Murley Score) | 1 | 54 | 53 | SMD=2.10 [0.80, 3.41] | very low | RH: functional training |
| | rESWT + RH vs RH | Pain intensity (Visual Analogue Scale) | 1 | 54 | 53 | SMD=-0.76 [-1.04, -0.48] | very low | |
| | **Plantar heel pain** | | | | | | | |
| | rESWT + TCMET vs MT | Pain intensity (Visual Analogue Scale) | 1 | 50 | 50 | SMD=-0.75 [-1.15, -0.34] | very low | TCMET: herbal fumigation; MT: ibuprofen |
| | rESWT + TCMET vs TCMET | Pain intensity (Visual Analogue Scale) | 1 | 19 | 19 | SMD=-1.07 [-1.75, -0.38] | very low | TCMET: manual acupuncture |
| | **Small Effect with Very low Certainty** | | | | | | | |
| | **Rotator cuff tear** | | | | | | | |
| | rESWT + RH + TCMET vs RH +TCMET | Quality of life (Modified Barthel Index) | 1 | 44 | 44 | SMD=0.45 [0.03, 0.87] | very low | RH: medium-frequency electrotherapy; TCMET: acupuncture |
| | **Frozen shoulder** | | | | | | | |
| | rESWT + DI vs DI | Funtion (Fugl-Meyer Assessment Scale) | 1 | 40 | 40 | SMD=-0.48 [-0.78, -0.18] | very low | DI: lidocaine and cobamamide |
| | **Large Negative Effect with Very low Certainty** | | | | | | | |
| | **Myofascial pain syndrome** | | | | | | | |
| | rESWT + RH vs RH | Pain intensity (Visual Analogue Scale) | 1 | 20 | 20 | SMD=1.23 [0.55, 1.91] | very low | RH: ultrasound therapy |
| Low energy | **Large or Moderate Positive Effect with Moderate Certainty** | | | | | | | |
| | **Cerebral palsy spasticity** | | | | | | | |
| | rESWT + RH vs RH | Symptom (Modified Ashworth Scale) | 9 | 277 | 273 | SMD=-1.28 [-1.46, -1.09] | moderate | RH: neurodevelopmental therapy or strengthening exercises or proprioceptive neuromuscular facilitation or balance training or gait training or physiotherapy or occupational therapy or speech therapy or orthotic treatment or electromyographic biofeedback therapy or massage or transcutaneous electrical nerve stimulation or wax therapy |
| | rESWT + RH vs RH | Funtion(Gross Motor Function Measure-88) | 6 | 189 | 198 | SMD=0.74 [0.53, 0.94] | moderate | |
| | **Post-stroke lower limb spasticity** | | | | | | | |
| | rESWT + RH vs RH | Symptom (Modified Ashworth Scale) | 6 | 191 | 193 | SMD=-1.18 [-1.39, -0.96] | moderate | RH: neurodevelopmental therapy or bridge exercises or transfer training or balance training or ladder safety training or physiotherapy or electromyographic biofeedback therapy or gait training or occupational therapy or manual therapy or orthotic treatment |
| | rESWT + RH vs RH | Funtion (Fugl-Meyer Assessment Scale) | 8 | 246 | 248 | SMD=1.13 [0.95, 1.32] | moderate | |
| | **Frozen shoulder** | | | | | | | |
| | rESWT + DI vs DI | Pain intensity (Visual Analogue Scale) | 4 | 177 | 177 | SMD=-1.62 [-1.86, -1.38] | moderate | DI: hyaluronic acid or betamethasone or lidocaine or ozone or triamcinolone acetonide or methylprednisolone |

**Low energy**

#### Large or Moderate Positive Effect with Low or Very low Certainty

| | | | | | | | |
|---|---|---|---|---|---|---|---|
| **Diabetic foot ulcers** | | | | | | | |
| rESWT + SC vs SC | Structure (Wound Surface Area) | 2 | 39 | 39 | SMD=-0.81 [-1.27, -0.34] | low | SC: debridement or dressing or pressure relief |
| rESWT + SC vs SC | Structure (Percentage of re-epithelialization) | 2 | 39 | 39 | SMD=1.00 [0.52, 1.47] | low | |
| **Cerebral palsy spasticity** | | | | | | | |
| rESWT + RH vs RH | Funtion (Range of Motion) | 5 | 138 | 135 | SMD=0.94 [0.56, 1.33] | low | RH: neurodevelopmental therapy or strengthening exercises or proprioceptive neuromuscular facilitation or balance training or gait training or physiotherapy or occupational therapy or speech therapy or orthotic treatment or electromyographic biofeedback therapy or massage or transcutaneous electrical nerve stimulation or wax therapy |
| rESWT + RH vs RH | Structure (Plantar Surface Area) | 3 | 122 | 118 | SMD=2.20 [1.04, 3.35] | low | |
| rESWT + RH vs RH | Funtion (Peabody Developmental Motor Scale) | 2 | 38 | 39 | SMD=1.01 [0.50, 1.53] | very low | |
| rESWT + RH vs RH + SESWT | Symptom (Modified Ashworth Scale) | 1 | 10 | 9 | SMD=-1.46 [-2.50, -0.42] | low | SESWT: shockwave separated by gauze; RH: sensory integration therapy and physiotherapy and orthotic treatment |
| rESWT + DI vs DI | Symptom (Modified Ashworth Scale) | 1 | 7 | 8 | SMD=-1.09 [-1.87, -0.30] | very low | DI: botulinum toxin type A |
| rESWT + DI vs DI | Funtion (Passive Range of Motion) | 1 | 7 | 8 | SMD=0.67 [-0.07, 1.42] | very low | |
| **Post-stroke upper limb spasticity** | | | | | | | |
| rESWT + RH vs RH | Symptom (Modified Tradieu Scale) | 1 | 30 | 25 | SMD=0.61 [0.33, 0.88] | low | RH: range of motion exercises or neurodevelopmental therapy or exercises |
| rESWT + RH vs RH | Symptom (Modified Ashworth Scale) | 1 | 24 | 24 | SMD=-1.25 [-1.87, -0.63] | very low | |
| rESWT + RH + TCMET vs RH + TCMET | Symptom (Modified Ashworth Scale) | 4 | 113 | 115 | SMD=-1.63 [-2.77, -0.48] | very low | RH: neurodevelopmental therapy or electromyographic biofeedback therapy or occupational therapy or exercises or balance training or manual therapy; TCMET: manual acupuncture or electro-acupuncture |
| rESWT + RH + TCMET vs RH + TCMET | Funtion (Fugl-Meyer Assessment Scale) | 5 | 148 | 150 | SMD=0.78 [0.50, 1.06] | very low | |
| rESWT + RH + TCMET vs RH + TCMET | Quality of life (Modified Barthel Index) | 3 | 83 | 85 | SMD=0.51 [0.18, 0.85] | very low | |
| rESWT + RH + TCMET vs RH + TCMET | Pain intensity (Visual Analogue Scale) | 1 | 35 | 35 | SMD=-0.97 [-1.47, -0.48] | very low | |
| rESWT + RH + DI vs RH + DI | Symptom (Modified Ashworth Scale) | 1 | 30 | 30 | SMD=-0.74 [-1.05, -0.44] | very low | RH: physiotherapy or neurodevelopmental therapy or proprioceptive neuromuscular facilitation or Rood's approach or exercises; DI: botulinum toxin type A |
| rESWT + RH + DI vs RH + DI | Funtion (Fugl-Meyer Assessment Scale) | 1 | 30 | 30 | SMD=4.47 [3.78, 5.15] | very low | |
| rESWT + RH + DI vs RH + DI | Quality of life (Modified Barthel Index) | 1 | 30 | 30 | SMD=1.49 [0.62, 2.36] | very low | |
| rESWT + TCMIT vs RH | Symptom (Modified Ashworth Scale) | 1 | 30 | 30 | SMD=-4.60 [-6.71, -2.50] | very low | RH: exercises; TCMIT: buyanghuanwu decoction |
| rESWT + TCMIT vs RH | Funtion (Fugl-Meyer Assessment Scale) | 1 | 30 | 30 | SMD=3.37 [1.64, 5.10] | very low | |
| rESWT + TCMIT vs RH | Quality of life (Modified Barthel Index) | 1 | 30 | 30 | SMD=0.95 [0.38, 1.52] | very low | |
| rESWT + MT vs RH + MT | Funtion (Fugl-Meyer Assessment Scale) | 1 | 30 | 30 | SMD=1.41 [0.84, 1.98] | very low | RH: range of motion exercises; MT: baclofen |
| rESWT + RH vs RH + SESWT | Symptom (Modified Ashworth Scale) | 1 | 28 | 28 | SMD=-0.77 [-1.20, -0.33] | very low | RH: neurodevelopmental therapy and range of motion exercises and balance training; SESWT: shockwave separated by gauze |
| rESWT + TCMET vs RH | Symptom (Modified Ashworth Scale) | 2 | 43 | 43 | SMD=-1.76 [-3.01, -0.52] | very low | RH: exercises or massage or neurodevelopmental therapy; TCMET: manual acupuncture and moxibustion |
| rESWT + TCMET vs RH | Funtion (Fugl-Meyer Assessment Scale) | 2 | 43 | 43 | SMD=1.16 [0.70, 1.62] | very low | RH: exercises or massage or neurodevelopmental therapy; TCMET: manual acupuncture and moxibustion |
| **Rotator cuff tendinopathy** | | | | | | | |

| | | | | | | | | |
|---|---|---|---|---|---|---|---|---|
| **Low energy** | rESWT + RH + DI vs SESWT + RH + DI | Function (Constant-Murley Score) | 1 | 52 | 54 | SMD=1.32 [0.22, 2.41] | low | SESWT: shockwave separated by gauze; RH: infrared therapy; DI: lidocaine |
| | rESWT + RH vs TCMET + RH | Function (Constant-Murley Score) | 1 | 52 | 54 | SMD=0.64 [0.12, 1.16] | very low | RH: wax therapy; TCMET: tuina |
| | **Post-stroke lower limb spasticity** | | | | | | | |
| | rESWT + RH + MT vs SESWT + RH + MT | Funtion (Passive Range of Motion) | 2 | 57 | 57 | SMD=0.51 [0.13, 0.88] | low | SESWT: shockwave of the same sound without energy stimulation; MT: not inform; RH: range of motion exercises |
| | rESWT + RH + MT vs SESWT + RH + MT | Funtion (Fugl-Meyer Assessment Scale) | 1 | 27 | 27 | SMD=0.70 [0.03, 1.38] | very low | |
| | rESWT + RH vs RH + SESWT | Funtion (Fugl-Meyer Assessment Scale) | 2 | 32 | 31 | SMD=0.80 [0.29, 1.32] | low | RH: joint mobilization or transfer training or balance training or gait training or neurodevelopmental therapy or weight shifting exercises or facilitation techniques or range of motion exercises and progressive resistance exercise or occupational therapy or visual feedback balance training |
| | rESWT + RH vs RH + SESWT | Funtion (Passive Range of Motion) | 1 | 12 | 11 | SMD=1.06 [0.42, 1.69] | low | |
| | rESWT + RH vs RH + SESWT | Pain intensity (Visual Analogue Scale) | 1 | 12 | 11 | SMD=-1.01 [-1.89, -0.13] | low | |
| | rESWT + RH vs RH + SESWT | Funtion (Range of Motion) | 1 | 17 | 16 | SMD=0.66 [0.16, 1.16] | low | |
| | rESWT + RH vs RH + SESWT | Quality of life (Modified Barthel Index) | 1 | 17 | 16 | SMD=1.28 [0.74, 1.81] | low | |
| | rESWT + RH vs RH + SESWT | Symptom (Modified Tradieu Scale) | 1 | 17 | 16 | SMD=1.28 [0.74, 1.81] | low | |
| | rESWT + RH vs RH + SESWT | Symptom (Composite Spasticity Scale) | 1 | 10 | 10 | SMD=-2.51 [-3.75, -1.28] | very low | |
| | rESWT + RH + DI vs RH + DI | Symptom (Modified Ashworth Scale) | 1 | 20 | 20 | SMD=-1.18 [-1.87, -0.48] | very low | RH: physiotherapy or neurodevelopmental therapy or proprioceptive neuromuscular facilitation or Rood's approach or exercises; DI: botulinum toxin type A |
| | rESWT + RH + DI vs RH + DI | Funtion (Fugl-Meyer Assessment Scale) | 1 | 20 | 20 | SMD=1.07 [0.07, 2.07] | very low | |
| | rESWT + RH + DI vs RH + DI | Quality of life (Modified Barthel Index) | 1 | 20 | 20 | SMD=0.56 [0.14, 0.99] | very low | |
| | rESWT + RH vs RH | Symptom (Composite Spasticity Scale) | 2 | 60 | 60 | SMD=-0.83 [-1.07, -0.59] | very low | RH: neurodevelopmental therapy or bridge exercises or transfer training or balance training or ladder safety training or physiotherapy or electromyographic biofeedback therapy or gait training or occupational therapy or manual therapy or orthotic treatment |
| | rESWT + RH vs RH | Funtion (Passive Range of Motion) | 2 | 60 | 60 | SMD=0.58 [0.34, 0.81] | very low | |
| | rESWT + RH vs RH | Funtion (Range of Motion) | 2 | 75 | 75 | SMD=0.67 [0.44, 0.91] | very low | |
| | rESWT + RH vs RH | Quality of life (Modified Barthel Index) | 1 | 60 | 60 | SMD=1.22 [0.83, 1.61] | very low | |
| | rESWT + RH vs RH | Funtion (Fugl-Meyer Assessment Scale) | 1 | 22 | 21 | SMD=1.10 [0.46, 1.75] | very low | |
| | rESWT + TCMET vs RH | Symptom (Composite Spasticity Scale) | 1 | 32 | 32 | SMD=-0.65 [-1.15, -0.14] | very low | RH: joint mobilization or balance training or gait training or muscle training; TCMET: electro-acupuncture |
| | rESWT + TCMET vs RH | Funtion (Fugl-Meyer Assessment Scale) | 1 | 32 | 32 | SMD=1.05 [0.53, 1.58] | very low | |
| | rESWT + TCMET vs TCMET | Symptom (Composite Spasticity Scale) | 1 | 32 | 32 | SMD=-0.60 [-1.10, -0.09] | very low | TCMET: electro-acupuncture |
| | rESWT + TCMET vs TCMET | Funtion (Fugl-Meyer Assessment Scale) | 1 | 32 | 32 | SMD=0.89 [0.37, 1.40] | very low | |
| | rESWT + TCMET + RH vs RH | Symptom (Modified Ashworth Scale) | 1 | 30 | 30 | SMD=-1.12 [-1.67, -0.57] | very low | RH: postural control or bridge exercises or transfer training or gait training or balance training or exercises; TCMET: electro-acupuncture or scalp acupuncture |
| | rESWT + TCMET + RH vs RH | Funtion (Fugl-Meyer Assessment Scale) | 1 | 25 | 25 | SMD=1.05 [0.45, 1.64] | very low | |
| | rESWT + RH + DI vs SESWT + RH + DI | Symptom (Modified Ashworth Scale) | 1 | 18 | 18 | SMD=-1.10 [-1.60, -0.60] | very low | SESWT: shockwave of the same sound without energy stimulation; DI: botulinum toxin type A; RH: range of motion exercises and balance training and muscle training |
| | rESWT + RH + DI vs SESWT + RH + DI | Funtion (Passive Range of Motion) | 1 | 18 | 18 | SMD=1.17 [0.66, 1.67] | very low | |
| | **Frozen shoulder** | | | | | | | |
| | rESWT + TCMET vs TCMET | Pain intensity (Visual Analogue Scale) | 1 | 20 | 20 | SMD=-1.71 [-2.45, -0.98] | low | TCMET: herbal fumigation |

| | | | | | | | | |
|---|---|---|---|---|---|---|---|---|
| **Low energy** | rESWT + RH vs RH | Pain intensity (Visual Analogue Scale) | 4 | 266 | 266 | SMD=-1.42 [-1.96, -0.88] | low | |
| | rESWT + RH vs RH | Funtion (Range of Motion) | 1 | 31 | 31 | SMD=1.84 [1.24, 2.44] | low | |
| | rESWT + RH vs RH | Pain intensity (Numerical Rating Scale) | 3 | 167 | 165 | SMD=-2.17 [-2.34, -2.00] | low | |
| | rESWT + RH vs RH | Funtion and pain intensity (Shoulder Pain and Disability Index) | 2 | 95 | 95 | SMD=-1.92 [-2.27, -1.57] | very low | RH: manual therapy or microwave therapy or joint mobilization or ultrashort-wave therapy or pulsed radiofrequency treatment or medium-frequency electrotherapy |
| | rESWT + RH vs RH | Funtion (Range of Motion) | 3 | 104 | 104 | SMD=1.43 [0.81, 2.05] | very low | |
| | rESWT + RH vs RH | Quality of life (36-item Short-Form) | 4 | 206 | 204 | SMD=1.99 [0.47, 3.50] | very low | |
| | rESWT + RH vs RH | Function (University of California at Los Angeles Shoulder Score) | 1 | 45 | 45 | SMD=0.47 [0.05, 0.89] | very low | |
| | rESWT + DI vs DI | Funtion (Range of Motion) | 1 | 20 | 20 | SMD=1.89 [1.36, 2.41] | very low | DI: hyaluronic acid or betamethasone or lidocaine or ozone or triamcinolone acetonide or methylprednisolone |
| | rESWT + DI vs DI | Quality of life(Activities of daily living scale) | 1 | 44 | 44 | SMD=0.71 [0.28, 1.14] | very low | |
| | rESWT + TCMET vs RH + TCMET | Pain intensity (Visual Analogue Scale) | 2 | 60 | 60 | SMD=-1.30 [-2.51, -0.10] | very low | RH: ultrashort wave therapy or phototherapy; TCMET: tuina or fufangnanxingzhitonggao |
| | rESWT + TCMET vs DI | Pain intensity (Visual Analogue Scale) | 1 | 91 | 91 | SMD=-0.78 [-1.08, -0.48] | very low | TCMET: balance acupuncture; DI: triamcinolone acetonide and lidocaine |
| | rESWT + TCMET vs DI | Function (Constant-Murley Score) | 1 | 91 | 91 | SMD=0.86 [0.55, 1.16] | very low | |
| | rESWT + RH + TCMET vs TCMET | Pain intensity (Visual Analogue Scale) | 1 | 43 | 43 | SMD=-0.61 [-1.05, -0.18] | very low | |
| | rESWT + RH + TCMET vs TCMET | Function (University of California at Los Angeles Shoulder Score) | 1 | 43 | 43 | SMD=0.59 [0.39, 0.78] | very low | TCMET: manual acupuncture; RH: medium-frequency electrotherapy |
| | rESWT + RH + TCMET vs TCMET | Funtion (Range of Motion) | 1 | 43 | 43 | SMD=0.56 [0.36, 0.75] | very low | |
| | rESWT + TCMET vs TCMET | Pain intensity (Visual Analogue Scale) | 2 | 56 | 56 | SMD=-1.30 [-1.82, -0.78] | very low | TCMET: tuina |
| | rESWT + RH + DI vs RH + DI | Function (Constant-Murley Score) | 1 | 49 | 50 | SMD=1.31 [0.98, 1.65] | very low | RH: range of motion exercises and balance training and muscle training; DI: hyaluronic acid |
| | **Myofascial pain syndrome** | | | | | | | |
| | rESWT + RH + MT vs RH + MT | Pain intensity (Visual Analogue Scale) | 1 | 19 | 18 | SMD=-1.20 [-1.91, -0.50] | low | RH: ultrasound therapy; MT: tizanidine and meloxicam |
| | rESWT + RH + MT vs RH + MT | Function (Neck Disability Index) | 1 | 38 | 36 | SMD=-0.46 [-0.95, 0.04] | very low | |
| | rESWT + MT vs MT | Pain intensity (Visual Analogue Scale) | 2 | 28 | 36 | SMD=-1.71 [-2.91, -0.51] | very low | MT: dichofenac |
| | rESWT + DI vs DI | Pain intensity (Visual Analogue Scale) | 1 | 26 | 29 | SMD=-2.76 [-3.97, -1.54] | very low | DI: triamcinolone acetonide and lidocaine and mecobalamin |
| | rESWT + DI vs DI | Funtion (Range of Motion) | 1 | 26 | 29 | SMD=2.13 [1.46, 2.80] | very low | |
| | rESWT + RH + DI vs RH + DI | Pain intensity (Visual Analogue Scale) | 1 | 43 | 44 | SMD=-0.59 [-0.91, -0.27] | very low | DI: methylprednisolone or lidocaine or mecobalamin; RH: hot pack therapy and exercises |
| | rESWT + TCMET vs TCMET | Pain intensity (Visual Analogue Scale) | 1 | 45 | 45 | SMD=-3.07 [-3.69, -2.45] | very low | TCMET: manual acupuncture and moxibustion |
| | rESWT + TCMET vs TCMET | Function (Oswestry Disability Index) | 1 | 45 | 45 | SMD=-2.84 [-3.43, -2.25] | very low | |
| | rESWT + TCMIT vs TCMET+ TCMIT | Function (Roland Morris Disability Questionnaire) | 1 | 32 | 32 | SMD=-1.37 [-1.92, -0.82] | very low | TCMET: tuina; TCMIT: juanbi decoction |
| | rESWT + TCMIT vs TCMET+ TCMIT | Function (Oswestry Disability Index) | 1 | 32 | 32 | SMD=-1.27 [-1.81, -0.73] | very low | TCMET: tuina; TCMIT: juanbi decoction |
| | **Knee osteoarthritis** | | | | | | | |
| | rESWT + DI vs DI | Pain intensity (Visual Analogue Scale) | 1 | 67 | 65 | SMD=-1.10 [-1.47, -0.73] | low | DI: hyaluronic acid |
| | rESWT + RH vs RH + DI | Pain intensity (Visual Analogue Scale) | 1 | 21 | 21 | SMD=-0.74 [-1.36, -0.11] | very low | RH: ozone therapy; DI: hyaluronic acid |

| | | | | | | | | |
|---|---|---|---|---|---|---|---|---|
| Low energy | rESWT + DI vs TCMET + DI | Pain intensity (Visual Analogue Scale) | 1 | 30 | 30 | SMD=-0.84 [-1.61, -0.06] | very low | DI: ozone; TCMET: manual acupuncture |
| | rESWT + DI vs TCMET + DI | Pain intensity, stiffness and physical function (Western Ontario and McMaster Universities Arthritis Index) | 1 | 30 | 30 | SMD=-1.70 [-2.12, -1.28] | very low | |
| | rESWT + TCMET vs TCMET | Pain intensity (Visual Analogue Scale) | 1 | 38 | 38 | SMD=-0.83 [-1.16, -0.49] | very low | TCMET: herbal fumigation |
| | rESWT + TCMET vs TCMET | Function (Lysholm Score) | 1 | 38 | 38 | SMD=0.67 [0.34, 1.00] | very low | |
| | rESWT + ST vs ST | Pain intensity (Visual Analogue Scale) | 1 | 40 | 40 | SMD=-1.45 [-1.95, -0.96] | very low | ST: arthroscopic knee debridement |
| | rESWT + ST vs ST | Pain intensity, stiffness and physical function (Western Ontario and McMaster Universities Arthritis Index) | 1 | 40 | 40 | SMD=-1.87 [-2.40, -1.34] | very low | |
| | rESWT + ST vs ST | Function (Lysholm Score) | 1 | 40 | 40 | SMD=1.24 [0.76, 1.72] | very low | |
| | rESWT + TCMIT vs TCMIT | Pain intensity, stiffness and physical function (Western Ontario and McMaster Universities Arthritis Index) | 1 | 33 | 32 | SMD=-1.82 [-2.40, -1.23] | very low | TCMIT: huamoyankeli |
| | rESWT + TCMIT vs MT | Pain intensity (Visual Analogue Scale) | 1 | 32 | 32 | SMD=-2.76 [-3.46, -2.07] | very low | TCMIT: herbal fumigation; MT: diclofenac |
| | rESWT + TCMIT vs MT | Pain intensity, stiffness and physical function (Western Ontario and McMaster Universities Arthritis Index) | 1 | 32 | 32 | SMD=-2.40 [-3.05, -1.75] | very low | |
| | **Carpal tunnel syndrome** | | | | | | | |
| | rESWT + UC vs SESWT + UC | Funtion and symptom (Boston Carpal Tunnel Syndrome Questionnaire) | 2 | 55 | 55 | SMD=-1.67 [-2.52, -0.83] | very low | SESWT: shockwave of the same sound without energy stimulation; UC: wrist splint |
| | rESWT + DI vs SESWT + DI | Pain intensity (Visual Analogue Scale) | 1 | 20 | 20 | SMD=-1.62 [-2.28, -0.97] | very low | SESWT: shockwave of the same sound without energy stimulation; DI: triamcinolone acetonide and lidocaine |
| | rESWT + RH vs RH | Pain intensity (Visual Analogue Scale) | 1 | 20 | 20 | SMD=-1.07 [-1.98, -0.16] | very low | RH: transcutaneous electrical nerve stimulation and ultrasound therapy |
| | rESWT + RH vs RH | Funtion and symptom (Boston Carpal Tunnel Syndrome Questionnaire) | 1 | 20 | 20 | SMD=-0.71 [-1.28, -0.15] | very low | |
| | rESWT + MT vs MT | Pain intensity (Visual Analogue Scale) | 1 | 30 | 30 | SMD=-0.69 [-1.21, -0.16] | very low | MT: mecobalamin |
| | rESWT + RH vs RH | Funtion and symptom (Boston Carpal Tunnel Syndrome Questionnaire) | 1 | 20 | 20 | SMD=-0.92 [-1.29, -0.54] | very low | |
| | **Cervical spondylotic radiculopathy** | | | | | | | |
| | rESWT + TCMET vs TCMET | Pain intensity (Visual Analogue Scale) | 3 | 141 | 141 | SMD=-1.54 [-2.37, -0.70] | very low | TCMET: tuina or manual acupuncture or electro-acupuncture |
| | rESWT + TCMET vs TCMET | Function (Neck Disability Index) | 1 | 43 | 43 | SMD=-4.17 [-4.94, -3.41] | very low | |
| | rESWT + TCMIT vs TCMIT | Function (Neck Disability Index) | 1 | 40 | 38 | SMD=-3.77 [-4.52, -3.02] | very low | TCMIT: jingshukeli and gegen decoction |
| | rESWT + RH vs RH | Function (Neck Disability Index) | 1 | 42 | 42 | SMD=-1.40 [-1.88, -0.92] | very low | RH: massage or cervical traction or joint mobilization |
| | rESWT + RH vs RH | Pain intensity (Visual Analogue Scale) | 3 | 90 | 90 | SMD=-1.87 [-2.90, -0.83] | very low | |
| | rESWT + RH vs TCMET + RH | Pain intensity (Visual Analogue Scale) | 1 | 35 | 35 | SMD=-0.62 [-1.10, -0.14] | very low | RH: cervical traction; TCMET: tuina |
| | **Stenosing tenosynovitis** | | | | | | | |
| | rESWT + RH vs RH | Pain intensity (Visual Analogue Scale) | 2 | 56 | 56 | SMD=-1.21 [-1.57, -0.85] | very low | RH: manual therapy or light therapy |
| | rESWT + RH vs RH | Funtion and symptom (Patient-Rated Wrist Evaluation) | 1 | 24 | 24 | SMD=-1.96 [-3.21, -0.71] | very low | |
| | rESWT + RH vs RH | Funtion (Range of Motion) | 1 | 32 | 32 | SMD=1.59 [1.11, 2.07] | very low | RH: manual therapy or light therapy |
| | **Decubitus ulceration** | | | | | | | |
| | rESWT + RH +SC vs SC | Pain intensity (Visual Analogue Scale) | 1 | 30 | 30 | SMD=-0.55 [-1.06, -0.03] | very low | RH: microcurrent therapy; SC: debridement and dressing and air cushion bed |
| | **Rotator cuff tear** | | | | | | | |

| | | | | | | | | |
|---|---|---|---|---|---|---|---|---|
| Low energy | rESWT + TCMIT + RH vs TCMIT + RH | Pain intensity (Visual Analogue Scale) | 1 | 30 | 30 | SMD=-1.08 [-1.62, -0.53] | very low | TCMIT: xujinjiegu decoction; RH: manual therapy |
| | rESWT + TCMIT + RH vs TCMIT + RH | Function (Constant-Murley Score) | 1 | 30 | 30 | SMD=0.61 [0.09, 1.13] | very low | |
| | rESWT + RH vs RH | Pain intensity (Visual Analogue Scale) | 1 | 31 | 31 | SMD=-1.59 [-2.17, -1.02] | very low | RH: water-filtered infrared-A irradiation |
| | rESWT + RH vs RH | Function (University of California at Los Angeles Shoulder Score) | 1 | 31 | 31 | SMD=2.18 [1.54, 2.81] | very low | |
| | rESWT + RH vs RH | Funtion (Range of Motion) | 1 | 31 | 31 | SMD=0.99 [0.47, 1.51] | very low | |
| | rESWT + DI vs DI | Pain intensity (Visual Analogue Scale) | 1 | 24 | 24 | SMD=-1.06 [-1.59, -0.54] | very low | DI: platelet-rich plasma |
| | rESWT + DI vs DI | Function (Constant-Murley Score) | 1 | 24 | 24 | SMD=1.44 [1.12, 1.76] | very low | |
| | **Post-stroke shoulder-hand syndrome** | | | | | | | |
| | rESWT + RH + MT vs TCMET + RH + MT | Pain intensity (Visual Analogue Scale) | 1 | 28 | 28 | SMD=-0.83 [-1.37, -0.28] | very low | TCMET: manual acupuncture; RH: general rehabilitation; MT: general medication |
| | rESWT + RH + MT vs TCMET + RH + MT | Funtion (Fugl-Meyer Assessment Scale) | 1 | 28 | 28 | SMD=0.55 [0.02, 1.09] | very low | |
| | rESWT + RH + TCMET + MT vs TCMET + RH + MT | Pain intensity (Visual Analogue Scale) | 3 | 89 | 89 | SMD=-1.25 [-1.89, -0.61] | very low | TCMET: electro-acupuncture or manual acupuncture; RH: general rehabilitation; MT: general medication |
| | rESWT + RH + TCMET + MT vs TCMET + RH + MT | Funtion (Fugl-Meyer Assessment Scale) | 2 | 50 | 50 | SMD=0.68 [0.27, 1.08] | very low | |
| | rESWT + RH + TCMET + MT vs TCMET + RH + MT | Funtion (Range of Motion) | 1 | 30 | 30 | SMD=0.77 [0.24, 1.30] | very low | |
| | rESWT + RH vs RH | Pain intensity (Visual Analogue Scale) | 3 | 94 | 94 | SMD=-1.60 [-2.20, -1.00] | very low | RH: joint mobilization or low-frequency sound wave therapy or sling exercise training or manual lymphatic drainage technique or transfer training or range of motion exercises |
| | rESWT + RH vs RH | Funtion (Fugl-Meyer Assessment Scale) | 2 | 75 | 75 | SMD=2.83 [0.82, 4.84] | very low | |
| | rESWT + RH vs RH | Funtion (Range of Motion) | 1 | 19 | 19 | SMD=0.83 [0.41, 1.24] | very low | |
| | rESWT + RH vs RH | Function (Constant-Murley Score) | 1 | 29 | 29 | SMD=5.91 [4.68, 7.13] | very low | |
| | rESWT + RH vs RH | Quality of life (Modified Barthel Index) | 1 | 46 | 46 | SMD=0.99 [0.69, 1.30] | very low | |
| | rESWT + RH vs TCMET + RH | Pain intensity (Visual Analogue Scale) | 1 | 43 | 43 | SMD=-1.18 [-1.64, -0.72] | very low | TCMET: manual acupuncture; RH: occupational therapy |
| | rESWT + RH vs TCMET + RH | Funtion (Fugl-Meyer Assessment Scale) | 1 | 43 | 43 | SMD=1.19 [0.73, 1.65] | very low | |
| | rESWT + RH + MT vs RH + MT | Pain intensity (Visual Analogue Scale) | 1 | 32 | 32 | SMD=-0.86 [-1.37, -0.35] | very low | RH: general rehabilitation; MT: general medication |
| | rESWT + RH + MT vs RH + MT | Funtion (Fugl-Meyer Assessment Scale) | 1 | 32 | 32 | SMD=2.90 [2.18, 3.61] | very low | |
| | **Low back pain** | | | | | | | |
| | rESWT + RH vs RH | Pain intensity (Visual Analogue Scale) | 3 | 99 | 98 | SMD=-1.03 [-1.33, -0.73] | very low | RH: posterior pelvic tilt exercise or abdominal stretching exercise or core strength training or respiratory muscle training |
| | rESWT + RH vs RH | Function (Oswestry Disability Index) | 1 | 37 | 36 | SMD=-1.23 [-1.79, -0.68] | very low | |
| | rESWT + RH vs RH | Function (Roland Morris Disability Questionnaire) | 1 | 32 | 32 | SMD=-0.79 [-1.30, -0.28] | very low | |
| | rESWT + RH vs RH | Quality of life (36-item Short-Form) | 1 | 32 | 32 | SMD=0.90 [0.39, 1.42] | very low | |
| | rESWT + MT vs MT | Pain intensity (Visual Analogue Scale) | 1 | 24 | 22 | SMD=-2.36 [-3.55, -1.17] | very low | MT: flurbiprofen |
| | rESWT + MT vs MT | Function (Oswestry Disability Index) | 1 | 24 | 22 | SMD=-1.48 [-2.15, -0.81] | very low | |
| | rESWT + TCMET + RH vs TCMET + RH | Pain intensity (Visual Analogue Scale) | 2 | 56 | 56 | SMD=-0.98 [-1.37, -0.58] | very low | TCMET: tuina or herbal fumigation; RH: interferential |

| | | | | | | | | |
|---|---|---|---|---|---|---|---|---|
| Low energy | rESWT + TCMET + RH vs TCMET + RH | Function (Roland Morris Disability Questionnaire) | 1 | 25 | 25 | SMD=-1.48 [-2.11, -0.85] | very low | [illegible] therapy or core strength training or magneto-thermo-vibration therapy |
| | rESWT + TCMET + RH vs TCMET + RH | Funtion (Quebec Back Pain Disability Scale) | 1 | 31 | 31 | SMD=-0.61 [-1.12, -0.10] | very low | |
| | rESWT + TCMET vs TCMET | Pain intensity (Visual Analogue Scale) | 1 | 60 | 60 | SMD=-1.80 [-2.23, -1.38] | very low | TCMET: herbal paste |
| | rESWT + TCMET vs TCMET | Function (Oswestry Disability Index) | 1 | 60 | 60 | SMD=-2.73 [-3.23, -2.23] | very low | |
| | **Plantar fasciitis** | | | | | | | |
| | rESWT + DI vs DI | Pain intensity (McGill Pain Questionnaire) | 1 | 40 | 40 | SMD=-0.85 [-1.12, -0.59] | very low | DI: lidocaine and betamethasone |
| | rESWT + DI vs DI | Structure (Plantar Fascia Thickness) | 1 | 40 | 40 | SMD=-0.50 [-0.76, -0.24] | very low | |
| | rESWT + DI vs DI | Pain intensity, function and alignment (American Orthopedic Foot and Ankle Society Score) | 1 | 40 | 40 | SMD=0.76 [0.50, 1.02] | very low | |
| | rESWT + RH vs RH | Pain intensity (Visual Analogue Scale) | 1 | 23 | 23 | SMD=-2.90 [-3.74, -2.05] | very low | RH: deep muscle stimulator |
| | rESWT + RH vs RH | Pain intensity, function and alignment (American Orthopedic Foot and Ankle Society Score) | 1 | 23 | 23 | SMD=2.29 [1.53, 3.05] | very low | |
| | rESWT + RH vs RH | Pain intensity and function (Roles and maudsley score) | 1 | 23 | 23 | SMD=-2.28 [-3.04, -1.52] | very low | |
| | **Osteonecrosis of the femoral head** | | | | | | | |
| | rESWT + TCMIT vs TCMIT | Function and symptom (Harris Hip Score) | 2 | 58 | 58 | SMD=0.89 [0.18, 1.61] | very low | TCMIT: boneerosionjiaonang or xiaozhonghuoxue decoction |
| | rESWT + TCMIT vs TCMIT | Pain intensity (Visual Analogue Scale) | 2 | 58 | 58 | SMD=-0.67 [-1.31, -0.04] | very low | |
| | **Breast cancer-related lymphedema** | | | | | | | |
| | rESWT + RH vs RH | Structure (Volume of Lymphedema) | 1 | 20 | 20 | SMD=-2.77 [-3.66, -1.88] | very low | RH: complex decongestive therapy |
| | rESWT + RH vs RH | Structure (Skin Thickness) | 1 | 21 | 22 | SMD=-0.68 [-1.30, -0.06] | very low | |
| | rESWT + RH vs RH | Funtion (Range of Motion) | 1 | 20 | 20 | SMD=1.07 [0.68, 1.46] | very low | |
| | rESWT + RH vs RH | Structure (Modified Rodnan Skin Score) | 1 | 20 | 20 | SMD=-0.82 [-1.28, -0.36] | very low | |
| | rESWT + RH vs RH | Symptom (Breast Cancer and Lymphedema Symptom Experience Index) | 1 | 20 | 20 | SMD=-0.76 [-1.21, -0.30] | very low | |
| | **Post-burn pathological scar** | | | | | | | |
| | rESWT + RH vs RH | Pain intensity (Visual Analogue Scale) | 1 | 46 | 46 | SMD=-0.75 [-1.17, -0.33] | very low | RH: pressure therapy and exercises and audiofrequency current therapy and occupational therapy |
| | **Bone marrow edema** | | | | | | | |
| | rESWT + DI vs DI | Function (Constant-Murley Score) | 1 | 30 | 30 | SMD=1.28 [0.72, 1.83] | very low | DI: hyaluronic acid |
| | **Chronic prostatitis/ chronic pelvic pain syndrome** | | | | | | | |
| | rESWT + MT + TCMIT vs MT + TCMIT | Pain intensity, urinary symptoms, and quality of life (National Institutes of Health-Chronic Prostatitis Symptom Index Total Score) | 1 | 30 | 30 | SMD=-0.77 [-1.04, -0.51] | very low | MT: α-blocker and nonsteroidal anti-inflammatory drugs and tamsulosin and diclofenac; TCMIT: qianliebeixijiaonang |
| | rESWT + MT + TCMIT vs MT + TCMIT | Pain intensity (Numerical Rating Scale) | 1 | 30 | 30 | SMD=-0.93 [-1.20, -0.66] | very low | |
| | **Calcific tendinitis of the shoulder** | | | | | | | |
| | rESWT + RH vs TCMET + RH | Pain intensity (Visual Analogue Scale) | 1 | 28 | 28 | SMD=-2.90 [-3.67, -2.14] | very low | TCMET: directional drug penetration treatment; RH: microwave therapy |
| | rESWT + RH vs TCMET + RH | Function (Constant-Murley Score) | 1 | 28 | 28 | SMD=1.24 [0.67, 1.82] | very low | TCMET: directional drug penetration treatment; RH: microwave therapy |

| | | | | | | | | |
|---|---|---|---|---|---|---|---|---|
| Low energy | **Plantar heel pain** | | | | | | | |
| | rESWT + TCMET vs TCMET | Pain intensity (Visual Analogue Scale) | 2 | 50 | 50 | SMD=-3.48 [-4.12, -2.84] | very low | TCMET: herbal fumigation |
| | rESWT + TCMET vs TCMET | Function (Maryland Foot Score) | 2 | 50 | 50 | SMD=2.55 [-0.46, 5.56] | very low | |
| | rESWT + RH vs DI | Pain intensity (Visual Analogue Scale) | 2 | 63 | 63 | SMD=-1.40 [-2.46, -0.34] | very low | DI: triamcinolone acetonide or lidocaine; RH: plantar aponeurosis traction training |
| | rESWT + RH vs RH | Pain intensity (Visual Analogue Scale) | 1 | 20 | 20 | SMD=-0.85 [-1.23, -0.47] | very low | RH: polarized light therapy or pulsed radiofrequency treatment |
| | rESWT + RH vs RH | Function (Maryland Foot Score) | 1 | 20 | 20 | SMD=1.00 [0.67, 1.32] | very low | |
| | **Patellar tendinopathy** | | | | | | | |
| | rESWT + RH vs RH | Pain intensity (Visual Analogue Scale) | 1 | 40 | 40 | SMD=-0.80 [-1.25, -0.34] | very low | RH: manual therapy |
| | rESWT + RH vs RH | Symptom (Lysholm Score) | 1 | 40 | 40 | SMD=22.29 [18.72, 25.86] | very low | |
| | **Small Effect with Very low Certainty** | | | | | | | |
| | **Carpal tunnel syndrome** | | | | | | | |
| | rESWT + UC vs UC | Funtion and symptom (Boston Carpal Tunnel Syndrome Questionnaire) | 1 | 60 | 75 | SMD=-0.23 [-0.43, -0.03] | very low | UC: wrist splint |
| | **Decubitus ulceration** | | | | | | | |
| | rESWT + RH +SC vs SC | Structure (Pressure ulcer scale for healing) | 1 | 30 | 30 | SMD=-0.42 [-0.71, -0.12] | very low | RH: microcurrent therapy; SC: debridement and dressing and air cushion bed |
| | **Chronic prostatitis/ chronic pelvic pain syndrome** | | | | | | | |
| | rESWT + MT + TCMIT vs MT + TCMIT | Symptom (International Prostatism Symptom Score) | 1 | 30 | 30 | SMD=-0.46 [-0.72, -0.20] | very low | MT: α-blocker and nonsteroidal anti-inflammatory drugs and tamsulosin and diclofenac; TCMIT: qianliebeixijiaonang |
| | **Erectile dysfunction** | | | | | | | |
| | rESWT + MT vs MT | Function(International Index of Erectile Function-Erectile Function Domain Score) | 1 | 81 | 42 | SMD=0.43 [0.07, 0.78] | very low | MT: tadalafil |
| | **Large Negative Effect with Very low Certainty** | | | | | | | |
| | **Knee osteoarthritis** | | | | | | | |
| | rESWT + RH vs RH + DI | Pain intensity, stiffness and physical function (Western Ontario and McMaster Universities Arthritis Index) | 1 | 21 | 21 | SMD=1.51 [0.82, 2.21] | very low | RH: ozone therapy; DI: hyaluronic acid |

Note: C, control group; CI, confidence interval; CoE, certainty of evidence assessed using the Grading of Recommendations Assessment, Development and Evaluation approach; DI, drug injection; MT, medication therapy; RCTs, randomised controlled trials; rESWT, radial extracorporeal shockwave therapy; RH, rehabilitation; SC, standard care; SESWT, sham extracorporeal shockwave therapy; SMD, standardized mean difference; T, treatment group; TCMET, traditional Chinese medicine external therapy; TCMIT, traditional Chinese medicine internal therapy; UC, usual care.
