## Supplementary Figure S12 for "Effectiveness and Safety of Type- and Energy-based Extracorporeal Shockwave Therapy in Clinical Practice: Umbrella Review and Evidence Mapping"

### Supplementary Figure S12 Distribution of single or adjunctive unknown extracorporeal shockwave therapy evidence

| Energy level | Disease or condition | Outcome | No of RCTs | T sample | C sample | Effect estimate (95%CI) | CoE | Control group intervention |
|---|---|---|---|---|---|---|---|---|
| High energy | **Large or Moderate or Small Effect with Low or Very low Certainty** | | | | | | | |
| | **Plantar heel pain** | | | | | | | |
| | ESWT vs SESWT | Pain intensity (Numerical Rating Scale) | 2 | 126 | 125 | SMD=-0.28 [-0.53, -0.03] | low | SESWT: shockwave separated by thin foam or air cushion or without energy stimulation |
| | **Myofascial pain syndrome** | | | | | | | |
| | ESWT vs TCMET | Pain intensity (Visual Analogue Scale) | 1 | 30 | 30 | SMD=-0.85 [-1.38, -0.32] | very low | TCMET: manual acupuncture |
| | **Plantar fasciitis** | | | | | | | |
| | ESWT vs DI | Pain intensity (Visual Analogue Scale) | 1 | 108 | 72 | SMD=0.37 [0.07, 0.67] | very low | DI: methylprednisolone and lidocaine and epinephrine |
| | **Stenosing tenosynovitis** | | | | | | | |
| | ESWT vs TCMET | Pain intensity (Visual Analogue Scale) | 1 | 12 | 12 | SMD=-1.25 [-2.14, -0.36] | very low | TCMET: electro-acupuncture |
| | ESWT + TCMET vs TCMET | Pain intensity (Visual Analogue Scale) | 1 | 12 | 12 | SMD=-1.41 [-2.12, -0.70] | very low | |
| | **Osteonecrosis of the femoral head** | | | | | | | |
| | ESWT + RH + MT vs RH | Quality of life (Modified Barthel Index) | 1 | 56 | 56 | SMD=1.19 [0.79, 1.59] | very low | RH: hyperbaric oxygen therapy; MT: nonsteroidal antiinflammatory drugs and bisphosphate and calcium carbonate |
| Medium energy | **Large or Moderate Effect with Low or Very low Certainty** | | | | | | | |
| | **Plantar heel pain** | | | | | | | |
| | ESWT vs SESWT | Pain intensity (Numerical Rating Scale) | 2 | 39 | 40 | SMD=-0.81 [-1.28, -0.35] | low | SESWT: shockwave of the same sound without energy stimulation |
| | **Lateral epicondylitis** | | | | | | | |
| | ESWT vs DI | Pain intensity (Visual Analogue Scale) | 1 | 20 | 20 | SMD=0.98 [0.32, 1.64] | low | DI: methylprednisolone |
| | ESWT vs DI | Function (Grip Strength) | 1 | 20 | 20 | SMD=-1.46 [-2.16, -0.75] | low | |
| | **Peyronie’s disease** | | | | | | | |
| | ESWT vs ST | Function(International Index of Erectile Function-Erectile Function Domain Score) | 1 | 10 | 10 | SMD=-1.06 [-2.01, -0.11] | very low | ST: autologous perididymal patch grafting |
| | ESWT vs ST | Symptom (Penile Curvature) | 1 | 10 | 10 | SMD=-1.08 [-2.03, -0.13] | very low | |
| | **Rotator cuff tear** | | | | | | | |
| | ESWT + RH vs RH | Function (University of California at Los Angeles Shoulder Score) | 1 | 56 | 64 | SMD=0.61 [0.24, 0.97] | low | RH: range of motion exercises and stretching exercises |
| | ESWT + RH vs RH | Function (Constant-Murley Score) | 1 | 56 | 64 | SMD=1.22 [0.83, 1.62] | low | |
| | ESWT + RH vs RH | Pain intensity (Visual Analogue Scale) | 1 | 56 | 64 | SMD=-0.96 [-1.78, -0.14] | low | |
| | ESWT + RH vs RH | Function (Range of Motion) | 1 | 56 | 64 | SMD=0.82 [0.45, 1.19] | low | |
| | ESWT vs TCMET | Pain intensity (Visual Analogue Scale) | 1 | 15 | 15 | SMD=1.55 [1.07, 2.03] | very low | TCMET: floating acupuncture |
| | ESWT vs TCMET | Function (Range of Motion) | 1 | 15 | 15 | SMD=-2.76 [-4.31, -1.21] | very low | |
| | ESWT vs TCMET | Function (Range of Motion) | 1 | 15 | 15 | SMD=-0.66 [-0.91, -0.42] | very low | |
| | ESWT + TCMET vs TCMET | Pain intensity (Visual Analogue Scale) | 2 | 53 | 53 | SMD=-1.16 [-2.20, -0.13] | very low | TCMET: manual acupuncture or floating acupuncture |
| | ESWT + TCMET vs TCMET | Function (University of California at Los Angeles Shoulder Score) | 1 | 26 | 26 | SMD=5.70 [4.44, 6.97] | very low | |
| | ESWT + TCMET vs TCMET | Quality of life (36-item Short-Form) | 1 | 26 | 26 | SMD=1.32 [0.73, 1.92] | very low | |
| | ESWT + RH +TCMET vs RH + TCMET | Pain intensity (Visual Analogue Scale) | 1 | 32 | 32 | SMD=-4.48 [-5.42, -3.54] | very low | RH: joint mobilization; TCMET: acupuncture and tuina |
| | ESWT + RH +TCMET vs RH + TCMET | Function (Range of Motion) | 1 | 32 | 32 | SMD=2.13 [1.35, 2.92] | very low | RH: joint mobilization; TCMET: acupuncture and tuina |

| | | | | | | | | |
|---|---|---|---|---|---|---|---|---|
| Medium energy | **Low back pain** | | | | | | | |
| | ESWT vs RH | Pain intensity (Visual Analogue Scale) | 2 | 115 | 115 | SMD=-1.53 [-2.30, -0.76] | low | RH: bridging exercises or adductor ball squeeze exercises or abdominal marching exercises or reverse curl exercises |
| | ESWT vs DI | Pain intensity (Visual Analogue Scale) | 1 | 15 | 15 | SMD=-1.80 [-2.67, -0.93] | very low | DI: trigger point injection |
| | ESWT + RH vs SESWT + RH | Pain intensity (Visual Analogue Scale) | 1 | 38 | 36 | SMD=-0.88 [-1.36, -0.40] | very low | SESWT: shockwave of the same sound without energy stimulation; RH: physical exercises |
| | **Post-stroke shoulder-hand syndrome** | | | | | | | |
| | ESWT + RH vs RH | Pain intensity (Visual Analogue Scale) | 1 | 58 | 58 | SMD=-1.26 [-1.66, -0.86] | very low | |
| | ESWT + RH vs RH | Funtion (Fugl-Meyer Assessment Scale) | 1 | 58 | 58 | SMD=1.01 [0.62, 1.40] | very low | RH: exercises |
| | ESWT + RH vs RH | Quality of life (Stroke Specific Quality of Life Scale) | 1 | 58 | 58 | SMD=0.69 [0.32, 1.07] | very low | |
| | ESWT + TCMET + RH vs TCMET + RH | Pain intensity (Visual Analogue Scale) | 2 | 70 | 70 | SMD=-0.76 [-1.14, -0.38] | very low | |
| | ESWT + TCMET + RH vs TCMET + RH | Funtion (Fugl-Meyer Assessment Scale) | 2 | 70 | 70 | SMD=0.86 [0.23, 1.49] | very low | RH: posture exercises or psychotherapy or medium-frequency electrotherapy or barotherapy; TCMET: acupuncture or tuina |
| | ESWT + TCMET + RH vs TCMET + RH | Quality of life (Stroke Specific Quality of Life Scale) | 2 | 70 | 70 | SMD=1.11 [0.64, 1.59] | very low | |
| | **Knee osteoarthritis** | | | | | | | |
| | ESWT vs SESWT | Pain intensity (Visual Analogue Scale) | 1 | 30 | 30 | SMD=-1.73 [-2.33, -1.14] | very low | |
| | ESWT vs SESWT | Pain intensity, stiffness and physical function (Western Ontario and McMaster Universities Arthritis Index) | 1 | 30 | 30 | SMD=-0.59 [-1.11, -0.08] | very low | SESWT: shockwave of the same sound without energy stimulation |
| | ESWT vs SESWT | Symptom (Lequesne Index) | 1 | 30 | 30 | SMD=-0.92 [-1.45, -0.39] | very low | |
| | ESWT vs MT | Pain intensity (Visual Analogue Scale) | 1 | 30 | 30 | SMD=-1.84 [-2.45, -1.23] | very low | |
| | ESWT vs MT | Pain intensity, stiffness and physical function (Western Ontario and McMaster Universities Arthritis Index) | 1 | 30 | 30 | SMD=-1.56 [-2.15, -0.98] | very low | MT: glucosamine |
| | ESWT vs MT | Symptom (Lequesne Index) | 1 | 30 | 30 | SMD=-1.69 [-2.29, -1.10] | very low | |
| | **Frozen shoulder** | | | | | | | |
| | ESWT + TCMET vs TCMET | Pain intensity (Visual Analogue Scale) | 1 | 23 | 23 | SMD=-0.62 [-1.21, -0.03] | very low | TCMET: small needle-knife therapy |
| | ESWT + TCMET vs TCMET | Function (Constant-Murley Score) | 1 | 23 | 23 | SMD=1.49 [0.83, 2.15] | very low | |
| | **Chronic prostatitis/ chronic pelvic pain syndrome** | | | | | | | |
| | ESWT + TCMET vs TCMET | Pain intensity, urinary symptoms, and quality of life (National Institutes of Health-Chronic Prostatitis Symptom Index Total Score) | 1 | 14 | 14 | SMD=-1.90 [-2.81, -0.98] | very low | |
| | ESWT + TCMET vs TCMET | Symptom (International Prostatism Symptom Score) | 1 | 14 | 14 | SMD=-1.27 [-2.10, -0.45] | very low | TCMET: herbal fumigation |
| | ESWT + TCMET vs TCMET | Pain intensity (Visual Analogue Scale) | 1 | 14 | 14 | SMD=-3.59 [-4.84, -2.33] | very low | |
| | **Calcific tendinitis of the shoulder** | | | | | | | |
| | ESWT + TCMET + RH vs TCMET + RH | Pain intensity (Visual Analogue Scale) | 1 | 16 | 12 | SMD=-2.45 [-3.47, -1.43] | very low | TCMET: manual acupuncture; RH: infrared therapy |
| | ESWT + TCMET + RH vs TCMET + RH | Function (Constant-Murley Score) | 1 | 16 | 12 | SMD=1.51 [0.65, 2.37] | very low | |
| | ESWT + ST vs ST | Pain intensity (Visual Analogue Scale) | 1 | 30 | 30 | SMD=-3.39 [-4.20, -2.59] | very low | ST: ultrasound-guided puncture decompression |
| | ESWT + ST vs ST | Function (University of California at Los Angeles Shoulder Score) | 1 | 30 | 30 | SMD=3.44 [2.63, 4.25] | very low | |
| | **Osteonecrosis of the femoral head** | | | | | | | |
| | ESWT + TCMIT vs TCMIT | Function and symptom (Harris Hip Score) | 1 | 24 | 24 | SMD=1.89 [0.60, 3.18] | very low | TCMIT: yishenxiaotongwan |
| | ESWT + RH + MT vs MT | Function and symptom (Harris Hip Score) | 1 | 31 | 31 | SMD=0.61 [0.10, 1.12] | very low | RH: hyperbaric oxygen therapy; MT: celecoxib |

#### Large or Moderate Effect with Low or Very low Certainty

**Low energy**

| | | | | | | | |
|---|---|---|---|---|---|---|---|
| **Post-stroke shoulder-hand syndrome** | | | | | | | |
| ESWT + TCMET vs TCMET | Pain intensity (Visual Analogue Scale) | 1 | 45 | 45 | SMD=-1.32 [-1.78, -0.87] | low | TCMET: manual acupuncture |
| ESWT + TCMET vs TCMET | Funtion (Fugl-Meyer Assessment Scale) | 1 | 45 | 45 | SMD=2.45 [1.90, 3.00] | low | |
| **Second-degree burn** | | | | | | | |
| ESWT + SC vs SC | Structure (Wound-Healing Time) | 1 | 22 | 22 | SMD=-1.45 [-2.12, -0.78] | low | SC: burn wound debridement and topical antiseptic therapy |
| ESWT + SC vs SC | Structure (Wound Infection Rate) | 1 | 22 | 22 | RR=0.67 [0.12, 3.61] | low | |
| **Plantar fasciitis** | | | | | | | |
| ESWT vs DI | Pain intensity (Visual Analogue Scale) | 2 | 203 | 208 | SMD=-1.92 [-3.72, -0.12] | low | DI: methylprednisolone or lidocaine or betamethasone |
| **Rotator cuff tear** | | | | | | | |
| ESWT + RH vs RH | Pain intensity (Visual Analogue Scale) | 1 | 30 | 32 | SMD=-0.83 [-1.36, -0.31] | very low | RH: functional training |
| ESWT + RH vs RH | Function (Constant-Murley Score) | 1 | 30 | 32 | SMD=1.00 [0.47, 1.53] | very low | |
| **Low back pain** | | | | | | | |
| ESWT vs RH | Pain intensity (Visual Analogue Scale) | 1 | 35 | 35 | SMD=-0.59 [-1.11, -0.07] | very low | RH: bridging exercises or adductor ball squeeze exercises or abdominal marching exercises or reverse curl exercises |
| ESWT vs RH | Function (Oswestry Disability Index) | 1 | 50 | 50 | SMD=-0.70 [-1.18, -0.21] | very low | |
| ESWT + RH vs RH | Pain intensity (Visual Analogue Scale) | 1 | 13 | 15 | SMD=-1.17 [-1.98, -0.35] | very low | RH: hot pack therapy and ultrasound therapy and electrotherapy |
| **Stenosing tenosynovitis** | | | | | | | |
| ESWT + RH vs RH | Funtion (Cooney Score) | 1 | 41 | 40 | SMD=0.99 [0.67, 1.32] | very low | RH: ultrashort wave electrotherapy |
| **Knee osteoarthritis** | | | | | | | |
| ESWT vs RH | Pain intensity (Visual Analogue Scale) | 1 | 35 | 35 | SMD=-1.65 [-2.19, -1.10] | very low | RH: general exercises |
| ESWT vs RH | Pain intensity, stiffness and physical function (Western Ontario and McMaster Universities Arthritis Index) | 1 | 35 | 35 | SMD=-2.13 [-2.72, -1.54] | very low | |
| ESWT vs RH | Symptom (Lequesne Index) | 1 | 35 | 35 | SMD=-1.36 [-1.88, -0.84] | very low | |
| **Patellar tendinopathy** | | | | | | | |
| ESWT vs RH | Pain intensity (Visual Analogue Scale) | 1 | 38 | 16 | SMD=-4.66 [-7.32, -1.99] | very low | RH: massage |
| ESWT vs TCMET | Pain intensity (Visual Analogue Scale) | 1 | 38 | 9 | SMD=-6.92 [-10.37, -3.47] | very low | TCMET: acupuncture |
| ESWT vs DI | Pain intensity (Visual Analogue Scale) | 1 | 38 | 4 | SMD=5.02 [3.49, 6.55] | very low | DI: prednisone and procaine |
| **Carpal tunnel syndrome** | | | | | | | |
| ESWT + MT vs MT | Funtion and symptom (Boston Carpal Tunnel Syndrome Questionnaire) | 1 | 47 | 45 | SMD=-2.85 [-3.27, -2.44] | very low | RH: neural mobilization; MT: methylcobalamin |
| **Frozen shoulder** | | | | | | | |
| ESWT + MT vs RH + MT | Pain intensity (Visual Analogue Scale) | 1 | 15 | 15 | SMD=-1.65 [-2.50, -0.81] | very low | RH: conservative physical therapy; MT: nonsteroidal anti-inflammatory drugs |
| ESWT + MT vs RH + MT | Function (Patient Specific Functional Scale) | 1 | 15 | 15 | SMD=-2.11 [-3.03, -1.19] | very low | |
| ESWT + RH vs RH | Pain intensity (Visual Analogue Scale) | 2 | 117 | 117 | SMD=-1.74 [-2.85, -0.63] | very low | RH: joint mobilization or functional exercises or pulsed radiofrequency |
| ESWT + RH vs RH | Funtion (Range of motion) | 2 | 117 | 117 | SMD=1.13 [0.90, 1.37] | very low | |
| ESWT + RH vs RH | Quality of life (36-item Short-Form) | 2 | 95 | 95 | SMD=2.00 [1.54, 2.45] | very low | |
| ESWT + RH vs RH | Function (University of California at Los Angeles Shoulder Score) | 1 | 57 | 57 | SMD=1.25 [0.85, 1.65] | very low | |
| ESWT + RH vs RH | Funtion and pain intensity (Shoulder Pain and Disability Index) | 1 | 38 | 38 | SMD=-4.06 [-4.86, -3.26] | very low | |
| **Decubitus ulceration** | | | | | | | |
| ESWT + RH vs RH | Structure (Percentage of the wound healing area) | 1 | 15 | 15 | SMD=1.35 [0.88, 1.82] | very low | RH: infrared polarized light therapy |

| | | | | | | | | |
|---|---|---|---|---|---|---|---|---|
| Low energy | **Post-stroke upper limb spasticity** | | | | | | | |
| | ESWT + RH + TCMET vs RH + TCMET | Funtion (Fugl-Meyer Assessment Scale) | 1 | 30 | 30 | SMD=1.20 [0.64, 1.75] | very low | RH: occupational therapy; TCMET: manual acupuncture |
| | ESWT + RH + TCMET vs RH + TCMET | Quality of life (Modified Barthel Index) | 1 | 30 | 30 | SMD=0.94 [0.40, 1.47] | very low | RH: occupational therapy; TCMET: manual acupuncture |
| | **Post-stroke lower limb spasticity** | | | | | | | |
| | ESWT + RH vs RH | Symptom (Modified Ashworth Scale) | 1 | 50 | 50 | SMD=-1.21 [-2.02, -0.40] | very low | RH: physiotherapy and exercises |
| | ESWT + RH vs RH | Funtion (Fugl-Meyer Assessment Scale) | 1 | 50 | 50 | SMD=0.62 [0.33, 0.90] | very low | |

Note: C, control group; CI, confidence interval; CoE, certainty of evidence assessed using the Grading of Recommendations Assessment, Development and Evaluation approach; DI, drug injection; ESWT, extracorporeal shockwave therapy; MT, medication therapy; RCTs, randomised controlled trials; RH, rehabilitation; RR, relative risk; SC, standard care; SESWT, sham extracorporeal shockwave therapy; SMD, standardized mean difference; T, treatment group; TCMET, traditional Chinese medicine external therapy; TCMIT, traditional Chinese medicine internal therapy.
