## Supplementary Methods S1 for "Effectiveness and Safety of Type- and Energy-based Extracorporeal Shockwave Therapy in Clinical Practice: Umbrella Review and Evidence Mapping"

### Supplementary Methods S1 Classification of energy level, effect size and study conclusion

Energy level,

Classification of energy level through energy flux density or pressure field.

| Energy level | Physical parameter | Specific dosage (mJ/mm$^2$ or bar) |
|---|---|---|
| High energy | Energy flux density | $0.28 \leqslant x < 0.60$ |
| | Pressure field | $x > 2.5$ |
| Medium energy | Energy flux density | $0.08 \leqslant y < 0.28$ |
| | Pressure field | $1.5 \leqslant y \leqslant 2.5$ |
| Low energy | Energy flux density | $z < 0.08$ |
| | Pressure field | $z < 1.5$ |

Note: x, high energy; y, medium energy; z, low energy.

Effect size,

Classification of effect size through specific scope regarding risk ratio and standardised mean difference.

| Estimated summary effect | Effect size | Specific scope |
|---|---|---|
| Relative risk | Large | $a \geqslant 2.0$ |
| | | $a \leqslant 0.5$ |
| | Small | $0.5 < a < 2.0$ |
| Standardized mean difference | Large | $b \geqslant 0.8$ |
| | | $b \leqslant -0.8$ |
| | Moderate | $0.5 \leqslant b < 0.8$ |
| | | $-0.8 < b \leqslant -0.5$ |
| | Small | $0.2 \leqslant b < 0.5$ |
| | | $-0.5 < b \leqslant -0.2$ |
| | Very small | $-0.2 < b < 0.2$ |

Note: a, relative risk; b, standardized mean difference.

Study conclusion,

Classification of study conclusion corresponding to each outcome.

| Conclusion class | Description |
|---|---|
| Positive | Results reported a positive effect compared to the control group and conclusion indicated that the intervention was effective. |
| Potentially positive | Results reported a positive or potentially positive effect compared to the control group and conclusion indicated that the intervention was potentially effective. |
| Unclear | Results reported a effect or not compared to the control group and conclusion did not indicate the intervention. |
| No effect | Results reported no effect compared to the control group and conclusion indicated the intervention was invalid. |
| Potentially negative | Results reported a negative or potentially negative effect compared to the control group and conclusion indicated that the intervention was potentially adverse. |
| Negative | Results reported a negative effect compared to the control group and conclusion indicated that the intervention was adverse. |
