## Supplementary Table S1 for "Effectiveness and Safety of Type- and Energy-based Extracorporeal Shockwave Therapy in Clinical Practice: Umbrella Review and Evidence Mapping"

### Supplementary Table S1 Characteristics of included systematic reviews

| First author / Year | Country | Disease or condition | No of primary studies | No of participants | Outcomes included in SRs | Conclusion from SRs | Quality assessment tool | GRADE tool |
|---|---|---|---|---|---|---|---|---|
| Ju Chan Kim 2019 | Korea | Carpal tunnel syndrome | 6 | 281 | Visual analogue scale | Positive | Cochrane risk of bias tool | not inform |
| | | | | | Boston carpal tunnel questionnaire | Positive | | |
| | | | | | Electrodiagnostic parameter | Positive | | |
| | | | | | Roles and maudsley scale | Positive | | |
| Wenhao Li 2020 | China | Carpal tunnel syndrome | 5 | 204 | Visual analogue scale | Unclear | Cochrane risk of bias tool | inform |
| | | | | | Boston carpal tunnel questionnaire | Unclear | | |
| | | | | | Sensory distal latency | Unclear | | |
| | | | | | Motor distal latency | Negative | | |
| | | | | | Compound muscle action potential amplitude | Positive | | |
| | | | | | Sensory nerve action potential amplitude | Positive | | |
| | | | | | Sensory nerve conduction velocity | Unclear | | |
| Yujie Xie 2020 | China | Carpal tunnel syndrome | 10 | 433 | Visual analogue scale | Positive | Cochrane risk of bias tool and Physiotherapy evidence database scale | not inform |
| | | | | | Boston carpal tunnel questionnaire | Positive | | |
| | | | | | Electrodiagnostic parameter | Unclear | | |
| Ko-Ta Chen 2022 | China | Carpal tunnel syndrome | 7 | 376 | Boston carpal tunnel questionnaire | Unclear | Cochrane risk of bias tool | not inform |
| | | | | | Visual analogue scale | Unclear | | |
| | | | | | Distal motor latency of median nerve | Unclear | | |
| | | | | | Sensory nerve conduction velocity | Unclear | | |
| | | | | | Incidence of adverse reaction | Positive | | |
| Qiangru Huang 2020 | China | Diabetic foot ulcers | 8 | 339 | Wound healing area | Positive | Cochrane risk of bias tool | not inform |
| | | | | | Complete cure rate | Positive | | |
| | | | | | Percentage of re-epithelialization | Positive | | |
| Zaid Munir 2023 | the UK | Peripheral arterial disease | 4 | 228 | Pain-free walking distance | Positive | Cochrane risk of bias tool | not inform |
| | | | | | Maximum walking distance | Positive | | |
| | | | | | Ankle brachial pressure index | Unclear | | |
| | | | | | Degree of arterial stenosis | Positive | | |
| L Bisset 2005 | Australia | Lateral epicondylitis | 2 | 317 | Visual analogue scale | No effect | Physiotherapy evidence database scale | not inform |
| | | | | | Grip strength | No effect | | |
| | | | | | Global improvement | No effect | | |
| Christoph Weber 2015 | Germany | Lateral epicondylitis | 4 | 327 | Overall pain relief | Potentially positive | Sign checklist | not inform |
| | | | | | Pain relief during maximum handgrip strength test | Potentially positive | | |
| | | | | | Grip strength | Potentially positive | | |
| Eli T. Sayegh 2015 | the USA | Lateral epicondylitis | 7 | 641 | Patient-rated tennis elbow evaluation | No effect | 22-point consolidated standards of reporting trials checklist | not inform |
| | | | | | Pain-free function index | No effect | | |
| | | | | | Overall function | No effect | | |
| | | | | | Disabilities of the arm, shoulder and hand score | No effect | | |

| | | | | | | | | |
|---|---|---|---|---|---|---|---|---|
| | | | | | EuroQol five dimensions questionnaire | No effect | | |
| | | | | | Grip strength | No effect | | |
| | | | | | Pain-free walking distance | No effect | | |
| Yuan Xiong 2019 | China | Lateral epicondylitis | 4 | 237 | Grip strength | Positive | Cochrane risk of bias tool | not inform |
| | | | | | Visual analogue scale | Positive | | |
| Gaowen Yao 2020 | China | Lateral epicondylitis | 13 | 1035 | Grip strength | Positive | Cochrane risk of bias tool | not inform |
| | | | | | Visual analogue scale | Positive | | |
| Seo Yeon Yoon 2020 | Korea | Lateral epicondylitis | 12 | 1104 | Grip strength | No effect | Cochrane risk of bias tool | not inform |
| | | | | | Visual analogue scale | No effect | | |
| Punam Patel 2018 | the USA | Lateral epicondylitis | 5 | not inform | Visual analogue scale | Positive | Physiotherapy evidence database scale | not inform |
| | | | | | Grip strength | No effect | | |
| You J. Kim 2021 | the USA | Lateral epicondylitis | 6 | 481 | Pain reduction | Unclear | Cochrane risk of bias tool | not inform |
| | | | | | Grip Strength | Unclear | | |
| | | | | | Patient-reported tennis elbow evaluation | Unclear | | |
| | | | | | Disabilities of the arm, shoulder and hand score | Unclear | | |
| Amarpal S. Cheema 2023 | Canada | Lateral epicondylitis | 2 | 129 | Visual analogue scale | No effect | Cochrane risk of bias tool | inform |
| | | | | | Disabilities of the arm, shoulder and hand score | No effect | | |
| Chenchen Yan 2019 | China | Lateral epicondylitis | 5 | 233 | Visual analogue scale | Positive | Cochrane risk of bias tool and Jadad score | not inform |
| | | | | | Grip Strength | Positive | | |
| | | | | | Elbow function evaluation score | Unclear | | |
| Chenxiao Zheng 2020 | China | Lateral epicondylitis | 9 | 715 | Visual analogue scale | No effect | Jadad score | not inform |
| | | | | | Pain reduction | Positive | | |
| | | | | | Thomsen test | Unclear | | |
| | | | | | Grip strength | Positive | | |
| Stefanos Karanasios 2021 | Greece | Lateral epicondylitis | 27 | 1871 | Grip strength | Unclear | Physiotherapy evidence database scale | inform |
| | | | | | Visual analogue scale | Unclear | | |
| Zongye Zhong 2018 | China | Lateral epicondylitis | 10 | 928 | Visual analogue scale | Positive | Cochrane risk of bias tool | not inform |
| | | | | | Grip strength | Positive | | |
| Yunfei Yuan 2023 | China | Lateral epicondylitis | 10 | 646 | Visual analogue scale | Positive | Cochrane risk of bias tool | not inform |
| | | | | | Grip strength | Potentially positive | | |
| | | | | | Incidence of recurrence | Potentially positive | | |
| | | | | | Upper limb function score | Potentially positive | | |
| Shuai Wang 2015 | China | Lateral epicondylitis | 5 | 389 | Visual analogue scale | Unclear | Jadad score | not inform |
| Zhengan Hao 2015 | China | Lateral epicondylitis | 11 | 708 | Visual analogue scale | Positive | Cochrane risk of bias tool | not inform |
| | | | | | Upper limb function score | Positive | | |
| | | | | | Roles and maudsley scale | Positive | | |
| | | | | | Incidence of adverse reaction | Positive | | |
| Lezheng Wang 2020 | China | Lateral epicondylitis | 9 | 481 | Visual analogue scale | Positive | Cochrane risk of bias tool and Jadad score | not inform |

| | | | | | | | | |
|---|---|---|---|---|---|---|---|---|
| | | | | | Grip strength | Unclear | | |
| Samuel P Sussmilch-Leitch 201 | Australia | Achilles tendinopathy | 5 | 290 | Visual analogue scale | Potentially positive | Physiotherapy evidence database scale | not inform |
| | | | | | Victorian institute of sports assessment–achilles questionnaire | Potentially positive | | |
| Yifei Fan 2020 | China | Achilles tendinopathy | 8 | 442 | Numerical rating scale | Positive | Cochrane risk of bias tool | not inform |
| | | | | | Visual analogue scale | Positive | | |
| | | | | | Roles and maudsley scale | Positive | | |
| | | | | | Likert scale | Positive | | |
| | | | | | Victorian institute of sports assessment–achilles questionnaire | Positive | | |
| | | | | | American orthopaedic foot and ankle society scale | Positive | | |
| | | | | | Tenderness score | Unclear | | |
| | | | | | Pain pressure threshold | Unclear | | |
| Magdalena Stania 2023 | Poland | Achilles tendinopathy | 6 | 344 | Numerical rating scale | Unclear | Physiotherapy evidence database scale | inform |
| | | | | | Victorian institute of sports assessment–achilles questionnaire | Unclear | | |
| Li Zhiyun 2013 | China | Plantar fasciitis | 5 | 716 | Visual analogue scale | Positive | Jadad score | not inform |
| Jan Natbleen C. Dizon 2013 | Philippines | Plantar fasciitis | 11 | 1384 | Visual analogue scale | Positive | Physiotherapy evidence database scale | not inform |
| | | | | | Roles and maudsley scale | Positive | | |
| Meng-Chen Yin 2014 | China | Plantar fasciitis | 7 | 550 | Visual analogue scale | Unclear | Cochrane risk of bias tool | not inform |
| | | | | | Roles and maudsley scale | Positive | | |
| Jiale Sun 2017 | China | Plantar fasciitis | 9 | 935 | Visual analogue scale | Potentially positive | Cochrane risk of bias tool | not inform |
| Shuxiang Li 2018 | China | Plantar fasciitis | 9 | 658 | Visual analogue scale | Positive | Cochrane risk of bias tool | inform |
| | | | | | Incidence of recurrence | Unclear | | |
| Chien-Min Chen 2018 | China | Plantar fasciitis | 5 | 369 | Visual analogue scale | Potentially negative | Jadad score | not inform |
| Ying-Chun Wang 2019 | China | Plantar fasciitis | 14 | 1392 | Visual analogue scale | Unclear | Cochrane risk of bias tool | not inform |
| Yuan Xiong 2019 | China | Plantar fasciitis | 6 | 454 | Thickness score | Unclear | Cochrane risk of bias tool and Jadad score | not inform |
| | | | | | Visual analogue scale | Positive | | |
| Hui Li 2019 | China | Plantar fasciitis | 5 | 177 | American orthopaedic foot and ankle society scale | Unclear | Cochrane risk of bias tool and Jadad score | not inform |
| | | | | | Visual analogue scale | Positive | | |
| | | | | | Foot function index | Unclear | | |
| | | | | | Pain and disability scale | Unclear | | |
| Zeyana Al-Siyabi 2022 | The Sultanate of Omar | Plantar fasciitis | 7 | 369 | American orthopaedic foot and ankle society scale | Unclear | Cochrane risk of bias tool | not inform |
| | | | | | Visual analogue scale | Potentially positive | | |
| | | | | | Functional impairment | Unclear | | |
| | | | | | Activity limitation | Positive | | |
| | | | | | Patient satisfaction | Positive | | |
| | | | | | Thickness score | Unclear | | |
| Janice de S. Guimarães 2023 | Brazil | Plantar fasciitis | 30 | 3096 | Visual analogue scale | Positive | Cochrane risk of bias tool | not inform |
| Gao Ning 2022 | China | Plantar fasciitis | 16 | 1198 | Visual analogue scale | Potentially positive | Cochrane risk of bias tool | not inform |
| | | | | | Thickness score | Potentially positive | | |

| | | | | | | | | |
|---|---|---|---|---|---|---|---|---|
| | | | | | Maximum continuous walking time | Potentially positive | | |
| Chen Ke-cun 2022 | China | Plantar fasciitis | 22 | 2240 | Foot function index | Unclear | Cochrane risk of bias tool and Jadad score | not inform |
| | | | | | Visual analogue scale | Positive | | |
| | | | | | Roles and maudsley scale | Positive | | |
| Zhou Yinan 2020 | China | Plantar fasciitis | 8 | 1489 | Visual analogue scale | Positive | Cochrane risk of bias tool | not inform |
| | | | | | Roles and maudsley scale | Positive | | |
| João Vitor Ferlito 2023 | Brazil | Plantar fasciitis | 19 | 1089 | Visual analogue scale | Potentially negative | Physiotherapy evidence database scale | inform |
| | | | | | Foot function index | Potentially positive | | |
| | | | | | American orthopaedic foot and ankle society scale | Potentially positive | | |
| Jing Lou 2017 | China | Plantar fasciitis | 9 | 1174 | Visual analogue scale | Potentially positive | Cochrane risk of bias tool | not inform |
| | | | | | Roles and maudsley scale | Potentially positive | | |
| Ruedi Steuri 2017 | Switzerland | Shoulder impingement | 3 | 117 | Visual analogue scale | Potentially positive | Cochrane risk of bias tool | inform |
| Stefano Salvioli 2017 | Italy | Plantar heel pain | 10 | 1114 | Visual analogue scale | Unclear | Cochrane risk of bias tool | inform |
| | | | | | Numerical rating scale | Unclear | | |
| BaoLin Li 2018 | China | Plantar heel pain | 3 | 312 | Visual analogue scale | Positive | Cochrane risk of bias tool | not inform |
| Yangquan Hao 2018 | China | Osteonecrosis of the femoral head | 4 | 230 | Visual analogue scale | Unclear | Cochrane risk of bias tool | not inform |
| | | | | | Harris hip score | Potentially positive | | |
| Jin Mei 2022 | China | Osteonecrosis of the femoral head | 9 | 409 | Harris hip score | Potentially positive | National institute for clinical excellence case series scoring, Newcastle-Ottawa scale and Jadad score | not inform |
| | | | | | Visual analogue scale | Potentially positive | | |
| | | | | | Magnetic resonance imaging | Potentially positive | | |
| JiaBin Li 2022 | China | Osteonecrosis of the femoral head | 14 | 860 | Harris hip score | Positive | Cochrane risk of bias tool | not inform |
| | | | | | Visual analogue scale | Positive | | |
| | | | | | Activity of daily living scale | Positive | | |
| HE Xiangbo 2021 | China | Osteonecrosis of the femoral head | 6 | 334 | Harris hip score | Positive | Cochrane risk of bias tool | not inform |
| Jie Li 2023 | China | Osteonecrosis of the femoral head | 11 | 780 | Harris hip score | Positive | Cochrane risk of bias tool | not inform |
| Chun-De Liao 2018 | China | Lower limb tendinopathy | 29 | 1865 | Visual analogue scale | Positive | Physiotherapy evidence database scale | not inform |
| | | | | | Function recovery | positive | | |
| Mohamed H. Elgendy 2023 | Egypt | Upper and lower limb tendinopathies | 22 | 1789 | Visual analogue scale | No effect | Cochrane risk of bias tool | not inform |
| | | | | | Disability of the arm, shoulder and hand score | No effect | | |
| Spencer Jonathan Lee 2017 | the USA | Patellar tendinopathy | 5 | 120 | Visual analogue scale | Positive | Physiotherapy evidence database scale | not inform |
| | | | | | Victorian institute of sport australia–patella questionnaire | Positive | | |
| Luca Andriolo 2018 | Italy | Patellar tendinopathy | 6 | 261 | Victorian institute of sport australia–patella questionnaire | Positive | Coleman methodology score | not inform |
| Magdalena Stania 2022 | Poland | Patellar tendinopathy | 7 | 311 | Visual analogue scale | Unclear | Physiotherapy evidence database scale | not inform |
| | | | | | Victorian institute of sport australia–patella questionnaire | Unclear | | |
| Chun-De Liao 2018 | China | Knee tendinopathies and other soft tissue disorders | 19 | 1189 | Visual analogue scale | Positive | Cochrane risk of bias tool and Physiotherapy evidence database scale | not inform |
| | | | | | Roles and maudsley scale | Positive | | |
| | | | | | Likert scale | Positive | | |
| | | | | | Incidence of adverse reaction | Positive | | |

| | | | | | | | | |
|---|---|---|---|---|---|---|---|---|
| | | | | | Range of motion | Positive | | |
| | | | | | Victorian institute of sport australia–patella questionnaire | Positive | | |
| Li Zhang 2017 | China | Acute and chronic soft tissue wounds | 10 | 473 | Wound healing rate | Positive | Cochrane risk of bias tool | not inform |
| | | | | | Wound healing time | Positive | | |
| | | | | | Percentage of the wound-healing area | Positive | | |
| | | | | | Incidence of adverse reaction | Positive | | |
| | | | | | Wound infection rate | Positive | | |
| Li Tengqi 2019 | China | Knee osteoarthritis | 7 | 366 | Visual analogue scale | Potentially positive | Cochrane risk of bias tool | not inform |
| | | | | | Range of motion | Potentially positive | | |
| | | | | | Lequesne index | Potentially positive | | |
| | | | | | Western ontario and mcmaster universities osteoarthritis index | Potentially positive | | |
| Ricardo Maia Ferreira 2019 | Portugal | Knee osteoarthritis | 1 | 105 | Visual analogue scale | Unclear | Physiotherapy evidence database scale | inform |
| | | | | | Western ontario and mcmaster universities osteoarthritis index | Unclear | | |
| Chi-Kun Hsieh 2020 | China | Knee osteoarthritis | 9 | 705 | Visual analogue scale | Potentially positive | Strengthening the reporting of observational studies in epidemiology statement | not inform |
| | | | | | Western ontario and mcmaster universities osteoarthritis index | Potentially positive | | |
| Ying-Chun Wang 2019 | China | Knee osteoarthritis | 9 | 431 | Visual analogue scale | Positive | Cochrane risk of bias tool | not inform |
| | | | | | Western ontario and mcmaster universities osteoarthritis index | Positive | | |
| Huanzhi Ma 2020 | China | Knee osteoarthritis | 6 | 589 | Visual analogue scale | Positive | Cochrane risk of bias tool | inform |
| | | | | | Western ontario and mcmaster universities osteoarthritis index | Positive | | |
| | | | | | Lequesne index | Positive | | |
| | | | | | Incidence of adverse reaction | Positive | | |
| Juan Avendaño-Coy 2020 | Spain | Knee osteoarthritis | 14 | 782 | Visual analogue scale | Positive | Cochrane risk of bias tool | inform |
| | | | | | Western ontario and mcmaster universities osteoarthritis index | Positive | | |
| | | | | | Range of motion | Positive | | |
| | | | | | Walking test | Positive | | |
| Lu Chen 2020 | China | Knee osteoarthritis | 32 | 2408 | Visual analogue scale | Positive | Cochrane risk of bias tool | not inform |
| | | | | | Western ontario and mcmaster universities osteoarthritis index | Positive | | |
| Zhimin Huangfu 2020 | China | Knee osteoarthritis | 21 | 1736 | Visual analogue scale | Positive | Cochrane risk of bias tool | inform |
| | | | | | Western ontario and mcmaster universities osteoarthritis index | Positive | | |
| | | | | | Range of motion | Positive | | |
| Dengcheng Huang 2020 | China | Knee osteoarthritis | 12 | 1040 | Visual analogue scale | Positive | Cochrane risk of bias tool and Jadad score | not inform |
| | | | | | Western ontario and mcmaster universities osteoarthritis index | Positive | | |
| | | | | | Lequesne index | Unclear | | |
| Shaoyong Yu 2020 | China | Knee osteoarthritis | 11 | 969 | Visual analogue scale | Positive | Cochrane risk of bias tool | not inform |
| | | | | | Western ontario and mcmaster universities osteoarthritis index | Positive | | |
| Qiang Hu 2021 | China | Knee osteoarthritis | 16 | 1259 | Visual analogue scale | Positive | Cochrane risk of bias tool | not inform |
| | | | | | Western ontario and mcmaster universities osteoarthritis index | Positive | | |
| | | | | | Lysholm score | Positive | | |

| | | | | | | | | | |
|---|---|---|---|---|---|---|---|---|---|
| | | | | | | Lequesne index | Positive | | |
| Jinyu Gu 2022 | China | Knee osteoarthritis | 13 | 1132 | | Visual analogue scale | Positive | Cochrane risk of bias tool | not inform |
| | | | | | | Western ontario and mcmaster universities osteoarthritis index | Positive | | |
| | | | | | | Lysholm score | Positive | | |
| | | | | | | Incidence of adverse reaction | Unclear | | |
| Zhipeng Ma 2023 | China | Knee osteoarthritis | 12 | 1043 | | Visual analogue scale | Positive | Cochrane risk of bias tool | not inform |
| | | | | | | Western ontario and mcmaster universities osteoarthritis index | Positive | | |
| AC Silva 2023 | Brazil | Knee osteoarthritis | 12 | 734 | | Visual analogue scale | Potentially positive | Physiotherapy evidence database scale | inform |
| | | | | | | Western ontario and mcmaster universities osteoarthritis index | Potentially positive | | |
| | | | | | | Knee injury and osteoarthritis outcome score | Potentially positive | | |
| | | | | | | Lequesne index | Potentially positive | | |
| Sofia Oliveira 2022 | Portugal | Knee osteoarthritis | 20 | 1414 | | Visual analogue scale | Positive | Cochrane risk of bias tool | inform |
| | | | | | | Western ontario and mcmaster universities osteoarthritis index | Positive | | |
| | | | | | | Lequesne index | Positive | | |
| Jayson Lian 2018 | the USA | Enthesopathy of the extensor carpi radialis brevis | 3 | 263 | | Visual analogue scale | Unclear | Jadad score | not inform |
| | | | | | | Grip Strength | Unclear | | |
| | | | | | | Incidence of adverse reaction | Unclear | | |
| Hani Al-Abbad 2020 | Australia | Musculoskeletal condition | 27 | 3110 | | Average particle size of the calcium deposition | Unclear | Cochrane risk of bias tool | not inform |
| | | | | | | Total calcification resorption | Unclear | | |
| | | | | | | Plantar fascial thickness | Unclear | | |
| | | | | | | Femoral head necrosis lesion size | Unclear | | |
| Ruihan Zhang 2021 | China | Frozen shoulder | 34 | 2293 | | Constant-murley score | Positive | Cochrane risk of bias tool | not inform |
| | | | | | | Visual analogue scale | Positive | | |
| | | | | | | External rotation range of motion | Unclear | | |
| | | | | | | Incidence of adverse reaction | Positive | | |
| Zhu Chang-e 2017 | China | Frozen shoulder | 11 | 816 | | Visual analogue scale | Positive | Cochrane risk of bias tool | not inform |
| | | | | | | Range of motion | Positive | | |
| Hou Chengzhi 2019 | China | Frozen shoulder | 17 | 1449 | | Visual analogue scale | Positive | Cochrane risk of bias tool | not inform |
| | | | | | | Shoulder pain and disability index | Positive | | |
| | | | | | | Range of motion | Positive | | |
| HAN Nahui 2020 | China | Frozen shoulder | 20 | 1404 | | Visual analogue scale | Positive | Jadad score | not inform |
| | | | | | | Constant-murley score | Positive | | |
| | | | | | | Range of motion | Positive | | |
| GUAN Fuxi 2018 | China | Adhesive capsulitis | 15 | 1063 | | Constant-murley score | Positive | Cochrane risk of bias tool | not inform |
| | | | | | | Visual analogue scale | Positive | | |
| Patrick Vavken 2009 | the USA | Calcific tendinitis of the shoulder | 14 | 995 | | Visual analogue scale | Positive | Jadad score | not inform |
| | | | | | | Constant-murley score | Positive | | |
| Francesco Ioppolo 2013 | Italy | Calcific tendinitis of the shoulder | 6 | 460 | | Total calcification resorption | Positive | Physiotherapy evidence database scale | not inform |

| | | | | | | | | |
|---|---|---|---|---|---|---|---|---|
| | | | | | Total calcification resorption | Positive | | |
| | | | | | Partial calcification resorption | Positive | | |
| | | | | | Visual analogue scale | Positive | | |
| | | | | | Constant-murley score | Positive | | |
| Jan K.G. Louwerens 2014 | Netherland | Calcific tendinitis of the shoulder | 20 | 1544 | Visual analogue scale | Positive | Cochrane risk of bias tool | inform |
| | | | | | Constant-murley score | Positive | | |
| | | | | | Total calcification resorption | Positive | | |
| Qianyuan WANG 2017 | China | Calcific tendinitis of the shoulder | 5 | 302 | Visual analogue scale | Positive | Jadad score | not inform |
| | | | | | Constant-murley score | Positive | | |
| | | | | | Total calcification resorption | Positive | | |
| | | | | | Average particle size of the calcium deposition | Positive | | |
| HAN Xuan 2021 | China | Rotator cuff tendinopathy | 11 | 529 | Visual analogue scale | Unclear | Physiotherapy evidence database scale | not inform |
| | | | | | Constant-murley score | Positive | | |
| | | | | | Incidence of adverse reaction | Potentially positive | | |
| Chen Mingfeng 2022 | China | Rotator cuff tear | 14 | 881 | Visual analogue scale | Positive | Cochrane risk of bias tool | not inform |
| | | | | | Constant-murley score | Positive | | |
| | | | | | Incidence of adverse reaction | Positive | | |
| | | | | | Range of motion | Positive | | |
| | | | | | University of california at los angeles shoulder rating scale | Positive | | |
| Hunter S. Angileri 2023 | the USA | Rotator cuff tendinopathy | 12 | 1128 | Visual analogue scale | Potentially positive | Cochrane risk of bias tool | not inform |
| | | | | | Constant-murley score | Potentially positive | | |
| | | | | | Total calcification resorption | Potentially positive | | |
| Lei Yue 2021 | China | Low back pain | 10 | 455 | Visual analogue scale | Positive | Cochrane risk of bias tool and Jadad score | inform |
| | | | | | Oswestry disability index | Positive | | |
| | | | | | Numeric rating scale | Positive | | |
| | | | | | Pain self-efficacy questionnaire | Positive | | |
| | | | | | Incidence of adverse reaction | Positive | | |
| Chunhong Li 2022 | China | Low back pain | 13 | 648 | Visual analogue scale | Positive | Cochrane risk of bias tool | inform |
| | | | | | Oswestry disability index | Positive | | |
| | | | | | Incidence of adverse reaction | Positive | | |
| Jinhui Ma 2020 | China | Low back pain | 5 | 222 | Visual analogue scale | Positive | Cochrane risk of bias tool | inform |
| | | | | | Numeric rating scale | Positive | | |
| | | | | | Oswestry disability index | Positive | | |
| | | | | | Incidence of adverse reaction | Positive | | |
| Kun Liu 2023 | China | Low back pain | 12 | 632 | Visual analogue scale | Positive | Cochrane risk of bias tool | inform |
| | | | | | Oswestry disability index | Positive | | |
| | | | | | Mental health score | Unclear | | |
| Su Muyang 2022 | China | Myofascial pain syndrome | 10 | 611 | Incidence of adverse reaction | Positive | Cochrane risk of bias tool | not inform |

| | | | | | | | | |
|---|---|---|---|---|---|---|---|---|
| | | | | | Visual analogue scale | Positive | | |
| | | | | | Oswestry disability index | Positive | | |
| | | | | | Pain pressure threshold | Positive | | |
| Ji Hyun Jun 2021 | Korea | Myofascial pain syndrome | 11 | 505 | Visual analogue scale | Positive | Cochrane risk of bias tool | not inform |
| | | | | | Pain pressure threshold | Positive | | |
| | | | | | Neck disability index | Unclear | | |
| Tao Wu 2022 | China | Myofascial pain syndrome | 8 | 571 | Visual analogue scale | Positive | Physiotherapy evidence database scale | not inform |
| | | | | | Pain pressure threshold | Positive | | |
| | | | | | Neck disability index | Unclear | | |
| Qing Zhang 2020 | China | Myofascial pain syndrome | 10 | 477 | Visual analogue scale | Positive | Cochrane risk of bias tool and physiotherapy evidence database scale | not inform |
| | | | | | Pain pressure threshold | No effect | | |
| | | | | | Neck disability index | No effect | | |
| Jun-Il Yoo 2020 | Korea | Myofascial pain syndrome | 5 | 371 | Visual analogue scale | Potentially positive | Cochrane risk of bias tool | not inform |
| Jin Ran 2017 | China | Myofascial pain syndrome | 8 | 370 | Visual analogue scale | Positive | Cochrane risk of bias tool | not inform |
| Carlos Avendaño-López 2023 | France | Myofascial pain syndrome | 27 | 1377 | Visual analogue scale | Positive | Cochrane risk of bias tool | inform |
| | | | | | Function recovery | Positive | | |
| | | | | | Pain pressure threshold | Positive | | |
| | | | | | Range of motion | Unclear | | |
| | | | | | 36-item short-form | Unclear | | |
| Ge Rui 2013 | China | Tendon enthesiopathy | 6 | 599 | Visual analogue scale | Positive | Jadad score | not inform |
| | | | | | Incidence of adverse reaction | Positive | | |
| Chengzhi Hou 2019 | China | Stenosing tenosynovitis | 7 | 387 | Visual analogue scale | Positive | Cochrane risk of bias tool | not inform |
| | | | | | Cooney score | Positive | | |
| Zimu Hu 2023 | China | Post-stroked shoulder-hand syndrome | 7 | 494 | Visual analogue scale | Positive | Cochrane risk of bias tool | not inform |
| | | | | | Stroke-specific quality of life scale | Positive | | |
| | | | | | Fugl-meyer assessment | Positive | | |
| Tingyu Zhang 2023 | China | Post-stroked shoulder-hand syndrome | 18 | 1248 | Visual analogue scale | Positive | Cochrane risk of bias tool | not inform |
| | | | | | Fugl-meyer assessment | Positive | | |
| | | | | | Range of motion | Positive | | |
| | | | | | Functional comprehensive assessment score | Positive | | |
| L Gao 2016 | China | Peyronie’s disease | 6 | 443 | Plaque size | Potentially positive | Not inform | not inform |
| | | | | | Penile curvature | No effect | | |
| | | | | | Pain reduction | Potentially positive | | |
| | | | | | Intercourse function | No effect | | |
| Ahmed M.Bakr 2021 | Egypt | Peyronie’s disease | 3 | 238 | Penile curvature | No effect | Cochrane risk of bias tool | not inform |
| | | | | | Visual analogue scale | No effect | | |
| | | | | | International index of erectile function-erectile function domain score | Unclear | | |
| Hesong Jiang 2017 | China | Peyronie’s disease | 8 | 930 | Plaque size | Potentially positive | Jadad score | not inform |

| | | | | | | | | |
|---|---|---|---|---|---|---|---|---|
| | | | | | Penile pain reduction | Positive | | |
| | | | | | Plaque size | Positive | | |
| | | | | | Penile curvature | Positive | | |
| | | | | | Intercourse function | Positive | | |
| Zhihua vLu 2016 | the USA | Erectile dysfunction | 14 | 833 | International index of erectile function-erectile function domain score | Positive | Cochrane risk of bias tool | not inform |
| | | | | | Erection hardness score | Positive | | |
| Zi-jun Zou 2017 | China | Erectile dysfunction | 15 | 695 | International index of erectile function-erectile function domain score | Potentially positive | Cochrane risk of bias tool and Methodological index for non-randomized studies | not inform |
| | | | | | Erection hardness score | Potentially positive | | |
| J.C.Angulo 2016 | Spain | Erectile dysfunction | 12 | 636 | International index of erectile function-erectile function domain score | Positive | Not inform | not inform |
| MAN Libo 2017 | China | Erectile dysfunction | 9 | 637 | International index of erectile function-erectile function domain score | Positive | Cochrane risk of bias tool | not inform |
| | | | | | Erection hardness score | Positive | | |
| Liang Dong 2019 | China | Erectile dysfunction | 7 | 522 | International index of erectile function-erectile function domain score | Positive | Cochrane risk of bias tool | not inform |
| | | | | | Erection hardness score | Positive | | |
| Ioannis Sokolakis 2019 | Germany | Erectile dysfunction | 10 | 873 | International index of erectile function-erectile function domain score | Positive | Cochrane risk of bias tool | not inform |
| | | | | | Peak systolic velocity | Positive | | |
| | | | | | Erection hardness score | Positive | | |
| MO Dun-sheng 2019 | China | Erectile dysfunction | 8 | 595 | International index of erectile function-erectile function domain score | Positive | Jadad score | not inform |
| | | | | | Erection hardness score | Positive | | |
| Jeffrey D.Campbell 2019 | Canada | Erectile dysfunction | 7 | 607 | International index of erectile function-erectile function domain score | Positive | Cochrane risk of bias tool | not inform |
| | | | | | Erection hardness score | Positive | | |
| | | | | | Incidence of adverse reaction | Positive | | |
| Huibao Yao 2022 | China | Erectile dysfunction | 16 | 1064 | International index of erectile function-erectile function domain score | Positive | Cochrane risk of bias tool | not inform |
| | | | | | Erection hardness score | Unclear | | |
| Junbo Liu 2020 | China | Erectile dysfunction | 10 | 697 | International index of erectile function-erectile function domain score | Positive | Cochrane risk of bias tool | not inform |
| | | | | | Erection hardness score | Positive | | |
| Wu Tao 2019 | China | Erectile dysfunction | 10 | 697 | International index of erectile function-erectile function domain score | Positive | Cochrane risk of bias tool | not inform |
| | | | | | Erection hardness score | Positive | | |
| Beom Yong Rho 2022 | Korea | Erectile dysfunction | 5 | 460 | International index of erectile function-erectile function domain score | Potentially positive | Cochrane risk of bias tool and Newcastle–Ottawa scale | inform |
| Raul I. Clavijo 2017 | the USA | Erectile dysfunction | 7 | 602 | International index of erectile function-erectile function domain score | Positive | Cochrane risk of bias tool | not inform |
| Dariusz Kałka 2021 | Poland | Erectile dysfunction | 5 | 354 | International index of erectile function-erectile function domain score | Positive | Cochrane risk of bias tool | not inform |
| | | | | | Erection hardness score | Positive | | |
| | | | | | Flow-mediated dilatation | Positive | | |
| LIAO Bo 2019 | China | Chronic prostatitis/chronic pelvic pain syndrome | 12 | 838 | National institute of health-chronic prostatitis symptom index total score | Positive | Cochrane risk of bias tool | not inform |
| | | | | | National institute of health-chronic prostatitis symptom index pain score | Positive | | |
| | | | | | National institute of health-chronic prostatitis symptom index urinary symptom score | Positive | | |
| | | | | | National institute of health-chronic prostatitis symptom index quality of life score | Positive | | |
| Penghui Yuan 2019 | China | Chronic prostatitis/chronic pelvic pain syndrome | 5 | 280 | National institute of health-chronic prostatitis symptom index total score | Positive | Cochrane risk of bias tool | not inform |
| | | | | | National institute of health-chronic prostatitis symptom index pain score | Positive | | |

| | | | | | | | | |
|---|---|---|---|---|---|---|---|---|
| | | | | | National institute of health-chronic prostatitis symptom index urinary symptom score | Positive | | |
| | | | | | National institute of health-chronic prostatitis symptom index quality of life score | Positive | | |
| Ioannis Mykoniatis 2021 | Germany | Chronic prostatitis/chronic pelvic pain syndrome | 6 | 316 | Numerical rating scale | Positive | Cochrane risk of bias tool | inform |
| | | | | | National institute of health-chronic prostatitis symptom index total score | Positive | | |
| | | | | | National institute of health-chronic prostatitis symptom index pain score | Positive | | |
| | | | | | National institute of health-chronic prostatitis symptom index urinary symptom score | Unclear | | |
| | | | | | National institute of health-chronic prostatitis symptom index quality of life score | Positive | | |
| | | | | | International index of erectile function-erectile function domain score | Unclear | | |
| | | | | | International prostate symptom score | Unclear | | |
| | | | | | Post-void residual urine volume | Unclear | | |
| | | | | | Maximum urinary flow rate | Unclear | | |
| Guizhong Li 2021 | China | Chronic prostatitis/chronic pelvic pain syndrome | 6 | 317 | National institute of health-chronic prostatitis symptom index total score | Potentially positive | Cochrane risk of bias tool | not inform |
| | | | | | National institute of health-chronic prostatitis symptom index urinary symptom score | Potentially positive | | |
| | | | | | National institute of health-chronic prostatitis symptom index quality of life score | Potentially positive | | |
| | | | | | Visual analogue scale | Potentially positive | | |
| GE Jinchao 2015 | China | Chronic prostatitis/chronic pelvic pain syndrome | 8 | 592 | National institute of health-chronic prostatitis symptom index total score | Positive | Cochrane risk of bias tool | not inform |
| Guoxian Deng 2020 | China | Chronic non-bacterial prostatitis | 4 | 428 | National institute of health-chronic prostatitis symptom index total score | Positive | Cochrane risk of bias tool | not inform |
| Ponco Birowo 2020 | Indonesia | Chronic non-bacterial prostatitis | 3 | 137 | National institute of health-chronic prostatitis symptom index total score | Positive | Cochrane risk of bias tool | not inform |
| | | | | | National institute of health-chronic prostatitis symptom index pain score | Positive | | |
| | | | | | National institute of health-chronic prostatitis symptom index urinary symptom score | Positive | | |
| | | | | | National institute of health-chronic prostatitis symptom index quality of life score | Positive | | |
| Jin-Youn Lee 2014 | Korea | Post-stroke spasticity | 5 | 117 | Modified ashworth scale | Positive | Cochrane risk of bias tool | not inform |
| Peipei Guo 2017 | China | Post-stroke spasticity | 6 | 301 | Modified ashworth scale | Positive | Jadad score and Newcastle–Ottawa scale | not inform |
| | | | | | Incidence of adverse reaction | Positive | | |
| Jie XIANG 2018 | China | Post-stroke spasticity | 8 | 385 | Modified ashworth scale | Positive | Cochrane risk of bias tool | not inform |
| | | | | | Modified tardieu scale | Positive | | |
| | | | | | Range of motion | Positive | | |
| | | | | | Hmax/Mmax ratio | Positive | | |
| Davood Azimpour 2017 | Iran | Post-stroke spasticity | 11 | 261 | Modified ashworth scale | Positive | Physiotherapy evidence database scale | not inform |
| | | | | | Passive rang of movement | Positive | | |
| Gongwei Jia 2020 | China | Post-stroke spasticity | 8 | 301 | Modified ashworth scale | Positive | Cochrane risk of bias tool | not inform |
| | | | | | Visual analogue scale | Positive | | |
| | | | | | Range of motion | Positive | | |
| | | | | | Fugl-meyer assessment | Positive | | |
| | | | | | Incidence of adverse reaction | Positive | | |
| Liang-Jun Ou-Yang 2023 | China | Post-stroke spasticity | 13 | 667 | Modified ashworth scale | Positive | Cochrane risk of bias tool | not inform |
| | | | | | Modified tardieu scale | Positive | | |
| Mohammad Etoom 2018 | Jordan | Multiple sclerosis spasticity | 1 | 64 | Fugl-meyer assessment | Positive | Physiotherapy evidence database scale | inform |

| | | | | | | | | |
|---|---|---|---|---|---|---|---|---|
| | | | | | Modified ashworth scale | Unclear | | |
| | | | | | Visual analogue scale | Unclear | | |
| | | | | | Hmax/Mmax ratio | Unclear | | |
| Jae Ho Oh 2019 | Korea | Spasticity | 9 | 385 | Modified ashworth scale | Positive | Cochrane risk of bias tool | not inform |
| Emanuela Elena Mihai 2021 | Romania | Post-stroke lower limb spasticity | 7 | 170 | Modified ashworth scale | Positive | Physiotherapy evidence database scale | not inform |
| | | | | | Modified tardieu scale | Positive | | |
| | | | | | Visual analogue scale | Positive | | |
| | | | | | Hmax/Mmax ratio | Unclear | | |
| | | | | | Passive rang of movement | Positive | | |
| | | | | | Timed up and go test | Unclear | | |
| Rosa Cabanas-Valdés 2019 | Spain | Post-stroke lower limb spasticity | 12 | 278 | Modified ashworth scale | Positive | Physiotherapy evidence database scale | not inform |
| | | | | | Range of motion | Positive | | |
| | | | | | Fugl-meyer assessment | Positive | | |
| | | | | | Incidence of adverse reaction | Positive | | |
| Jiabao Guo 2017 | China | Post-stroke lower limb spasticity | 6 | 192 | Modified ashworth scale | Positive | Cochrane risk of bias tool | not inform |
| | | | | | Fugl-meyer assessment | Positive | | |
| Rosa Cabanas-Valdés 2020 | Spain | Post-stroke upper limb spasticity | 16 | 764 | Modified ashworth scale | Positive | Physiotherapy evidence database scale | not inform |
| | | | | | Fugl-meyer assessment | Positive | | |
| | | | | | Visual analogue scale | Unclear | | |
| Hui-Ling Zhang 2022 | China | Post-stroke upper limb spasticity | 42 | 1973 | Modified ashworth scale | Potentially positive | Cochrane risk of bias tool | inform |
| | | | | | Incidence of adverse reaction | Potentially positive | | |
| LIU Guoqing 2021 | China | Cerebral palsy spasticity | 7 | 350 | Modified ashworth scale | Positive | Cochrane risk of bias tool | not inform |
| | | | | | Range of motion | Positive | | |
| | | | | | Gross motor function measure-88 | Positive | | |
| | | | | | Plantar surface area | Positive | | |
| | | | | | Step length and pace | Positive | | |
| Min Cheol Chang 2023 | Korea | Cerebral palsy spasticity | 3 | 47 | Modified ashworth scale | Positive | Cochrane risk of bias tool | not inform |
| | | | | | Range of motion | Positive | | |
| | | | | | Plantar surface area | Positive | | |
| Yingying Guo 2022 | China | Cerebral palsy spasticity | 8 | 475 | Modified ashworth scale | Positive | Cochrane risk of bias tool | not inform |
| | | | | | Gross motor function measure-88 | Positive | | |
| | | | | | Range of motion | Positive | | |
| | | | | | Plantar surface area | Positive | | |
| | | | | | Plantar surface pressure | Positive | | |
| | | | | | Incidence of adverse reaction | Positive | | |
| Hyun-Jung KIM 2019 | Korea | Cerebral palsy spasticity | 5 | 104 | Modified ashworth scale | Potentially positive | Physiotherapy evidence database scale | not inform |
| | | | | | Range of motion | Potentially positive | | |
| | | | | | Plantar surface area | Potentially positive | | |

| | | | | | | | | |
|---|---|---|---|---|---|---|---|---|
| | | | | | Plantar surface pressure | Potentially positive | | |
| Haiyang Wu 2023 | China | Cervical spondylotic radiculopathy | 9 | 610 | Visual analogue scale | Positive | Cochrane risk of bias tool and Jadad score | not inform |
| | | | | | Neck disability index | Positive | | |
| | | | | | Pressure test score | No effect | | |
| | | | | | Median nerve F wave conduction velocity | Positive | | |
| Li Zhang 2017 | China | Chronic wounds | 7 | 301 | Wound healing rate | Positive | Jadad score | not inform |
| | | | | | Wound healing area | Positive | | |
| | | | | | Wound healing time | Positive | | |
| | | | | | Wound infection rate | Positive | | |
| | | | | | Incidence of adverse reaction | Positive | | |
| Jonathan Häußer 2021 | Germany | Bone marrow oedema | 6 | 168 | Visual analogue scale | Potentially positive | Downs and Black checklist | not inform |
| | | | | | Function recovery | Potentially positive | | |
| Yu Lin Tsai 2021 | China | Breast cancer-related lymphedema | 8 | 274 | Volume of lymphedema | Positive | Cochrane risk of bias tool, Jadad score and Physiotherapy evidence database scale | inform |
| | | | | | Arm circumference | Positive | | |
| | | | | | Skin thickness | Positive | | |
| | | | | | Range of motion | Positive | | |
| | | | | | Disability of the arm, shoulder and hand score | Positive | | |
| Yanhui Yang 2022 | China | Post-burn pathological scar | 9 | 422 | Numerical rating scale | Positive | Cochrane risk of bias tool | not inform |
| | | | | | Scar thickness | Positive | | |
| | | | | | Trans-epidermal water loss | Positive | | |
| | | | | | Vancouver scar scale | Positive | | |
| Farshad Nikouei 2022 | Iran | Coccydynia | 4 | 81 | Visual analogue scale | Positive | Cochrane risk of bias tool | not inform |
| Diogo Simões Fonseca 2023 | Indonesia | Patellar tendinopathy | unknown | unknown | Visual analogue scale | unknown | Physiotherapy evidence database scale | inform |
| | | | | | Victorian institute of sport australia–patella questionnaire | | | |
| Yang Han 2022 | China | Lumbar disc herniation | unknown | unknown | Curative effect | unknown | Cochrane risk of bias tool | not inform |
| | | | | | Visual analogue scale | | | |
| | | | | | Japanese orthopaedic association score | | | |
| | | | | | Oswestry disability index | | | |
| Yan Chenchen 2019 | China | Lateral epicondylitis | unknown | unknown | Visual analogue scale | unknown | Cochrane risk of bias tool | not inform |
| | | | | | Evaluation of the elbow function | | | |
| LiTing Wang 2023 | China | Knee osteoarthritis | unknown | unknown | Visual analogue scale | unknown | Cochrane risk of bias tool | not inform |
| | | | | | Western ontario and mcmaster universities osteoarthritis index | | | |
| Lezheng Wang 2019 | China | Lateral epicondylitis | unknown | unknown | Visual analogue scale | unknown | Cochrane risk of bias tool | not inform |
| | | | | | Grip strength | | | |
| Xiaoxi Mou 2020 | China | Achilles tendinopathy | unknown | unknown | Visual analogue scale | unknown | Cochrane risk of bias tool and Newcastle-Ottawa scale | not inform |
| | | | | | Numerical rating scale | | | |
| | | | | | Victorian institute of sports assessment–achilles questionnaire | | | |
| Rocky Nurakbariansyah 2020 | Indonesia | Chronic prostatitis/chronic pelvic pain syndrome | unknown | unknown | American orthopaedic foot and ankle society scale | unknown | Cochrane risk of bias tool | not inform |

| | | | | | | | | |
|---|---|---|---|---|---|---|---|---|
| | | | | | National institute of health-chronic prostatitis symptom index total score<br>National institute of health-chronic prostatitis symptom index pain score<br>National institute of health-chronic prostatitis symptom index urinary symptom score<br>National institute of health-chronic prostatitis symptom index quality of life score | | | |
| Shan Xie 2023 | China | Spasticity after brain injury | unknown | unknown | Modified ashworth scale<br>Modified tardieu scale<br>Motor impairment evaluation<br>Activities of daily living<br>Pain evaluation<br>Electrophysiological parameters<br>Indicators of the economic cost<br>Incidence of adverse reaction<br>Dropout rate | unknown | Cochrane risk of bias tool | not inform |
| Kim Da jeong 2023 | South Korea | Chronic musculoskeletal disorders | unknown | unknown | Pain evaluation<br>Function evaluation | unknown | Cochrane risk of bias tool | not inform |
| Kiyeun Nam 2017 | South Korea | Cerebral palsy spasticity | unknown | unknown | Modified ashworth scale<br>Range of motion<br>Plantar surface area<br>Peak pressure value of heel | unknown | Physiotherapy evidence database scale | not inform |
| Xue Xiali 2023 | China | Rotator cuff tear | unknown | unknown | Visual analogue scale<br>Constant-murley score<br>University of california at los angeles shoulder rating scale<br>Range of motion<br>American shoulder and elbow surgeons form | unknown | Cochrane risk of bias tool | not inform |
| Chaitanya J 2022 | India | Cerebral palsy spasticity | unknown | unknown | Modified ashworth scale<br>Modified tardieu scale<br>Range of motion<br>Passive and reflex mediated stiffness | unknown | Cochrane risk of bias tool and Physiotherapy evidence database scale | not inform |
| Lun-Xue QING 2018 | China | Knee osteoarthritis | unknown | unknown | Pain intensity evaluation<br>Quality of life<br>Function status | unknown | Cochrane risk of bias tool | not inform |
| LUO ZHIQIANG 2023 | China | Lumbar disc herniation | unknown | unknown | Visual analogue scale<br>Japanese orthopaedic association score<br>Oswestry disability index<br>Recurrence rate<br>Numerical rating scale | unknown | Cochrane risk of bias tool and Jadad score | not inform |
| Yan Yan 2021 | China | Low back pain | unknown | unknown | Incidence of adverse reaction<br>Visual analogue scale | unknown | Physiotherapy evidence database scale | inform |

| | | | | | | | | |
|---|---|---|---|---|---|---|---|---|
| | | | | | Numerical rating scale<br>Oswestry disability index<br>Incidence of adverse reaction | | | |
| Wang Fangqi 2022 | China | Subacromial shoulder pain | unknown | unknown | Visual analogue scale<br>Numerical rating scale<br>Constant-murley score<br>Shoulder pain and disability Index<br>Disabilities of the arm, shoulder and hand score<br>Health assessment questionnaire<br>Simple shoulder test<br>Shoulder disability questionnaire | unknown | Physiotherapy evidence database scale | not inform |
| Jia Chen 2023 | China | Bone tissue disease | unknown | unknown | Visual analogue scale<br>Lequesne index<br>Western ontario and mcmaster universities osteoarthritis index<br>Harris hip score<br>Range of motion<br>Bone healing rate<br>Femandezesteve radiological evaluation<br>Overall response rate<br>Fracture healing time | unknown | Cochrane risk of bias tool | not inform |
| IRIS OTERO LUIS 2023 | Spain | Spasticity | unknown | unknown | Espasticity evaluation | unknown | Cochrane risk of bias tool | not inform |
| Samah omara 2023 | Egypt | Plantar fasciitis | unknown | unknown | Visual analogue scale<br>Foot function index<br>Heel tenderness index<br>Pain pressure threshold<br>American orthopaedic foot and ankle society scale<br>Flexor digitorum brevis thickness<br>Skin blood flow and temperature<br>Plantar fascia thickness | unknown | Cochrane risk of bias tool | not inform |
| Patricia ventura 2020 | Brazil | Breast cancer-related lymphedema | unknown | unknown | Incidence of adverse reaction<br>Volume evaluation | unknown | Cochrane risk of bias tool and Physiotherapy evidence database scale | not inform |
| Lehua Yu 2022 | China | Post-burn pathological scar | unknown | unknown | Pain intensity evaluation<br>Pruritus evaluation<br>Scar thickness evaluation<br>Skin elasticity evaluation<br>Vascularity evaluation | unknown | Physiotherapy evidence database scale | not inform |
| Liu Shuai 2020 | China | Erectile dysfunction | unknown | unknown | Transepidermal water loss evaluation<br>International index of erectile function-erectile function domain score | unknown | Cochrane risk of bias tool | not inform |

| | | | | | | | | |
|---|---|---|---|---|---|---|---|---|
| | | | | | Erection hardness score | | | |
| Yijun Lin 2023 | China | Myofascial pain syndrome | unknown | unknown | Visual analogue scale<br>Numerical rating scale<br>Pain pressure threshold | unknown | Cochrane risk of bias tool and Physiotherapy evidence database scale | not inform |
| Ervandy Rangganata 2020 | Indonesia | Chronic non-bacterial prostatitis | unknown | unknown | Visual analogue scale<br>National institute of health-chronic prostatitis symptom index total score<br>National institute of health-chronic prostatitis symptom index urinary symptom score<br>National institute of health-chronic prostatitis symptom index quality of life score | unknown | Cochrane risk of bias tool | not inform |
| Huan Liu 2020 | China | Stenosing tenosynovitis | unknown | unknown | Pain intensity evaluation<br>Incidence of adverse reaction | unknown | Cochrane risk of bias tool | not inform |
| Li Zhang 2021 | China | Hypertrophic scars and Keloids | unknown | unknown | Size of scars<br>Pain symptoms of scars<br>Pigmentation of scars<br>Elasticity or pliability of scars<br>Related function evaluation | unknown | Cochrane risk of bias tool | not inform |
| Li Zhang 2022 | China | Post-burn pathological scar | unknown | unknown | Thickness of scars<br>Uncomfortable symptoms of scars<br>Vancouver scar scale<br>Elasticity or pliability of scars<br>Incidence of adverse reaction | unknown | Cochrane risk of bias tool | not inform |
| Kun Liu 2023 | China | Low back pain | unknown | unknown | Visual analogue scale<br>Oswestry disability index<br>Incidence of adverse reaction | unknown | Cochrane risk of bias tool | not inform |
| Apurba Barman 2020 | India | Musculoskeletal soft tissue injuries | unknown | unknown | Visual analogue scale<br>Numerical rating scale<br>Incidence of adverse reaction | unknown | Cochrane risk of bias tool | not inform |
| Yu-Chi Su 2022 | China | Plantar and palmar fibromatosis | unknown | unknown | Visual analogue scale<br>Range of motion | unknown | Cochrane risk of bias tool and Joanna Briggs Institute critical appraisal checklist | not inform |
| Hongcheng Tao 2023 | China | Osteonecrosis of the femoral head | unknown | unknown | Harris hip score<br>Visual analogue scale<br>Incidence of adverse reaction | unknown | Cochrane risk of bias tool and Newcastle-Ottawa scale | not inform |
| Kunyuan Wang 2023 | China | Rotator cuff tear | unknown | unknown | Visual analogue scale<br>Constant-murley score<br>Range of motion<br>Shoulder pain and disability index | unknown | Cochrane risk of bias tool and Jadad score | not inform |
| Yafeng Li 2020 | China | Tenosynovitis | unknown | unknown | Visual analogue scale | unknown | Cochrane risk of bias tool | not inform |
| Xin Gao 2023 | China | Delayed fracture union and nonunion | unknown | unknown | Cooney score<br>Recovery rate | unknown | Cochrane risk of bias tool and Jadad score | not inform |

| | | | | | | | | |
|---|---|---|---|---|---|---|---|---|
| | | | | | Healing time<br>Callus line score<br>Economic burden evaluation<br>Fracture gap<br>Incidence of adverse reaction | | | |
| Qiangru Huang 2018 | China | Diabetic foot ulcers | unknown | unknown | Blood flow perfusion rate<br>Average wound healing time<br>Numeric box scale<br>Visual analogue scale | unknown | Cochrane risk of bias tool | not inform |
| Yu Nong Ao 2018 | China | Knee osteoarthritis | unknown | unknown | Visual analogue scale<br>Range of motion<br>Western ontario and mcmaster universities osteoarthritis index | unknown | Cochrane risk of bias tool and Physiotherapy evidence database scale | not inform |
| Sayed Anvar 2022 | India | Musculoskeletal conditions | unknown | unknown | Visual analogue scale<br>Grip strength<br>Pinch strength<br>Foot function index<br>Quick disability of arm, shoulder and hand | unknown | Cochrane risk of bias tool and Physiotherapy evidence database scale | not inform |
| Zhuorao Wu 2023 | China | Low back pain | unknown | unknown | Finger-floor distance<br>Visual analogue scale<br>Japanese orthopaedic association score<br>Oswestry disability index<br>Beck depression inventory<br>Incidence of adverse reaction | unknown | Cochrane risk of bias tool | not inform |
| Juan Avendaño-Coy 2022 | Spain | Myofascial pain syndrome | unknown | unknown | Pain intensity evaluation<br>Pressure pain threshold<br>Funcionality evaluation<br>Range of motion<br>Incidence of adverse reaction<br>Quality of life | unknown | Cochrane risk of bias tool | not inform |
| Shanshan Liu 2022 | China | Temporomandibular joint disorder | unknown | unknown | Visual analogue scale<br>Maximum mouth opening<br>Disability evaluation | unknown | Physiotherapy evidence database scale | not inform |
| Bijan Forogh 2022 | Iran | Calcific tendinitis of the shoulder | unknown | unknown | Pain intensity evaluation<br>Range of motion | unknown | National Heart, Lung, and Blood Institute study quality assessment tool | not inform |
| Kai LI 2020 | China | Post-stroke upper limb spasticity | unknown | unknown | Modified ashworth scale<br>Modified tardieu scale | unknown | Not inform | not inform |
| Fangqi Wang 2022 | China | Rotator cuff tendinopathy | unknown | unknown | Visual analogue scale<br>Numerical rating scale | unknown | Physiotherapy evidence database scale | not inform |

| | | | | | | | | |
|---|---|---|---|---|---|---|---|---|
| | | | | | Constant-murley score<br>Shoulder pain and disability Index<br>Disabilities of the arm, shoulder and hand score<br>Health assessment questionnaire<br>Simple shoulder test<br>Shoulder disability questionnaire | | | |
| Grzegorz Fojecki 2015 | Denmark | Urological diseases | unknown | unknown | Visual analogue scale<br>Penile curvature<br>Quality of life<br>International index of erectile function-erectile function domain score<br>Erection hardness score | unknown | Not inform | not inform |
| Shristi Shakya 2023 | India | Cerebral palsy spasticity | unknown | unknown | Head control<br>Trunk control<br>Head and trunk control<br>Gross motor function<br>Activities of daily living | unknown | Physiotherapy evidence database scale | not inform |
| Hong li Xu 2023 | China | Low back pain | unknown | unknown | Pain intensity evaluation<br>Oswestry disability index<br>Beck depression scale<br>Finger-floor distance<br>36-item short-form | unknown | Cochrane risk of bias tool | not inform |
| Chaoqun Feng 2023 | China | Plantar fasciitis | unknown | unknown | Curative effect<br>Pain intensity evaluation<br>Function evaluation | unknown | Cochrane risk of bias tool | not inform |
| Xiangyu Zhu 2022 | China | Achilles tendinopathy | unknown | unknown | Visual analogue scale | unknown | Cochrane risk of bias tool | not inform |
| Jin Mei 2020 | China | Osteonecrosis of the femoral head | unknown | unknown | Harris hip score<br>Visual analogue scale<br>Radiography evaluation | unknown | National Institute for Clinical Excellence case series scoring standard tool, Newcastle-Ottawa scale and Jadad score | not inform |
| Jun-Il Yoo 2019 | South Korea | Myofascial pain syndrome | unknown | unknown | Visual analogue scale<br>Neck disability index<br>Constant-murley score | unknown | Cochrane risk of bias tool | not inform |
| Yueting Wang 2022 | China | Postpartum dysfunction | unknown | unknown | Abdominal circumference<br>Rectus abdominis separation index<br>Visual analogue scale<br>Electromyography evaluation<br>Sexual satisfaction | unknown | Cochrane risk of bias tool | not inform |
| Wang-Sheng Lin 2021 | China | Breast cancer-related lymphedema | unknown | unknown | Female sexual function index<br>Volume evaluation | unknown | Not inform | not inform |

| | | | | | | | | |
|---|---|---|---|---|---|---|---|---|
| | | | | | Circumference evaluation<br>Bioelectrical impedance<br>Skin thickness<br>Visual analogue scale<br>Quick disability of arm, shoulder and hand | | | |
| Xinchao Shi 2020 | China | Patellar tendinopathy | unknown | unknown | Visual analogue scale<br>Numerical rating scale<br>Victorian institute of sport australia–patella questionnaire | unknown | Physiotherapy evidence database scale | not inform |
| Yu Qin 2020 | China | Low back pain | unknown | unknown | Pain intensity evaluation<br>Functional evaluation<br>Quality of life<br>Psychological evaluation<br>Incidence of adverse reaction | unknown | Cochrane risk of bias tool | not inform |
| Anuj Punnoose 2013 | England(UK) | Lower limb tendinopathy | unknown | unknown | Visual analogue scale<br>Numerical rating scale<br>Victorian institute of sport australia–patella questionnaire<br>Victorian institute of sports assessment–achilles questionnaire<br>Tendon thickness<br>Range of motion<br>Strength evaluation | unknown | Physiotherapy evidence database scale | not inform |
| Rosa Cabanas-Valdes 2018 | Spain | Post-stroke upper limb spasticity | unknown | unknown | Modified ashworth scale<br>Modified tardieu scale<br>Range of motion<br>F waves and H-reflex latency<br>Motricity, dexterity, pain, muscle spams evaluation | unknown | Physiotherapy evidence database scale | not inform |
| Danyang liu 2019 | China | Spasticity after upper motor neuron injury | unknown | unknown | Modified ashworth scale<br>Gross motor function measurement<br>Passive range of motion<br>Plantar pressure<br>Plantar area | unknown | Cochrane risk of bias tool | not inform |
| Wei Changhao 2020 | China | Knee osteoarthritis | unknown | unknown | Visual analogue scale<br>Western ontario and mcmaster universities osteoarthritis index<br>Lysholm score<br>Incidence of adverse reaction | unknown | Cochrane risk of bias tool | not inform |
| Lezheng Wang 2020 | China | Chronic prostatitis/chronic pelvic pain syndrome | unknown | unknown | International index of erectile function-erectile function domain score<br>Maximum urinary flow rate<br>Post void residual volume<br>Visual analogue scale | unknown | Cochrane risk of bias tool and Jadad score | not inform |

| | | | | | | | | |
|---|---|---|---|---|---|---|---|---|
| | | | | | International prognostic scoring system<br>National institute of health-chronic prostatitis symptom index total score<br>National institute of health-chronic prostatitis symptom index pain score<br>National institute of health-chronic prostatitis symptom index urinary symptom score<br>National institute of health-chronic prostatitis symptom index quality of life score | | | |
| Chao Li 2023 | China | Post-stroke lower limb spasticity | unknown | unknown | Modified ashworth scale<br>Modified tardieu scale<br>Passive range of motion<br>Fugl-meyer assessment<br>Incidence of adverse reaction | unknown | Cochrane risk of bias tool | not inform |
| Xiaofeng Wang 2023 | China | Peyronie’s Disease | unknown | unknown | Penile curvature reduction<br>Proportion of patients with plate reduction<br>Visual analogue scale<br>Incidence of adverse reaction<br>Self-made scale<br>International index of erectile function-erectile function domain score | unknown | Cochrane risk of bias tool | not inform |
| Lu Chen 2019 | China | Osteoarthritis | unknown | unknown | Visual analogue scale<br>Western ontario and mcmaster universities osteoarthritis index | unknown | Newcastle-Ottawa scale | not inform |

Note: GRADE, Grading of Recommendations Assessment, Development and Evaluation; SRs, systematic reviews.
