## Supplementary Table S2 for "Effectiveness and Safety of Type- and Energy-based Extracorporeal Shockwave Therapy in Clinical Practice: Umbrella Review and Evidence Mapping"

**Supplementary Table S2 Number of randomised controlled trials reporting specific adverse reactions related to system organ class and preferred term**

| System organ class | Preferred term | Number of RCTs (%) |
|---|---|---|
| General disorders and administration site conditions | Pain | 35 (16.5) |
| | Flushing | 26 (12.3) |
| | Swelling | 18 (8.5) |
| | Oedema | 3 (1.4) |
| | Pyrexia | 3 (1.4) |
| | Tenderness | 2 (1.0) |
| | Fatigue | 1 (0.5) |
| | Discomfort | 1 (0.5) |
| Skin and subcutaneous tissue disorders | Paraesthesia | 7 (3.3) |
| | Hypoaesthesia | 7 (3.3) |
| | Skin discomfort | 7 (3.3) |
| | Contusion | 6 (2.8) |
| | Erythema | 5 (2.4) |
| | Rash | 2 (1.0) |
| | Skin injury | 1 (0.5) |
| | Skin abrasion | 1 (0.5) |
| | Alopecia | 1 (0.5) |
| | Skin irritation | 1 (0.5) |
| | Hyperhidrosis | 1 (0.5) |
| | Pallor | 1 (0.5) |
| | Skin burning sensation | 1 (0.5) |
| | Sensitive skin | 1 (0.5) |
| Vascular disorders | Haematoma | 9 (4.3) |
| | Skin haemorrhage | 8 (3.8) |
| | Ecchymosis | 8 (3.8) |
| | Petechiae | 7 (3.3) |
| | Orthostatic hypotension | 1 (0.5) |
| | Haematuria | 1 (0.5) |
| | Haematospermia | 1 (0.5) |
| | Penile haemorrhage | 1 (0.5) |
| | Vasodilatation | 1 (0.5) |
| Nervous system disorders | Dizziness | 5 (2.4) |
| | Headache | 4 (1.9) |
| | Neuropathy peripheral | 2 (1.0) |
| | Dyssomnia | 1 (0.5) |
| | Migraine | 1 (0.5) |
| | Tremor | 1 (0.5) |
| | Syncope | 1 (0.5) |
| Gastrointestinal disorders | Nausea | 10 (4.7) |
| | Abdominal discomfort | 2 (1.0) |
| Musculoskeletal and connective tissue disorders | Arthralgia | 3 (1.4) |
| | Myalgia | 2 (1.0) |
| | Hypertonia | 1 (0.5) |
| | Joint dislocation | 1 (0.5) |
| | Joint effusion | 1 (0.5) |
| | Joint stiffness | 1 (0.5) |
| | Muscular weakness | 1 (0.5) |
| Infections and infestations | Infection | 2 (1.0) |
| General system disorders | Asthenia | 1 (0.5) |
| Renal and urinary disorders | Dysuria | 1 (0.5) |
| Cardiac disorders | Palpitations | 1 (0.5) |
| Immune system disorders | Hypersensitivity | 1 (0.5) |
| Respiratory, thoracic and mediastinal disorders | Nasal discomfort | 1 (0.5) |

Note: RCTs, randomised controlled trials.
